# Pandemic-risk-related behaviour change in England from June 2020 to March 2022: REACT-1 study among over 2 million people

**DOI:** 10.1101/2025.03.03.25323250

**Authors:** Nicholas Steyn, Marc Chadeau-Hyam, Matthew Whitaker, Christina Atchison, Deborah Ashby, Graham S. Cooke, Helen Ward, Paul Elliott, Christl A. Donnelly

## Abstract

**Objective:** To determine how people in England changed their infection risk-related behaviours during the COVID-19 pandemic and in response to control measures, 19 June 2020 to 31 March 2022.

**Design:** Over 18 (of 19) rounds, randomly selected participants across England completed a questionnaire about risk-related behaviours, socio-demographics, and symptoms.

**Participants:** Between 85,018 and 154,060 randomly selected participants per round, aged 5+ years, totalling 2,177,657 responses with relevant data.

**Main outcome measures:** Primary outcomes were self-reported shielding and/or taking specific precautions, not leaving home in the prior week, not being in close proximity with anyone outside their household the day before, and wearing face coverings outside the home. Secondary community-level measures of mobility and public health policy stringency were compared to the primary outcomes to provide population-level context to the observed findings.

**Results:** Infection risk-related behaviours varied considerably over the nearly two years under study. Protective behaviours peaked in January 2021, during England’s winter wave, before widespread vaccination. At that time, the estimated proportion of self-reported shielding and/or taking specific precautions reached 21.6% (95% confidence interval 21.4% to 21.8%), of self-reported not leaving home in the week prior to completing the questionnaire reached 7.99% (7.85% to 8.13%) and of self-reported not having contact with anyone outside their household on the day before answering the questionnaire reached 89.2% (89.1% to 89.4%). As self-reported vaccination rates increased and prevalence of infection decreased, protective behaviours decreased, although patterns varied by demographics. Protective behaviours were strongly correlated with community-level mobility data and the stringency of public health measures.

**Conclusions:** Individual-based data showed sizeable proportions of people undertook protective behaviours during the pandemic especially during the second lockdown in January 2021 although there was evidence of “pandemic fatigue” in the study’s later stages. Self-reported behaviours were closely aligned with community mobility data and the stringency of government policies, indicating policy-driven behaviour changes.

## Introduction

The COVID-19 pandemic in England was a complex and quickly evolving public health crisis consisting of multiple waves of infection. The REal-time Assessment of Community Transmission-1 (REACT-1) study provided real-time estimates of the prevalence of SARS-CoV-2 swab positivity in England from 1 May 2020 to 31 March 2022 (Elliott et al., 2023). Conducted over 19 rounds, more than 2.5 million nasal swab samples were processed using reverse transcriptase polymerase chain reaction (RT-PCR), and participants completed an extensive questionnaire about their behaviours related to infection risk, socio-demographic characteristics, and symptoms.

Various non-pharmaceutical interventions were utilised throughout the pandemic, including stay-at-home orders (“lockdowns”) on multiple occasions, notably in March 2020 (prior to the start of REACT-1) (Knock et al., 2021), and then in the winter of 2020/21 (around the time of REACT-1 rounds 7 to 9) (Riley et al., 2021). These interventions were mostly lifted in the middle of 2021, although some intermediate measures, such as recommended working from home, were re-introduced in December 2021 to slow the spread of the Omicron variant (Elliott, Bodinier, et al., 2022).

Changes in individual behaviours, influenced by perceived risks, governmental guidelines, societal norms, and “pandemic fatigue”, could directly impact the trajectory of the epidemic and the effectiveness of policy decisions. Understanding how these behaviours changed over the course of the pandemic is critical to quantify the impact of non-pharmaceutical interventions, to gauge their effectiveness at reducing SARS-CoV-2 transmission, and to understand how compliance varied across demographic groups and over time (Williams et al., 2020).

Community-level mobility data, such as the Google community mobility reports (Google LLC, n.d.), have been widely used in epidemiological studies to evaluate the effectiveness of social-distancing measures (Snoeijer et al., 2021) or to predict the rate of virus transmission (Nouvellet et al., 2021). These data offer valuable insights but are only aggregated proxies for individual behaviours and for social interaction.

The Oxford COVID-19 Government Response Tracker (OxCGRT) (Hale et al., 2021) collected information on a variety of policy measures that governments used during the pandemic such as school closures and workplace restrictions. In particular, their stringency index has been used for comparing the strictness of policy responses between countries, and has featured directly in epidemiological models (Askitas et al., 2021). Their containment and health index closely resembles the stringency index, with additional consideration given to contact tracing, testing, face covering policy, vaccinations, and protection of elderly people.

Focusing on self-reported behaviours relating to shielding and/or taking specific precautions, not leaving the home, not having physical contact/close proximity with people outside the household, and wearing face coverings outside the home, we estimated the self-reported prevalence of these behaviours from 19 June 2020 to 31 March 2022 in England and by socio-demographic group. We investigated the consistency of individually reported and community-level behavioural data by relating these estimates to (i) (Google) mobility data and (ii) the OxCGRT stringency index and OxCGRT containment and health index.

## Methods

### REACT-1 Data overview

The REACT-1 study conducted 19 rounds of sampling from 1 May 2020 to 31 March 2022 including between 85,018 and 154,060 participants in each round. Participants completed a comprehensive survey reporting their risk-related behaviours, socio-demographic characteristics, health and health history, history of COVID-19, and risk tolerances. The survey was completed by the respondent (if over the age of 18), by the respondent’s parent/guardian (if aged between 5 and 12), and either by the parent/guardian or by the respondent themselves (if aged between 13 and 17). A Public Advisory Panel provided input into the design, conduct, and dissemination of the REACT research program.

Of the questions on behaviours, we focused on one related to shielding (“*Are you shielding and/or taking specific precautions because you are concerned that you/your child will become severely ill with COVID-19?*”), one to leaving the home (“*Did you/your child leave home for any reason in the last 7 days?”* and later “*In the last 7 days, for what reasons have you left home? Select all that apply.*”), one to social distancing (“*Not including members of your household, how many different people did you have contact with yesterday? By contact we mean: any direct skin-to-skin physical contact (e.g kiss/embrace/handshake), being less than 2 metres from another person for over 5 minutes.*”), and one to wearing face coverings outside home (“*Do you/does your child mainly wear any kind of face covering or mask when you/they are outside your/their home, because of COVID-19?*”). The specific question wording occasionally changed between study rounds (supplementary material section 1a).

During study round 1, the relevant questions of interest were not included in questionnaire; we therefore exclude the (N=120,620) round 1 participants from this analysis.

Round-specific survey proportions and confidence intervals were weighted using round-level random iterative method (RIM) weights (Sharot, 1986) as implemented in the *survey* package in R (Lumley, 2023). This was done to correct prevalence estimates of self-reported behaviours to be representative of the population of England with respect to participants’ age, sex, area-level deciles of deprivation (McLennan et al., 2019), lower-tier local authority counts and ethnicity (Elliott et al., 2023).

Of the (N=3,239,620) REACT-1 participants in study rounds 2-to-19 (19 June 2020 – 31 March 2022), we excluded, prior to analysis, participants with invalid survey weights (N=1,019) as well as those that registered but did not complete the questionnaire (N=1,060,944). Some participants did not answer every question, provided an invalid response, or were not asked a specific question (the latter primarily being due to age). When responses to individual questions were missing, the response was discarded (set as “NA”). Hence, reported proportions are relative to the total number of respondents to each question. A round-by-round breakdown of non-response rates by question is provided in supplementary material section 1b.

### Statistical methods

Results are presented at national level, as well as for four age-groups in years (5-17, 18-34, 35-64, 65 and over) and four household sizes (lives alone, 2 people, 3-4 people, 5+ people). Additionally (supplementary material section 2), we consider geographical area, area-level deciles of deprivation (McLennan et al., 2019), ethnicity, and sex (Elliott et al., 2023).

The *svyciprop* function with the logit method is used to estimate weighted round-level binomial proportions and corresponding confidence intervals. Daily prevalence estimates were unweighted proportions because day-specific RIM weights were not available, and the corresponding confidence intervals were calculated using the *binconf* function from the *Hmisc* package (Harrell, 2024) in R.

We analyse if and to what extent responses to the behavioural questions varied by age-group, sex, geographical area, deprivation, ethnicity, and household size, via survey-weighted logistic regression models. These models were fit to data from each study round using the *svyglm* function with the binomial family from the *survey* package in R (Lumley, 2023). Results from these models are presented as odds ratios, providing the relative odds of reporting a given behaviour compared to a reference group (see figure 2 for the specification of reference groups).

### Google community mobility

Community mobility data were obtained from Google for the period 15 February 2020 to 15 October 2022 (Google LLC, n.d.). These publicly available data were aggregated from device users who had turned on the location history setting (by default it is off). These data present the day-to-day percentage change (relative to a day-of-the-week-specific baseline calculated from the five-week period from 3 January 2020 to 6 February 2020) in the total number of visitors to places of retail and recreation, grocery and pharmacy, parks, transit stations, and workplaces as well as the daily percentage change in time spent at residential locations.

To evaluate the consistency of these mobility data and individually reported behavioural data from REACT-1 assessing if and for what (potentially multiple) reason(s) participants left their home *in the past 7-days*, we fitted a random forest model for the mobility data (as the outcome) from questionnaire data (as predictor). To remove the day-of-the-week variability observed in the mobility data, these were smoothed using a trailing 7-day moving average prior to analysis. Specifically, models were implemented for the smoothed daily mobility metrics as a function of the proportion of people reporting leaving their home for various reasons (and not leaving). To limit measurement error in our predictors we only considered days when more than 200 survey responses were recorded.

Three separate models for each mobility series were constructed to reflect changes in questions and/or in question formulation across the study period. We defined three main periods (i) from 19 June 2020 to 11 November 2020 (study rounds 2-to-6, N=94 days with > 200 survey question responses, 11 predictors), (ii) 4 January 2021 to 28 September 2021 (study rounds 8-to-14, N=135 days with > 200 survey question responses, 8 predictors), and (iii) 19 October 2021 to 31 March 2022 (study rounds 15-to-19, N=104 days with > 200 survey question responses, 14 predictors). Study round 7 is not included in this analysis as the question regarding reasons for leaving the home in the past 7-days was not asked. The specific questions used in each model can be found in supplementary material section 1.

To assess model performance, we calculate the proportion of the within-bag variance explained (the proportion of variance in the response that is explained by the predictors when testing on data used to fit the models) and out-of-bag variance explained (the proportion of variance in the response that is explained when testing on data not used to fit the models). Default options from the *randomForest* package were used: 500 trees were fit each using approximately 63.2% of the available observations during training. The within-bag variance explained is analogous to R^2^ in a linear regression context.

### Oxford COVID-19 Government Response Tracker

The OxCGRT stringency index and containment and health index were obtained for the period 1 January 2020 to 31 December 2022. These data aggregate indicators measuring the implementation of school closures, workplace closures, public event cancellations, restrictions on gathering sizes, closure of public transport, stay-at-home requirements, restrictions on internal movement, restrictions on international travel, and public health information campaigns. The containment and health index additionally considers testing, contact tracing, face covering requirements, and vaccination policy.

Similar to our approach with the Google community mobility data, we evaluated the consistency of these indices with response to four self-reported behavioural questions by fitting a random forest regression model. This model was fit to cover the period 16 October 2020 to 31 March 2022 (study rounds 6 and 8-to-19, N=259 days with > 200 survey responses, 4 predictors).

We have developed an online portal to facilitate visualisation and download of aggregated behavioural data while protecting the confidentiality of survey participants [https://m-whit-ic.shinyapps.io/react-social-shiny/].

## Results

### Behaviours over time

We observed a peak in reported protective behaviours in January 2021 (REACT-1 round 8), at the time of the third national lockdown in England (Figure 1). At this time, the estimated proportion of self-reported shielding and/or taking specific precautions reached 20.8% (95% CI 20.6%, 21.1%), estimated proportion of self-reported not leaving home in the 7 days prior to completing the questionnaire reached 9.7% (95% CI 9.5%, 9.9%), and estimated proportion of self-reported not having physical contact/close proximity with anyone outside their household on the day before answering the questionnaire reached 93.3% (95% CI 92.8%, 93.7%).

**Figure 1.**
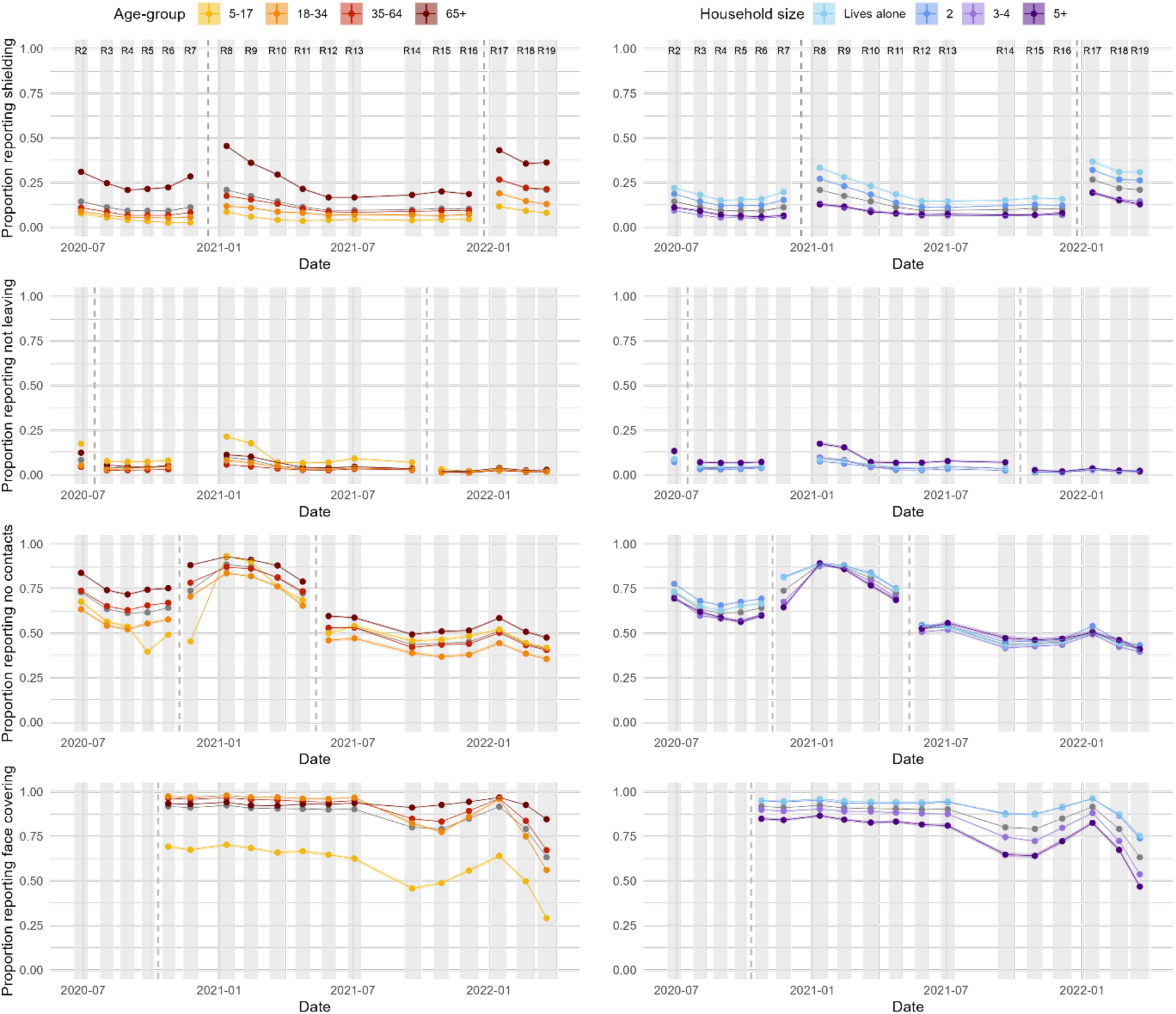
The proportion of people in England self-reporting shielding and/or taking specific precautions because they consider themselves “to be at risk of severe illness for COVID-19” (first row), not leaving the home on the day preceding answering the questionnaire (second row), having no contacts outside the home in the preceding seven days (third row), and wearing a face covering outside the home (fourth row) by age-group (left column) and household size (right column). Changes in question wording are demarked with a vertical dashed grey line – caution should be exercised in comparing values on either side. Shaded regions and vertical-coloured lines show 95% confidence intervals about the estimated proportion, although they are frequently too small to discern. Individual points are joined by lines and shading for clarity, but this should not be interpreted as an interpolation. National averages are shown in grey.

Following this peak, reported protective behaviours decreased over the summer months in 2021, with a noticeable uptick again in January 2022 (Figure 1, REACT-1 round 17), during the first Omicron wave and the government’s “Plan-B” interventions and messaging (*Prime Minister Confirms Move to Plan B in England*, 2021). This uptick in self-reported social-distancing behaviours did not reach the same level as winter 2020/21, except for the proportion of people reporting shielding and/or taking specific precautions. Some of the reported increase from December 2021 to January 2022 (REACT-1 rounds 16 to 17) in this behaviour can likely be attributed to rewording of the question (specifically, the removal of the word “shielding”, a term with a much narrower definition than “specific precautions”) (see supplementary material section 1 for details on the wording changes). Shortly after the uptick in December 2021/January 2022, protective behaviours rapidly decreased over February and March 2022 (REACT-1 rounds 18 and 19) (Elliott, Eales, et al., 2022). This is despite infection prevalence reaching the highest measured by the study in March 2022 (weighted swab positivity in round 19 was 6.37%), while the proportion self-reporting wearing face coverings reached the lowest recorded level in the study of 63.2% (95% CI 62.9%, 63.6%), down from 91.6% (95% CI 91.4%, 91.8%) in January 2022 (REACT-1 round 17) and 92.4% (95% CI 92.2%, 92.6%) in January 2021 (REACT-1 study round 8). Similarly, we observed reductions in the proportion self-reporting no contacts outside the home (41.2% (95% CI 40.9%, 41.6%) in March 2022, down from 51.3% (95% CI 50.9%, 51.7%) in January 2022 and 88.6% (95% CI 88.3%, 88.8%) in January 2021.

Figure 1 shows that those aged 65 years and older were more likely to report protective behaviours to limit the risk of SARS-CoV-2 infection (except for reporting not leaving home) than younger participants. A similar but less marked difference was observed for those reporting living alone when asked whether they had left the home. Responses to these behavioural questions disaggregated by other demographics are considered in supplementary material section 2: those living in neighbourhoods with greater socioeconomic deprivation were more likely to report protective behaviours, although less likely to report wearing face coverings. Results were less consistent between questions and over time for region, sex, and ethnicity.

Using survey-weighted logistic regression models to estimate the independent effect of each demographic variable, we show that, compared to 18–34-year-olds, those aged 65+ were more likely to report having no contacts outside the home, with odds ratios (OR) between 1.64 (95% CI 1.55, 1.73) and 2.99 (95% CI 2.84, 3.14, P<0.05) (Figure 2). The impact of 5-17-year-olds returning to school between September and November 2020 (REACT-1 rounds 5 to 7) is also clearly visible (Figure 2); compared to 18–34-year-olds, OR were between 0.34 (95% CI 0.32, 0.35) and 0.64 (95% CI 0.61, 0.67) for having no contacts outside the home over this period. These ORs were largely unaffected by further adjustment for suspected or confirmed COVID-19 in the preceding two weeks (supplementary section 3).

**Figure 2.**
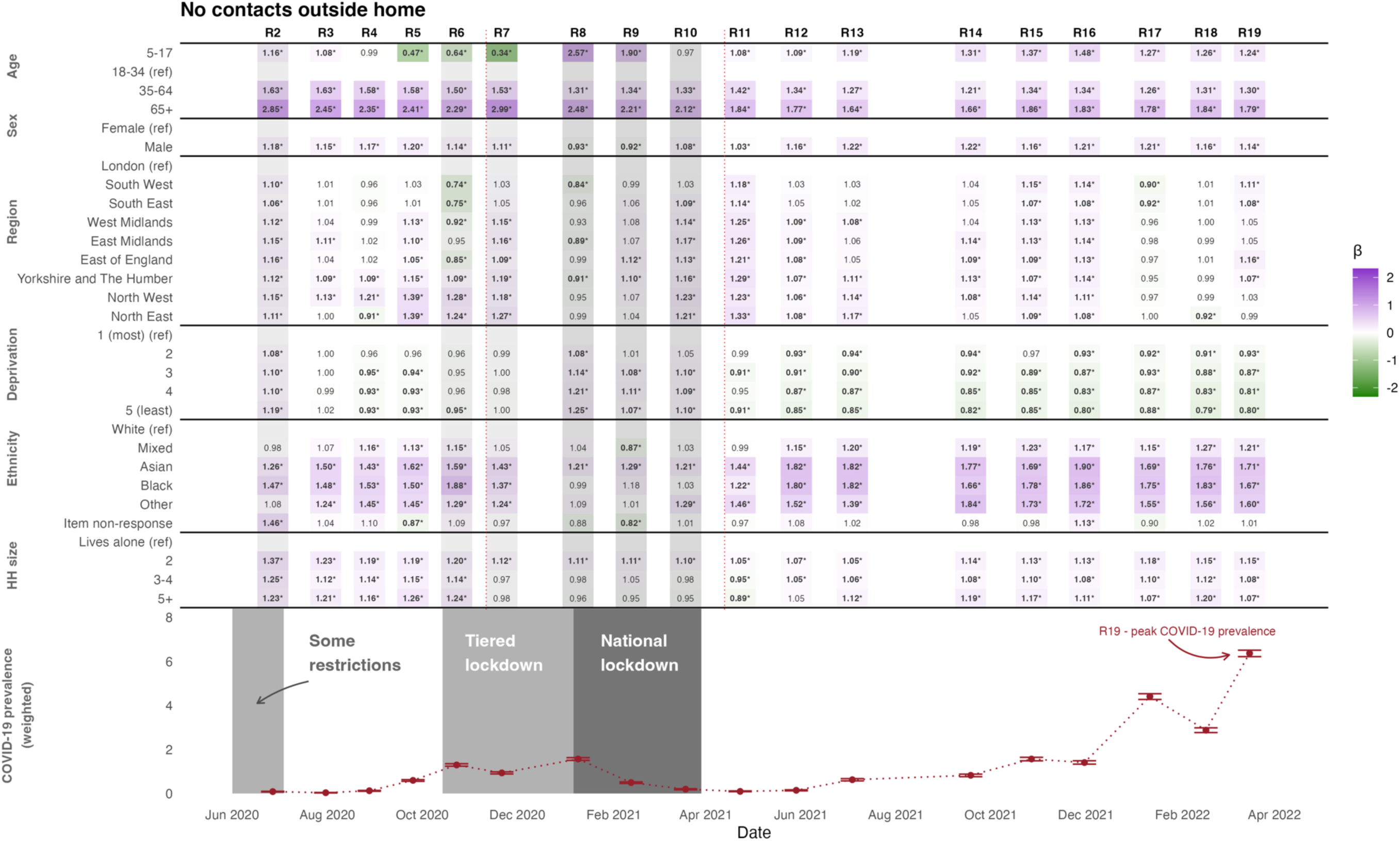
Odds ratios for whether an individual self-reports having no contacts outside the home in the preceding seven days. Purple shading indicates odds ratios greater than 1 (compared to the reference group) while green shading indicates odds ratios less than 1. Darker shading indicates estimates further from 1. Reference groups are indicated with (ref) in the row-labels. Vertical dotted red lines denote changes in question wording. Odds ratios that are statistically significantly different to 1 at P<0.05 are denoted with an asterisk and set in bold face. The bottom panel shows weighted prevalence of SARS-CoV-2 swab positivity in each round of REACT-1.

Figure 2 also suggests that men were more likely than women to report having no contacts outside the home, except in January and February 2021, while those living with other people were more likely to report having no contacts outside the home, except between November 2020 and April 2021. Results for each demographic variable on other outcomes are included in supplementary material section 3.

We also consider responses to these questions conditional on self-reported risk, vaccination status, and infection history (Figure 3). Those that considered themselves to be at risk of severe risk were more likely to report shielding, as were those that reported having been vaccinated, and those who had not had not reported previous infection. When the question was first asked, there was high self-reported use of face coverings, although over time those that did not consider themselves at severe risk and those that reported no vaccination became relatively less likely to report wearing a face covering.

**Figure 3.**
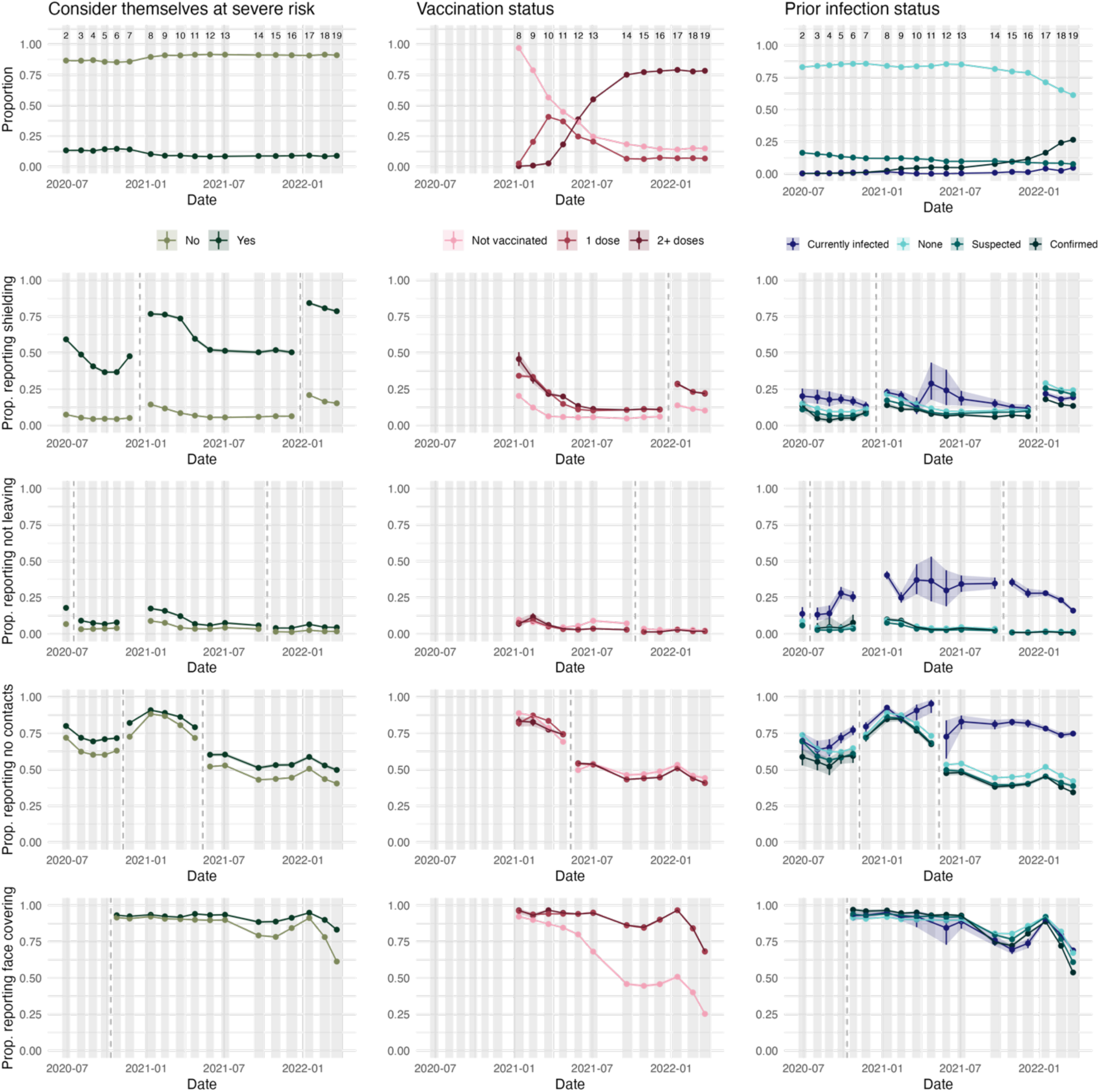
The proportion of people in England self-reporting considering themselves at severe risk, having no/one/two-plus doses of a vaccine, and having no/a suspected/a confirmed prior infection or current infection (top row). Rows 2-5 then present responses to the four focus questions, disaggregated by these responses. Vertical dashed lines denote changes in question wording. Study round numbers are provided at the top of row 1 and shaded regions indicate periods when the study was actively collecting data.

### Comparisons with mobility data

Despite the limited duration of each period considered (19 June 2020 – 11 November 2020, 4 January 2021 – 28 September 2021, and 19 October 2021 – 31 March 2022), substantial variation in mobility and behaviours was observed within each period. The random forest models of the mobility measures capture key trends while missing some of the finer day-to-day variations (Figure 4). The out-of-bag proportion of variance explained (PVE) (supplementary table S8) was high for all six mobility indices (ranging from 86% to 97%) for data from rounds 8-14 and was lower for rounds 15-19 (ranging from 68% to 86%). Additional results show that shopping was consistently the main reason for leaving the home, while the proportion of individuals reporting errands and work as a reason for leaving the home increased from January 2021 onwards (supplementary material section 2e).

**Figure 4.**
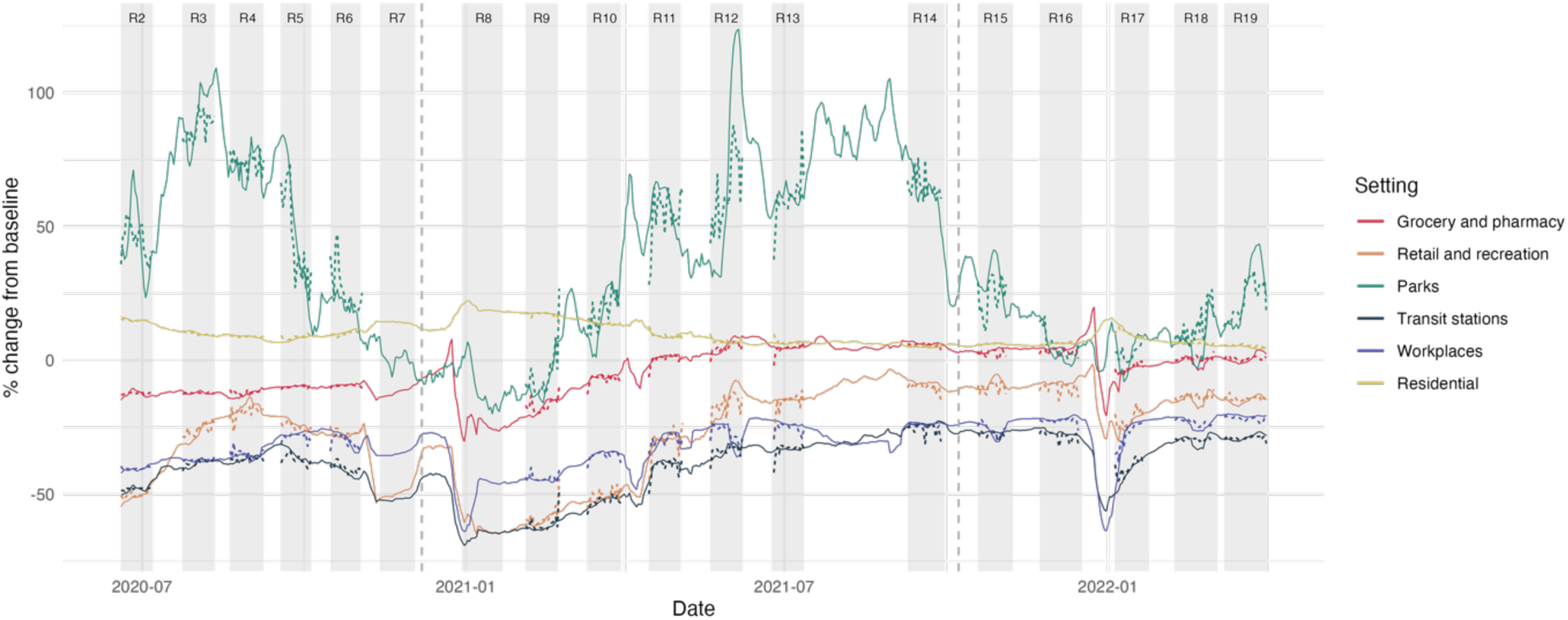
The Google mobility data (solid lines) between June 2020 and March 2022 and the random forest model predictions based on behaviours reported in REACT-1 questionnaires (dotted lines). Modelled values shown are out-of-bag predictions, so the model has not been trained on the data-point being predicted. The vertical dashed lines demark individual model fits, selected to account for substantial changes in question wording. We do not fit a model to data from study round 7 as the relevant questions were not asked.

### Comparisons with policy response indices

Results from the random forest models for the two policy response indices demonstrate a strong association between self-reported behavioural data and the stringency indices. The out-of-bag PVE was 0.98 for both models demonstrating a strong association between self-reported behaviours and aggregated measures of the stringency of public health policy.

The fitted index values based on behavioural data also identified the timing of when the change from baseline in the stringency index switched from being above to below that for the containment and health index, which occurred near the start of study round 11 (Figure 5).

**Figure 5.**
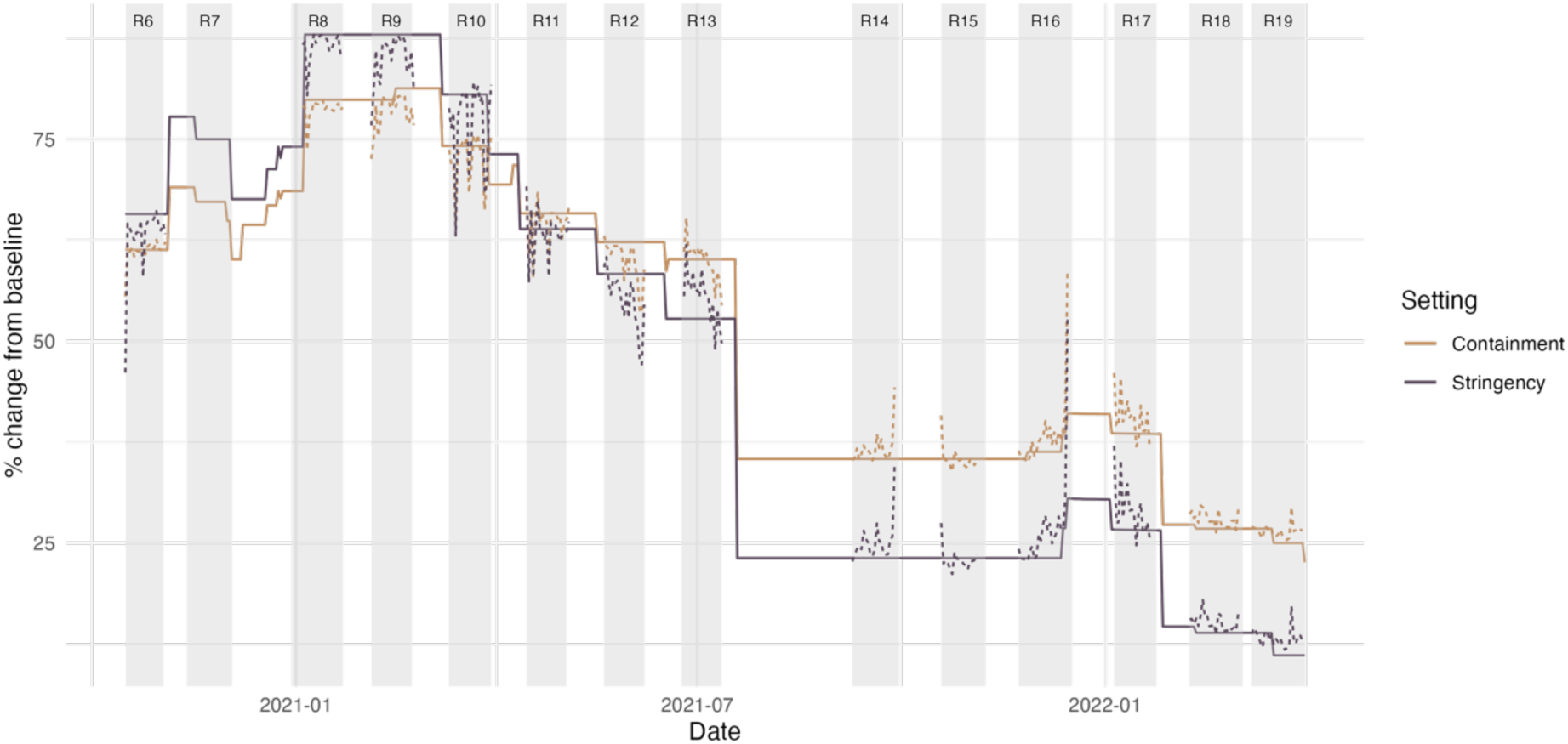
The OxCGRT stringency index and containment and health index (solid lines) between October 2020 and March 2022 and the model predictions based on behaviours reported in REACT-1 questionnaires (dotted lines). We do not fit the model to data from study round 7 as the relevant question was not asked in this round.

For comparison, we also fit a logistic regression model to the policy indices using the Google mobility data as the predictors (supplementary section 5). Responses to the four risk-related behavioural questions from the REACT-1 study explained more variance in the policy index than did the six Google mobility variates (Table S8), with an out-of-bag PVE (using the Google data) of 0.91 for the stringency index and 0.90 for the containment and health index (compared to the out-of-bag PVE of 0.99 for the model leveraging REACT-1 data), despite the Google mobility series using more covariates.

## Discussion

The COVID-19 pandemic was a major shock to health systems requiring near-unprecedented response from governments and individuals to protect individual and public health. Government response in England and many other countries included imposition of lockdowns and other containment measures, with legal restrictions on population mobility and behaviours. We show strong evidence that individuals substantially changed their behaviours over time, with behaviours to reduce risk of infection closely reflecting these policy shifts during the pandemic.

Within the period considered (19 June 2020 – 31 March 2022), protective behaviours were most pronounced in January 2021, during the winter wave of COVID-19 in England, when very few people had been vaccinated. Over the following months, vaccination rates increased while the prevalence of infection decreased, and the proportion of people that reported protective behaviours decreased.

Infection prevalence sharply increased over December 2021 and January 2022 due to the Omicron BA.1 variant (Elliott, Bodinier, et al., 2022) accompanied by an uptick in protective behaviours, although not to the previous levels seen in January 2021 during the second national lockdown. This was then followed by a second wave during February and March 2022 due to the Omicron BA.2 variant, resulting in the highest recorded SARS-CoV-2 prevalence in REACT (Elliott, Eales, et al., 2022). However, at the same time there was a fall in protective behaviours, indicating possible “pandemic fatigue”. This is particularly noticeable in the wearing of face coverings, which decreased rapidly over February and March 2022 to an all-time low, after being at relatively constant levels since face covering wearing was first recommended in July 2020.

We found important differences between demographic groups. For example, those living in larger households were generally less likely to report shielding, but were more likely to report not leaving the home. There were also large differences by age, and at times, by sex, ethnicity, and region. Understanding potential differences in behaviours by demographic groups is critical for planning and implementing non-pharmaceutical interventions (Seale *et al*., 2020).

Our results show that reported behaviours and reasons for leaving the home were strongly correlated with the OxCGRT stringency index and the OxCGRT containment and health index, and the publicly available Google mobility data, demonstrating the potential value of both the Google mobility data and the OxCGRT indices as population-level proxies for social-distancing behaviours. These are complementary to individual-based self-reported variables enabling the investigation of potential determinants of behaviour changes throughout the pandemic. However, there is no guarantee these relationships would generalise outside the time periods considered, particularly as signs of pandemic fatigue in later rounds could decrease the correlation between self-reported behaviours and the OxCGRT indices. We also emphasise that our analyses do not indicate whether policy changes are leading or lagging indicators of behavioural change, a key issue in preparing for any future pandemic.

## Limitations

Interpretation of our data is dependent on the accuracy of the self-reported behaviours. Previous studies have indicated that behaviour-related signals may be drowned out by bias and noise in self-report data (Hansen *et al*., 2021), and adherence to protective behaviours may be substantially overestimated in self-reported data both historically (Davies et al., 2022), and during the COVID-19 pandemic (Davies et al., 2023). While the strong associations between our data and aggregated community mobility data and policy indices lend some context validity to our findings, we cannot exclude biases in reporting, for example, across demographic groups. We made changes to the questionnaire over the nearly two years of the study, in some cases to reflect the evolving situation and terminology such as “shielding”, “support bubbles”, and the tier system of restrictions, which meant that participant responses over time were not always directly comparable. Additionally, the design of the study involving repeated cross-sectional sampling per round (Elliott et al., 2023) meant that the observed trends reflect changes across representative samples of the population, and in this analysis we did not investigate how an individual’s behaviour changed over time.

## Conclusion

Despite speculation that populations such as England would not be willing to change their behaviours in response to an infectious disease threat (Savage *et al*. 2020), our study, along with others (Smith et al., 2022; Wright et al., 2022) show that this is patently not the case. We show that the behavioural response closely followed the implementation of government restrictions, but these large behavioural changes may not be replicated in future events, particularly given the strong evidence of “pandemic fatigue” at the end of the study in March 2022. COVID-19 shaped and strengthened many people’s views about the effectiveness of such measures (Surano *et al*., 2021), and with multiple pandemic-potential infectious diseases being monitored (such as H5N1 (*Avian Influenza A(H5N1) – United States of America*, n.d.) and Mpox (*WHO Declares Mpox Outbreak a Public Health Emergency of International Concern*, n.d.)), understanding people’s behavioural decisions, notably though self-reported data, remains a critical component of future pandemic preparedness.

## Data Availability

Data are available online in the form of a portal. A link to this portal is provided in the preprint.

## Acknowledgements

This study was funded by the Department of Health and Social Care in England. N.S. acknowledges support from the Oxford-Radcliffe Scholarship from University College, Oxford, the EPSRC CDT in Modern Statistics and Statistical Machine Learning (Imperial College London and University of Oxford), and A. Maslov for studentship support. M.C-H. acknowledges support from Cancer Research UK, Population Research Committee Project grant “Mechanomics” (grant 22184), the H2020-EXPANSE (Horizon 2020 grant 874627) and H2020-LongITools (Horizon 2020 grant 874739). M.W. acknowledges support from the Department of Health and Social Care, England, Expanse project (Horizon 2020 grant 874627). D.A. acknowledges support from the NIHR Imperial Biomedical Research Centre. GC is an NIHR Senior Investigator and is supported by the NIHR BRC of Imperial College NHS Trust. P.E. is director of the MRC Centre for Environment and Health (MR/L01341X/1 and MR/S019669/1). P.E. acknowledges support from the Department of Health & Social Care in England (NIHR and UKRI, REACT-LC, COV-LT-0040), the Medical Research Council (MR/V030841/1), the MRC Centre for Environment and Health (MR/S019669/1), and the UK Health Security Agency for the REACT-3 study (2024-2025). C.A.D. acknowledges support from the MRC Centre for Global Infectious Disease Analysis, the NIHR Health Protection Research Unit in Emerging and Zoonotic Infections, and the NIHR funded Vaccine Efficacy Evaluation for Priority Emerging Diseases (PR-OD-1017-20007). The Oxford Martin Programme in Digital Pandemic Preparedness is acknowledged for research funding (C.A.D.).

## Notes

### Competing Interest Statement

The authors have declared no competing interest.

### Author Declarations

We obtained research ethics approval from the South Central-Berkshire B Research Ethics Committee (IRAS ID: 283787).

