## Supplementary material for "Pandemic-risk-related behaviour change in England from June 2020 to March 2022: REACT-1 study among over 2 million people"

### Supplementary Section 1: Data

#### Section 1a: Overview of survey questions

We primarily focussed on four survey questions (Table 1):

1. Are you shielding and/or taking specific precautions because you are concerned that you/your child will become severely ill with COVID-19?
2. Did you/your child leave home for any reason in the last 7 days?
3. Not including members of your household, how many different people did you have contact with yesterday? By contact we mean: any direct skin-to-skin physical contact (e.g. kiss/embrace/handshake), being less than 2 metres from another person for over 5 minutes.
4. Do you/does your child mainly wear any kind of face covering or mask when you/they are outside your/their home, because of COVID-19?

We also considered responses to additional questions:

a) In the last 7 days for what reasons have you left home?
b) Do you consider yourself/your child to be at risk of severe illness for COVID-19, for example due to an underlying health condition?
c) Do you think you have/your child has or have/has had COVID-19?
d) When were you told/did you think you/your child first had COVID-19?
e) Have you/has your child had a coronavirus vaccine?
f) If yes, how many doses (injections) have you/has your child had so far?

Variations in question wording and other nuances in interpretation are described in this section.

These questions were answered by the survey respondent (if over the age of 18), by the respondent's parent (if aged between 5 and 12), and either by the parent or by the respondent themselves (if aged between 13 and 17).

*Are you shielding and/or taking specific precautions because you are concerned that you/your child will become severely ill with COVID-19?*

Variations of this question were asked from June 2020 to March 2022 (REACT-1 rounds 2 to 19). Between June 2020 and November 2020 (REACT-1 rounds 2 to 7) the question specifically asked about shielding, then between January 2021 and December 2021 (REACT-1 rounds 8 to 16) the question was expanded to include "taking specific precautions", before the reference to shielding was removed between January 2022 and March 2022 (REACT-1 rounds 17 to 19). See Table S1 for a summary of these changes.

**Table S1** Changes to the wording of the shielding-related question in the REACT study.

| | |
|---|---|
| Jun 2020 – Nov 2020 (Rounds 2 – 7) | “Are you **shielding** because you are concerned that you/your child will become severely ill with COVID-19?” |
| Jan 2021 – Dec 2021 (Rounds 8 – 16) | “Are you **shielding** or **taking specific precautions** because you are concerned that…” |
| Jan 2022 – Mar 2022 (Rounds 17 – 19) | “Are you **taking specific precautions** because you are concerned that…” |

*Did you/your child leave home for any reason in the last 7 days, that is since <DATE/MONTH>?*

This question was first asked in June/July 2020 (REACT-1 round 2). Between July 2020 and September 2021 (REACT-1 rounds 3 to 14, except round 7) additional wording was added to clarify that short trips such as shopping or exercise should be included. This question was not asked in the November 2020 survey (REACT-1 round 7). Between October 2021 and March 2022 (REACT-1 rounds 15 to 19) this question was replaced by a list of reasons for leaving the home, with one option being “I did not leave the home”. These changes are outlined in Table S2.

**Table S2** Changes to the wording of the question asking whether respondents had left home.

| | |
|---|---|
| Jun 2020 – Jul 2020 Round 2 | “Did you/your child leave home for any reason in the last 7 days, that is since <DATE/MONTH>?” |
| Jul 2020 – Nov 2020 (Rounds 3 – 6) | “Did you/your child leave home for any reason in the last 7 days, that is since <DATE/MONTH>? Please include even short trips outside the home, e.g. for shopping, exercise.” |
| Nov 2020 – Dec 2020 (Round 7) | Question not asked |
| Jan 2021 – Sep 2021 (Rounds 8 – 14) | “Did you/your child leave home for any reason in the last 7 days, that is since <DATE/MONTH>? Please include even short trips outside the home, e.g. for shopping, exercise.” |
| Oct 2021– Mar 2022 (Rounds 15 – 19) | “In the last 7 days, for what reasons have you left home?”*<br><br>**There is an option to select “I haven’t left home”.* |

*Not including members of your household, how many different people did you have contact with yesterday? If you/they had contact with a person more than one time, please count them only once. By contact we mean: any direct skin-to-skin physical contact (e.g. kiss/embrace/handshake), being less than 2 metres from another person for over 5 minutes.*

This question was asked between June 2020 and March 2022 (REACT-1 rounds 2 to 19). Between November 2020 and April 2021 (REACT-1 rounds 7 to 11), the question wording appeared differently for those that reported being in a support or childcare bubble, beginning with: “Not including members of your household or people in your support and childcare bubbles…”. Furthermore, between November 2020 and March 2022 (REACT-1 rounds 7 to 19), those aged 17 or less were asked not to include contacts at school.

Respondents were asked to enter 0 if they had no contacts yesterday, and to give their best guess if they were not sure.

While we focussed on the proportion of people that reported having no contacts in the main paper, we also presented the mean number of self-reported contacts in supplementary section 2b.

*Do you/does your child mainly wear any kind of face covering or mask when you/they are outside your/their home, because of COVID-19?*

This question was asked between October 2020 and March 2022 (REACT-1 rounds 6 to 19). There are five options: (1) No, (2) Yes, at work/school only, (3) Yes, in other situations only (including public transport and shops), (4) Yes, usually both at work/school and in other situations, and (5) My/their face is already covered for other reasons (e.g., religious or cultural reasons). We calculate the proportion of people that responded with any yes (options 2, 3, and 4) given that the individual does not already cover their face for other reasons (option 5).

*In the last 7 days for what reasons have you left home? Please select all that apply.*

Between June 2020 and September 2021 (REACT-1 rounds 2 to 14) participants were only asked this question if they responded yes when asked if they had left the home in the past 7-days. Between October 2021 and March 2022 (REACT-1 rounds 15 to 19) all participants were asked this question. The available options varied by round – see Table S3.

**Table S3** Available options for reasons for leaving home by round. *The wording of these questions sometimes changes. **This option replaced the original question about whether or not the participant had left the home in the last 7 days, in rounds 15 onwards participants were simply presented with this list.

| Reason | 2 | 3 | 4 | 5 | 6 | 7 | 8 | 9 | 10 | 11 | 12 | 13 | 14 | 15 | 16 | 17 | 18 | 19 |
|---|---|---|---|---|---|---|---|---|---|---|---|---|---|---|---|---|---|---|
| For work | Y | Y | Y | Y | Y | | Y | Y | Y | Y | Y | Y | Y | Y | Y | Y | Y | Y |
| To volunteer | Y | Y | Y | Y | Y | | Y | Y | Y | Y | Y | Y | Y | Y | Y | Y | Y | Y |
| For medical or dental appointments | Y | Y | Y | Y | Y | | Y | Y | Y | Y | Y | Y | Y | Y | Y | Y | Y | Y |
| To care for someone else* | Y | Y | Y | Y | Y | | Y | Y | Y | Y | Y | Y | Y | Y | Y | Y | Y | Y |
| To socialise with people in a public place | Y | Y | Y | Y | Y | | Y | | | | | | | | | | | |
| To socialise with people in a personal place | Y | Y | Y | Y | Y | | Y | | | | | | | | | | | |
| To meet with someone outside | | | | | | | | Y | Y | Y | | | | | | | | |
| To meet with people in your childcare bubble | | | | | | | | Y | Y | Y | | | | | | | | |
| To meet with people in your support bubble | | | | | | | | | Y | Y | | | | | | | | |
| To socialise with people outside | | | | | | | | | | | Y | Y | Y | Y | Y | Y | Y | Y |
| To socialise with people inside | | | | | | | | | | | Y | Y | Y | Y | Y | Y | Y | Y |
| For outdoor exercise | Y | Y | Y | Y | Y | | Y | Y | Y | Y | Y | Y | Y | Y | Y | Y | Y | Y |
| To go shopping | Y | Y | Y | Y | Y | | Y | Y | Y | Y | Y | Y | Y | Y | Y | Y | Y | Y |
| For errands | Y | Y | Y | Y | Y | | Y | Y | Y | Y | Y | Y | Y | Y | Y | Y | Y | Y |
| To take a child to school or childcare | | | | | | | | Y | Y | Y | Y | Y | Y | Y | Y | Y | Y | Y |
| To get a vaccination | | | | | | | | | Y | Y | Y | Y | Y | Y | Y | Y | Y | Y |
| To walk a dog/other pet care | | | | | | | | | Y | Y | Y | Y | Y | Y | Y | Y | Y | Y |
| To go to school/college/university | | | | | | | | | | Y | Y | Y | Y | Y | Y | Y | Y | Y |
| To go on holiday (in the UK or abroad) | | | | | | | | | | | Y | Y | Y | Y | Y | Y | Y | Y |
| Other reasons | Y | Y | Y | Y | Y | | Y | Y | Y | Y | Y | Y | Y | Y | Y | Y | Y | Y |
| I have not left home in the past 7 days** | | | | | | | | | | | | | | Y | Y | Y | Y | Y |

*Do you consider yourself/your child to be at risk of severe illness for COVID-19, for example due to an underlying health condition?*

A variation of this question was asked throughout the study. Table S4 shows changes to question wording and available answers.

**Table S4** Question wording and available answers for the question relating to whether an individual believes they are at risk of severe illness for COVID-19.

| **Period** | **Question** | **Available answers** |
|---|---|---|
| *Round 1*<br>*(May 2020)* | *"Have you/has your child been contacted by letter or text message to say you/they are at severe risk from COVID-19 due to an underlying health condition and should be shielding?"* | 1. Yes<br>2. No |
| Round 2 to 7<br>(June 2020 – November 2020) | "Do you consider yourself/your child to be at risk for severe illness for COVID-19, for example due to an underlying health condition?" | |
| Round 8 to 16<br>(January 2021 – December 2021) | "Do you consider yourself/your child to be at risk for severe illness for COVID-19, for example due to an underlying health condition **or because you/they are clinically extremely vulnerable?**" | 1. Yes<br>2. No<br>3. Don't know |
| Round 17 to 19<br>(January 2022 – March 2022) | "Do you consider yourself/your child to be at risk for severe illness for COVID-19, for example due to an underlying health condition?" | |

*Do you think that you have/your child has or have/has had COVID-19?*

This question was asked in all rounds of the study. The wording did not change. Four answers were available:

1. Yes, confirmed by a positive test
2. Yes, suspected by a doctor but not tested
3. Yes, my own suspicions
4. No

If the individual responded with (1), (2), or (3), they were then asked*: "When were you told/did you think you/your child first had COVID-19? If you are not sure, please give an estimate."* We use responses to this question to derive three variables:

- Reports a current suspected/confirmed infection: when the individual responds with option (1), (2), or (3), and their estimated date of first infection is within 13 days prior to completing the questionnaire.
- Reports a previous suspected/confirmed infection: when the individual responds with option (1), (2), or (3), and their estimated date of first infection is at least 14 days prior to completing the questionnaire. We also include those who do not report an estimated date of first infection in this group.

- Reports a previous confirmed infection: when the individual responds with option (1) and their estimated date of first infection is at least 14 days prior to completing the questionnaire. We also include those who do not report an estimated date of first infection in this group.

*Vaccination-related questions*

In round 8, all vaccination-related questions were asked during the follow-up survey (the survey featuring behavioural questions, typically completed when self-administering the swab, as opposed to the short registration survey). These questions were:

1. Have you/has your child had a coronavirus vaccine? (Yes, No, Don't know)
2. If yes, how many doses (injections) have you/has your child had so far? (One, Two, More than two)

In round 9 onwards, the above questions were asked during the registration survey (the survey asked when signing up to participate in the study). The follow-up survey then asked:

1. We asked you this at when you registered to receive a swab test kit but we would just like to double check, have you/has your child ever had a coronavirus vaccine? (Yes, No, Don't know)
2. If yes, can we just check, have you/has your child had a vaccination since you/they registered to take part in this study? (Yes, No, Don't know)
3. If yes, how many doses (injections) have you/has your child had so far? (One, Two, More than two)

Between rounds 8 and 13, these questions were asked of all respondents (or parents of respondents) aged 16 and over. From round 14 onwards these questions were asked of all respondents (or parents of respondents) aged 12 and over. From rounds 15 onwards, questions asking about the number of doses received also included the options for "Three" and "More than three".

When processing the data, we treat anyone that reports at least one dose in either the registration survey or follow-up survey as being "vaccinated", and anyone that reports at least two doses in either survey as being "double vaccinated".

#### SECTION 1B: NON-RESPONSE RATES AND INCLUSION/EXCLUSION CRITERIA

There are a total of 3,401,391 participants listed across the 19 rounds of REACT-1. Of these, 1023 did not have valid survey weights, these are removed prior to performing any analysis, leaving a potential sample size of 3,400,368.

When considering behavioural questions, we do not explicitly exclude those with invalid swab results, although we omit round 1 of the study from our analysis as only a small subset of behavioural questions were asked. Of the 3,238,601 participants with valid weights in study rounds 2-to-19, a total of 2,177,657 participants at least partially responded to the follow-up survey. Individuals were not required to answer all questions in the follow-up survey, so sample sizes for individual questions may be smaller than this total. Missing responses to individual questions were ignored in the analysis. Sample sizes by study round are provided in Table S5.

When considering behavioural variables on a daily basis, the date the survey was last accessed was used. If this was missing, the date the swab was taken was used. If both dates were missing, and the individual is still reported as responding to the follow-up survey, then their records are ignored. There are 10,502 individuals for which the date is missing while they still responded to the follow-up survey, and this only occurred in round 12 (n = 5559) and round 13 (n = 4943).

Comparisons with mobility and stringency data leverage daily aggregations of individual responses. In these cases, we only consider days on which at least 200 individuals responded to the survey.

Table S5. Total participants and number of participants with and without valid survey weights by round. Also, the number of participants that returned a valid RT-PCR swab, the number with a valid survey date, the number that (at least partially) completed the follow-up survey and had a valid survey date, and the number that satisfy all three criteria. Percentages in parentheses are of those with valid survey weights, as we filter out invalid weights before performing any analysis.

| R | Total | Invalid weights | Valid weights | Valid swabs | Valid survey date | Responded to survey & valid survey date | All valid |
|---|---|---|---|---|---|---|---|
| 2 | 219862 | 0 | 219862 | 159199 (72%) | 171591 (78%) | 145753 (66%) | 132790 (60%) |
| 3 | 225729 | 0 | 225729 | 162822 (72%) | 171869 (76%) | 144638 (64%) | 135742 (60%) |
| 4 | 211340 | 0 | 211340 | 154406 (73%) | 162494 (77%) | 137406 (65%) | 129411 (61%) |
| 5 | 232190 | 0 | 232190 | 174948 (75%) | 182694 (79%) | 152007 (65%) | 144026 (62%) |
| 6 | 211285 | 0 | 211285 | 160173 (76%) | 168089 (80%) | 140967 (67%) | 133315 (63%) |
| 7 | 212891 | 2 | 212889 | 168181 (79%) | 176050 (83%) | 148656 (70%) | 141011 (66%) |
| 8 | 214751 | 1015 | 213736 | 167625 (78%) | 175596 (82%) | 154060 (72%) | 145059 (68%) |
| 9 | 210000 | 0 | 210000 | 165456 (79%) | 172807 (82%) | 150315 (72%) | 143118 (68%) |
| 10 | 180037 | 0 | 180037 | 140844 (78%) | 148698 (83%) | 129689 (72%) | 122022 (68%) |
| 11 | 170715 | 0 | 170715 | 127407 (75%) | 134783 (79%) | 118398 (69%) | 111187 (65%) |
| 12 | 161421 | 0 | 161421 | 108911 (67%) | 110920 (69%) | 93716 (58%) | 91812 (57%) |
| 13 | 146460 | 1 | 146459 | 98231 (67%) | 100230 (68%) | 85018 (58%) | 83124 (57%) |
| 14 | 146860 | 0 | 146860 | 100527 (68%) | 108008 (74%) | 93281 (64%) | 86088 (59%) |
| 15 | 142905 | 0 | 142905 | 100112 (70%) | 107976 (76%) | 95791 (67%) | 88504 (62%) |
| 16 | 130351 | 0 | 130351 | 97089 (74%) | 101493 (78%) | 88903 (68%) | 83866 (64%) |
| 17 | 139789 | 0 | 139789 | 102174 (73%) | 108764 (78%) | 98579 (71%) | 92478 (66%) |
| 18 | 134252 | 1 | 134251 | 94950 (71%) | 99985 (74%) | 87104 (65%) | 82416 (61%) |
| 19 | 148782 | 0 | 148782 | 109181 (73%) | 115177 (77%) | 102850 (69%) | 97275 (65%) |

Table S6. Total valid responses (individuals who responded to the follow-up survey and have a valid survey response date), the number of individuals with missing responses to each of four specific questions, and the number of individuals with responses to all four questions. Percentages in parentheses are of the “total valid” column.

| R | Total valid | Is shielding | Didn’t leave home | No contacts outside home | Wears a face covering | Total valid (for all four) |
|---|---|---|---|---|---|---|
| 2 | 145753 | 2690 (1.8%) | 2348 (1.6%) | 5720 (3.9%) | - | 138931 (95.3%) |
| 3 | 144638 | 1507 (1.0%) | 1711 (1.2%) | 4831 (3.3%) | - | 139807 (96.7%) |
| 4 | 137406 | 1357 (1.0%) | 1564 (1.1%) | 4800 (3.5%) | - | 132603 (96.5%) |
| 5 | 152007 | 1609 (1.1%) | 1856 (1.2%) | 5292 (3.5%) | - | 146714 (96.5%) |
| 6 | 140967 | 133 (0.1%) | 362 (0.3%) | 2484 (1.8%) | 894 (0.6%) | 138476 (98.2%) |
| 7 | 148656 | 1760 (1.2%) | - | 8894 (6.0%) | 2105 (1.4%) | 139762 (94.0%) |
| 8 | 154060 | 1135 (0.7%) | 1201 (0.8%) | 6793 (4.4%) | 1330 (0.9%) | 147266 (95.6%) |
| 9 | 150315 | 1109 (0.7%) | 1179 (0.8%) | 6478 (4.3%) | 1343 (0.9%) | 143836 (95.7%) |
| 10 | 129689 | 1134 (0.9%) | 1233 (1%) | 6617 (5.1%) | 1516 (1.2%) | 123068 (94.9%) |
| 11 | 118398 | 882 (0.7%) | 950 (0.8%) | 5302 (4.5%) | 1182 (1.0%) | 113093 (95.5%) |
| 12 | 93716 | 611 (0.7%) | 662 (0.7%) | 2112 (2.3%) | 889 (0.9%) | 91602 (97.7%) |
| 13 | 85018 | 573 (0.7%) | 613 (0.7%) | 1832 (2.2%) | 828 (1.0%) | 83185 (97.8%) |
| 14 | 93281 | 711 (0.8%) | 763 (0.8%) | 2171 (2.3%) | 1107 (1.2%) | 91110 (97.7%) |
| 15 | 95791 | 622 (0.6%) | 710 (0.7%) | 2793 (2.9%) | 803 (0.8%) | 92998 (97.1%) |
| 16 | 88903 | 582 (0.7%) | 676 (0.8%) | 2475 (2.8%) | 749 (0.8%) | 86427 (97.2%) |
| 17 | 98579 | 681 (0.7%) | 764 (0.8%) | 2664 (2.7%) | 829 (0.8%) | 95915 (97.3%) |
| 18 | 87104 | 566 (0.6%) | 643 (0.7%) | 2549 (2.9%) | 725 (0.8%) | 84553 (97.1%) |
| 19 | 102850 | 626 (0.6%) | 717 (0.7%) | 2972 (2.9%) | 806 (0.8%) | 99877 (97.1%) |

### Supplementary Section 2: Additional descriptive results

#### Section 2a. Additional demographic stratification

In the main paper we consider responses to the four key risk-related behavioural questions (table 1) by age-group and household size. In this section we consider responses to these four questions by additional demographic disaggregations: region, deprivation quintile of neighbourhood, ethnicity, and sex (figures S1a-S1d).

For visual clarity, we present results by region using four regions. These are created by aggregating the standard nine English regions as follows: “North” includes North West, North East, and Yorkshire and The Humber; “Midlands and East” includes West Midlands, East Midlands, East of England), “London” (London only), and “South England” including South West and South East).

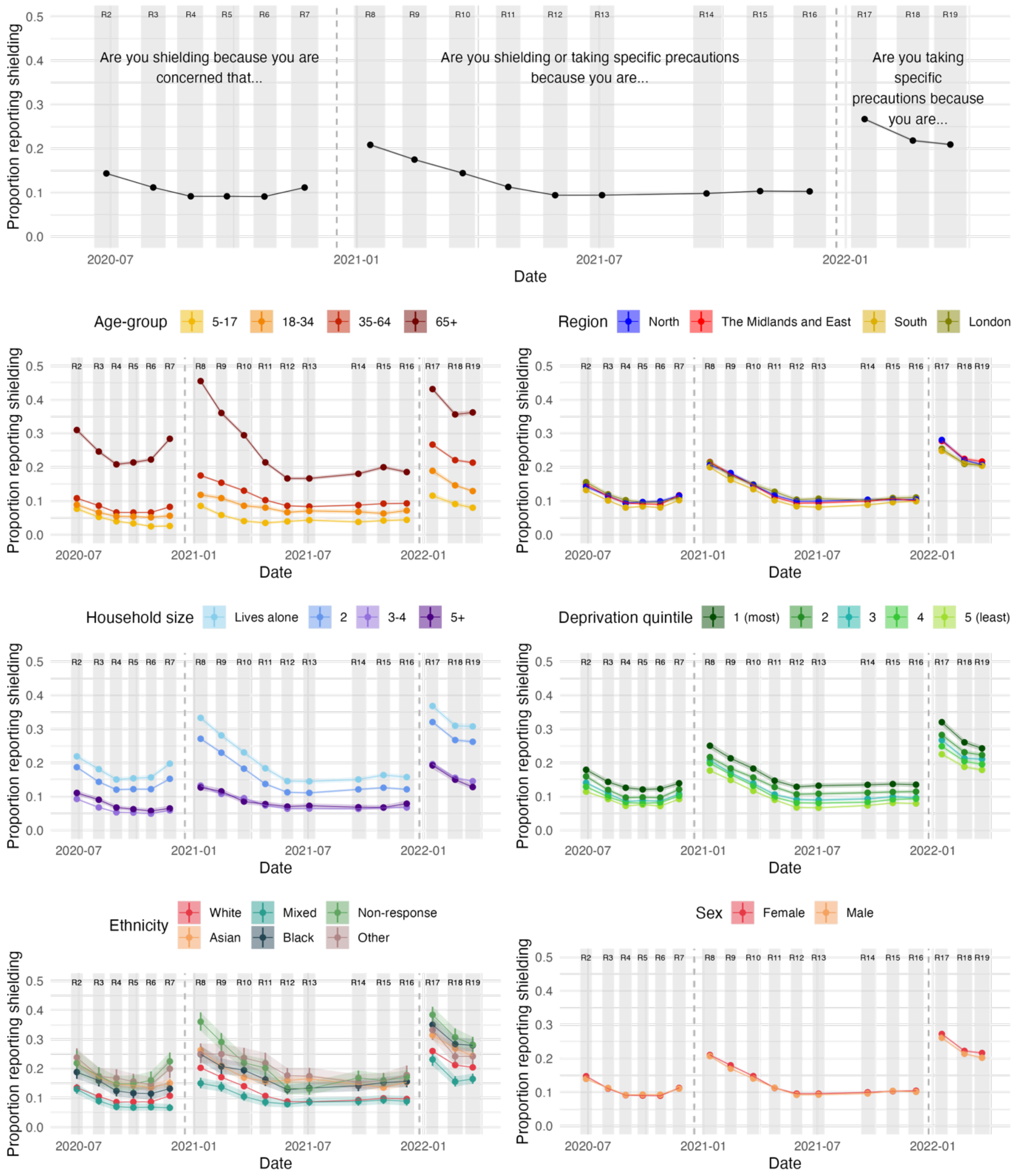

**Figure S1a** The proportion of people that report shielding and/or taking specific precautions because they consider themselves "to be at risk of severe illness for COVID-19". The question wording changes between REACT-1 rounds 7 and 8, and again between REACT-1 rounds 16 and 17 – this is marked by a dashed grey line. Shaded areas and vertical lines show 95% confidence intervals about the estimated proportion. Individual points are joined by lines and shading for clarity, but this should not be interpreted as an interpolation.

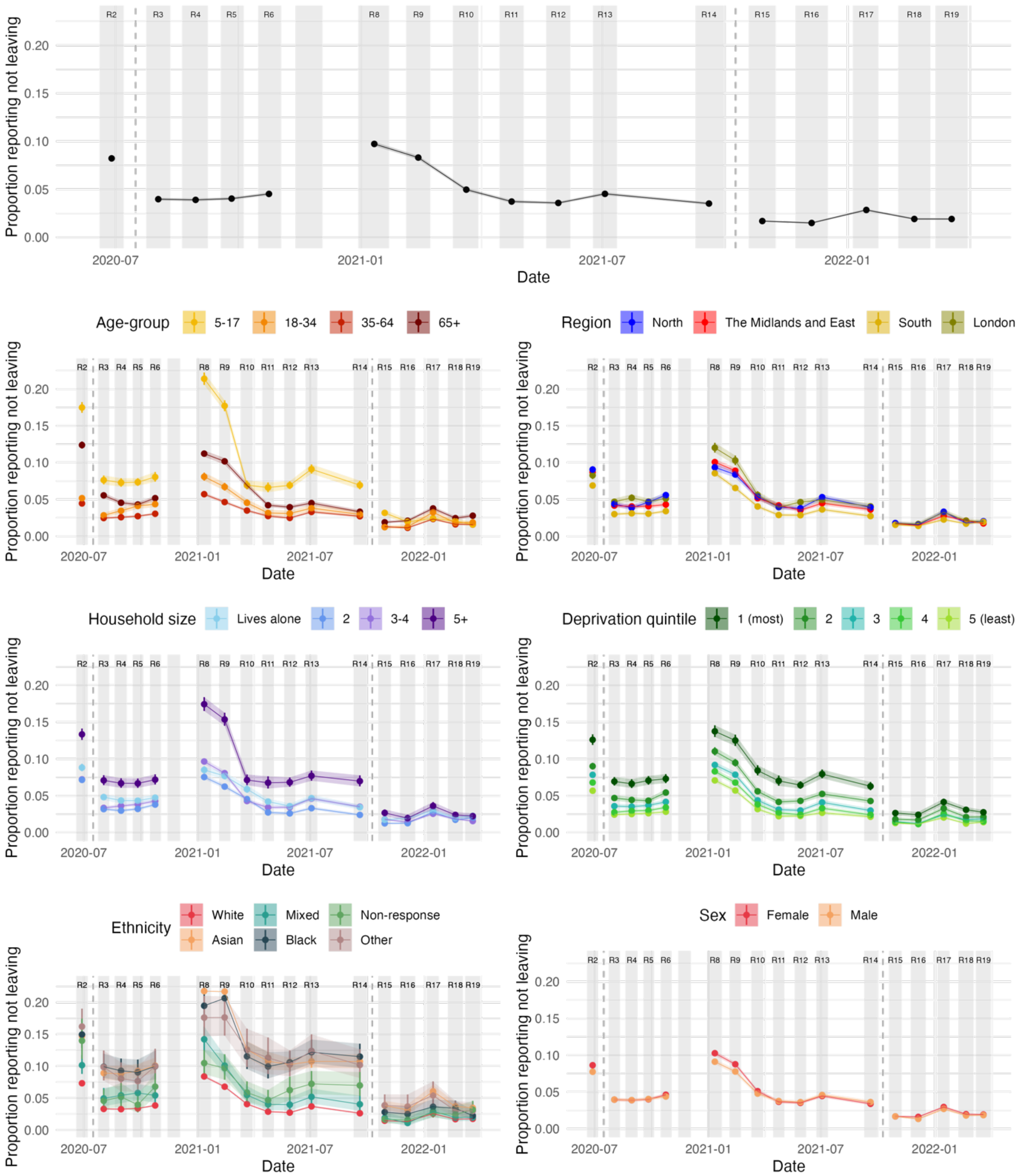

**Figure S1b** The proportion of people that report not leaving the home in 7 days prior to completing the questionnaire. The question wording changes between REACT-1 rounds 2 and 3, and again between REACT-1 rounds 14 and 15 – this is marked by a dashed grey line. This question was not asking in round 7. Shaded areas and vertical lines show 95% confidence intervals about the estimated proportion. Individual points are joined by lines and shading for clarity, but this should not be interpreted as an interpolation.

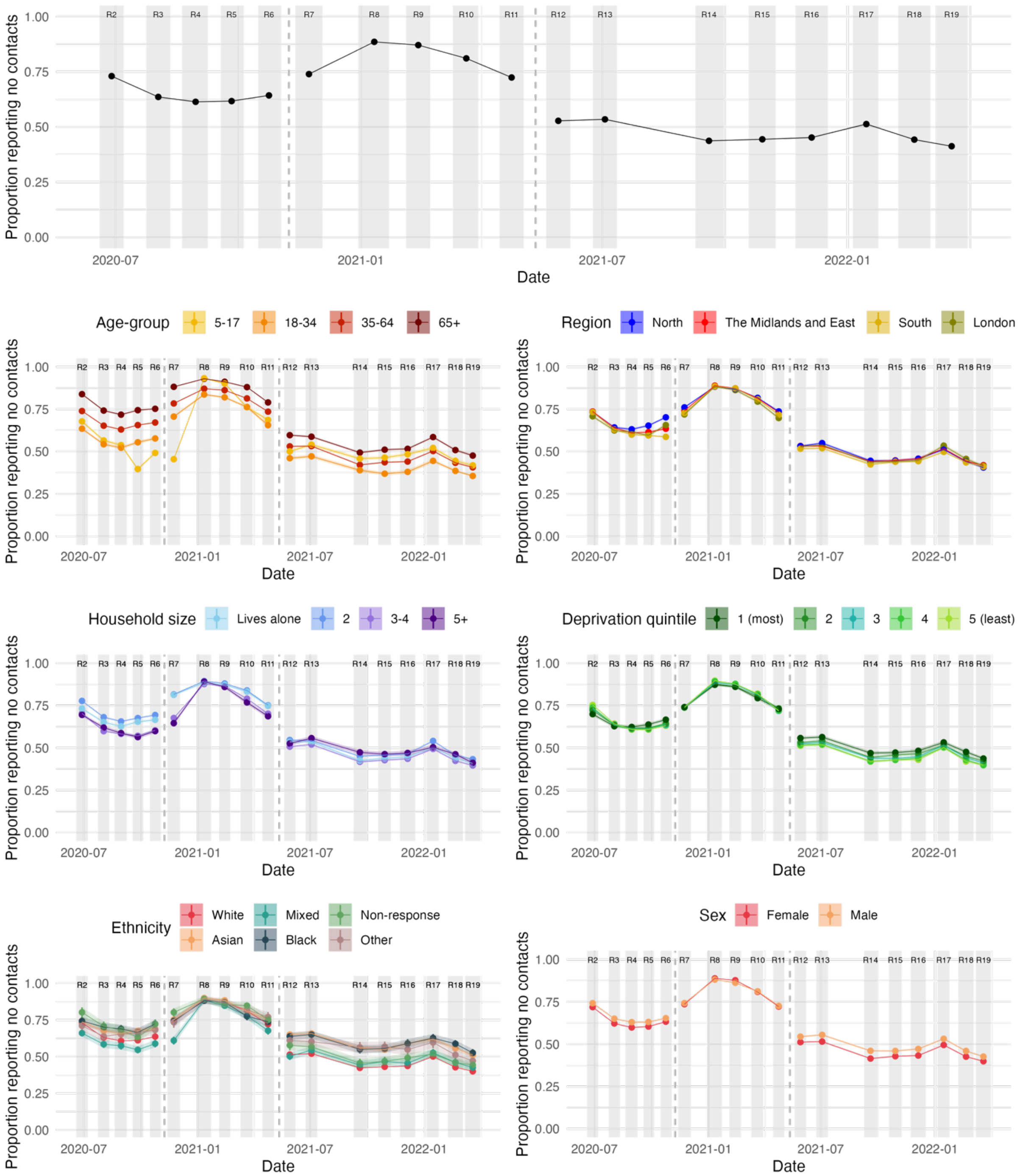

**Figure S1c** The proportion of people that report having no contacts outside the home on the day preceding the questionnaire. The question wording changes between REACT-1 rounds 6 and 7, and again between REACT-1 rounds 11 and 12 – this is marked by a dashed grey line. Shaded areas and vertical lines show 95% confidence intervals about the estimated proportion. The mean number of contacts are also presented in supplementary Figure S2. Individual points are joined by lines and shading for clarity, but this should not be interpreted as an interpolation.

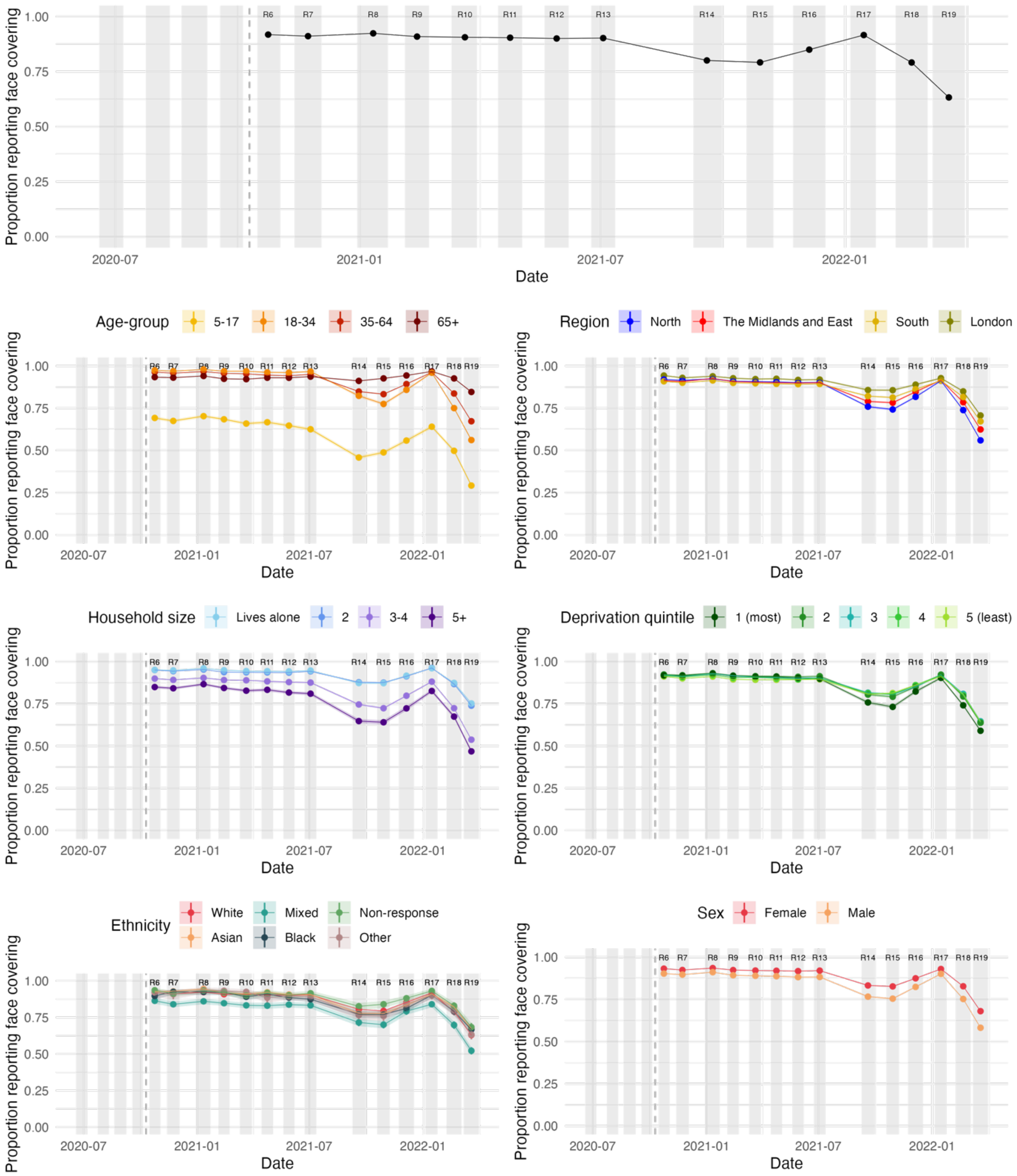

**Figure S1d** The proportion of people that report wearing a face covering at least sometimes when leaving the home. This question asked in REACT-1 round 6 onwards. Shaded areas and vertical lines show 95% confidence intervals about the estimated proportion. Individual points are joined by lines and shading for clarity, but this should not be interpreted as an interpolation.

#### SECTION 2B: MEAN CONTACTS

In the main results we present the proportion of people that would report having no contacts on the day prior to answering the survey. However, the question as outlined in supplementary section 1 asks about the number of contacts, allowing us to additionally consider the mean number of contacts that people would report. We use the *svymean()* function from the *survey* package in R to calculate the survey mean and 95% confidence intervals. These are reported for the standard demographic stratifications in Figure S2.

To prevent rare extremely high reported total contacts from biasing the results (over the study, a total of 350 individuals reported at least 500 contacts, 137 individuals reported at least 1,000 contacts, and 17 reported at least 10,000 contacts), we remove responses with at least 1,000 reported contacts.

Despite removing individual outliers in these data, supplementary figure 2 highlights high mean total contacts in 5-17-year-olds in rounds 5, 6, and 7, coinciding with the re-opening of schools in September 2020. These high means were not repeated in the same period in 2021 (study rounds 14 and 15), suggesting that parents were more likely to report higher rates of "close contacts" for their children in 2020 than 2021.

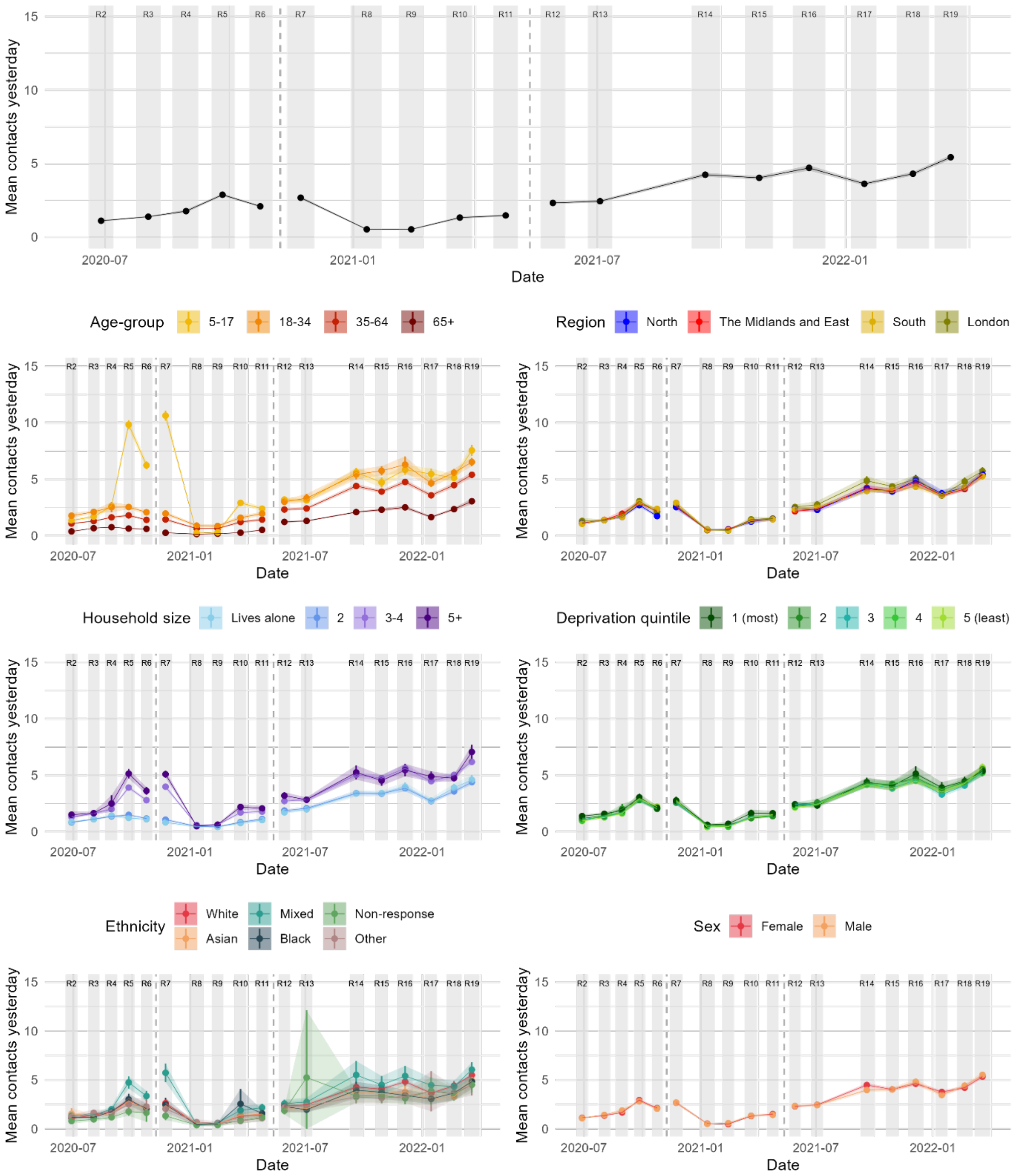

**Figure S2** The mean number of contacts that people would report having on the day preceding the questionnaire. The question wording changes between REACT-1 rounds 6 and 7, and again between REACT-1 rounds 11 and 12 – this is marked by a dashed grey line. Shaded areas and vertical lines show 95% confidence intervals about the estimated mean.

#### SECTION 2C: RISK BELIEFS, VACCINATION, AND INFECTION

The proportion of people that would report believing they are at risk of *severe* illness for COVID-19 remained relatively stable throughout the pandemic (Figure S3). A decrease was observed between November 2020 and January 2021 (REACT-1 rounds 7 and 8), however this corresponded with the addition of “Don’t know” as an answer to the question. Older people, those living in neighbourhoods with greater levels of socioeconomic deprivation, and those living alone were more likely to report believing they are at risk of severe illness.

The proportion of people that reported a suspected or confirmed past infection fell from 16.7% in May 2020 (REACT-1 round 2) to 13.3% in November 2020 (REACT-1 round 7), while the proportion of people that reported a confirmed past infection (by positive test) increased from 0.3% to 1.6% over the same time period (Figures S5 and S6).

Figures S7 and S8 show a distinct decrease in the proportion of 18–34-year-olds that report being vaccinated between January 2022 (REACT-1 round 17) and February 202 (REACT-1 round 18). In fact, between these study rounds, the proportion of 18–34-year-olds that would report not being vaccinated increased from 5.1% (95% confidence interval 4.8% to 5.6%) to 9.1% (8.8% to 9.4%). The proportion of this group that would report not being double vaccinated increased from 7.9% (7.4% to 8.3%) to 14.0% (13.6% to 14.4%).

*The proportion of people in England that would report believing they are at risk of severe illness for COVID-19*

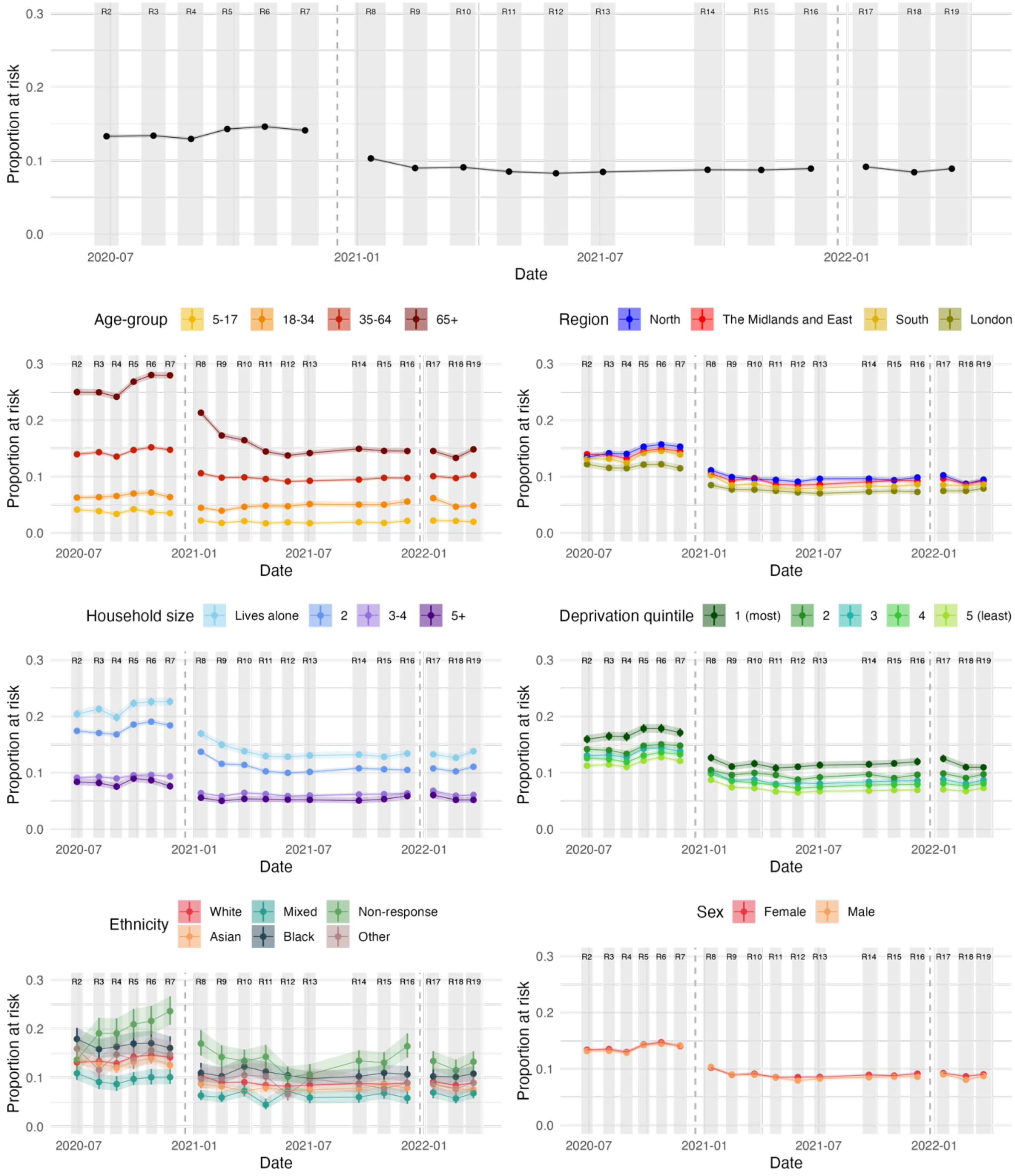

**Figure S3** The proportion of people in England that would report believing they are "at risk of severe illness for COVID-19". The question wording changes between rounds 7 and 8, and again between rounds 16 and 17 – these changes are marked by vertical dashed lines. The list of possible answers also changes between rounds 7 and 8, with the addition of a "Don't know" option. A detailed description of these changes is given in supplementary Table S4. We report the proportion of people that would respond "Yes" to this question. Shaded areas and vertical lines show 95% confidence intervals about the estimated proportion.

*The proportion of people in England that would report recent infection (suspected or confirmed)*

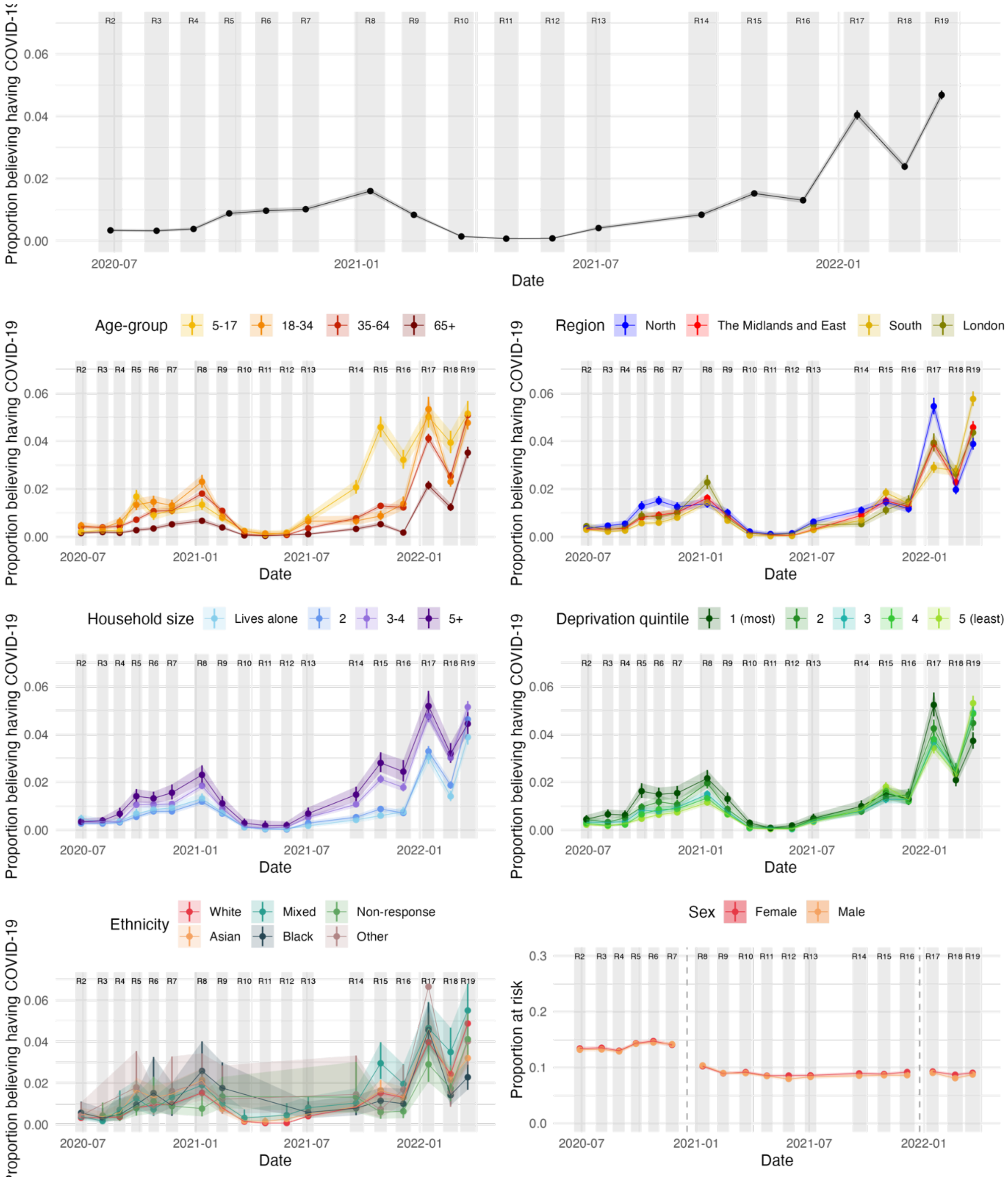

**Figure S4** The proportion of people in England that would report a recent infection (first suspected/tested positive within the preceding 13 days), either confirmed by a positive test, suspected by a doctor, or by their own suspicions. Shaded areas and vertical lines show 95% confidence intervals about the estimated proportion.

*The proportion of people in England that would report past infection (suspected or confirmed)*

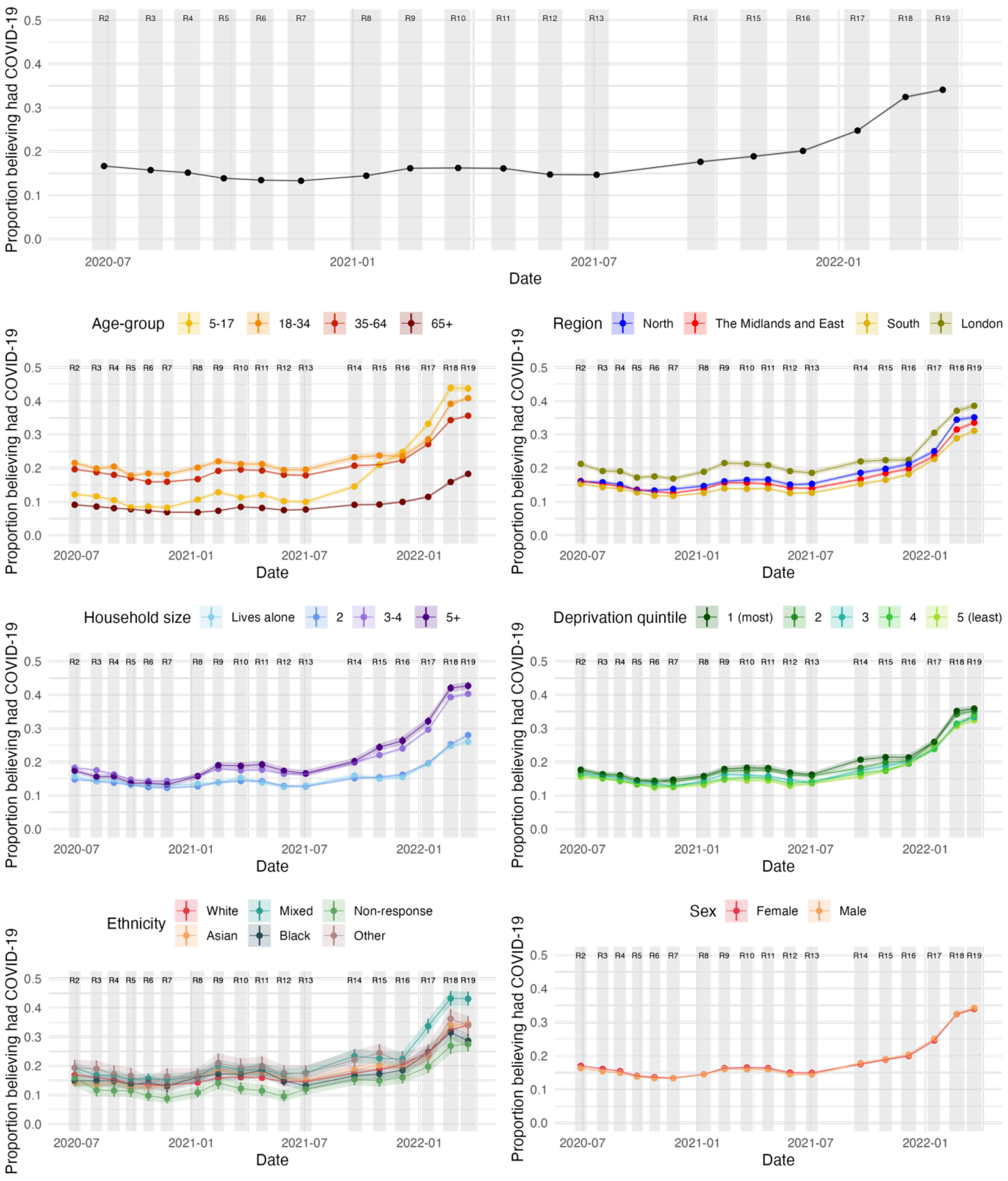

**Figure S5** The proportion of people in England that would report a past infection (first suspected/tested positive at least 14 days ago), either confirmed by a positive test, suspected by a doctor, or by their own suspicions. Shaded areas and vertical lines show 95% confidence intervals about the estimated proportion.

*The proportion of people in England that report past infection (confirmed only)*

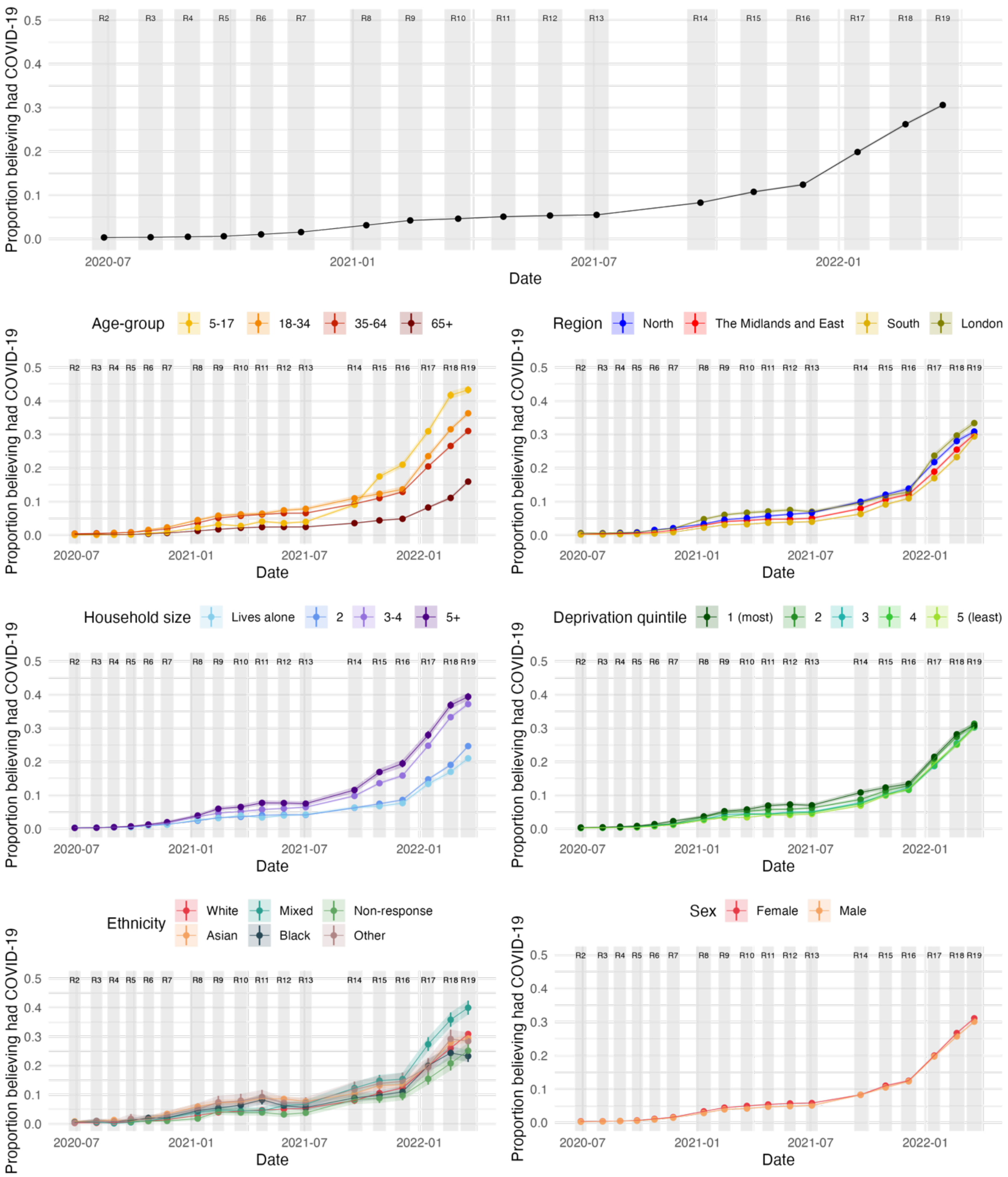

**Figure S6** The proportion of people in England that would report a past infection confirmed by a positive test, at least 14 days before completing the questionnaire. Shaded areas and vertical lines show 95% confidence intervals about the estimated proportion.

*The proportion of people in England that would report being vaccinated with at least one dose*

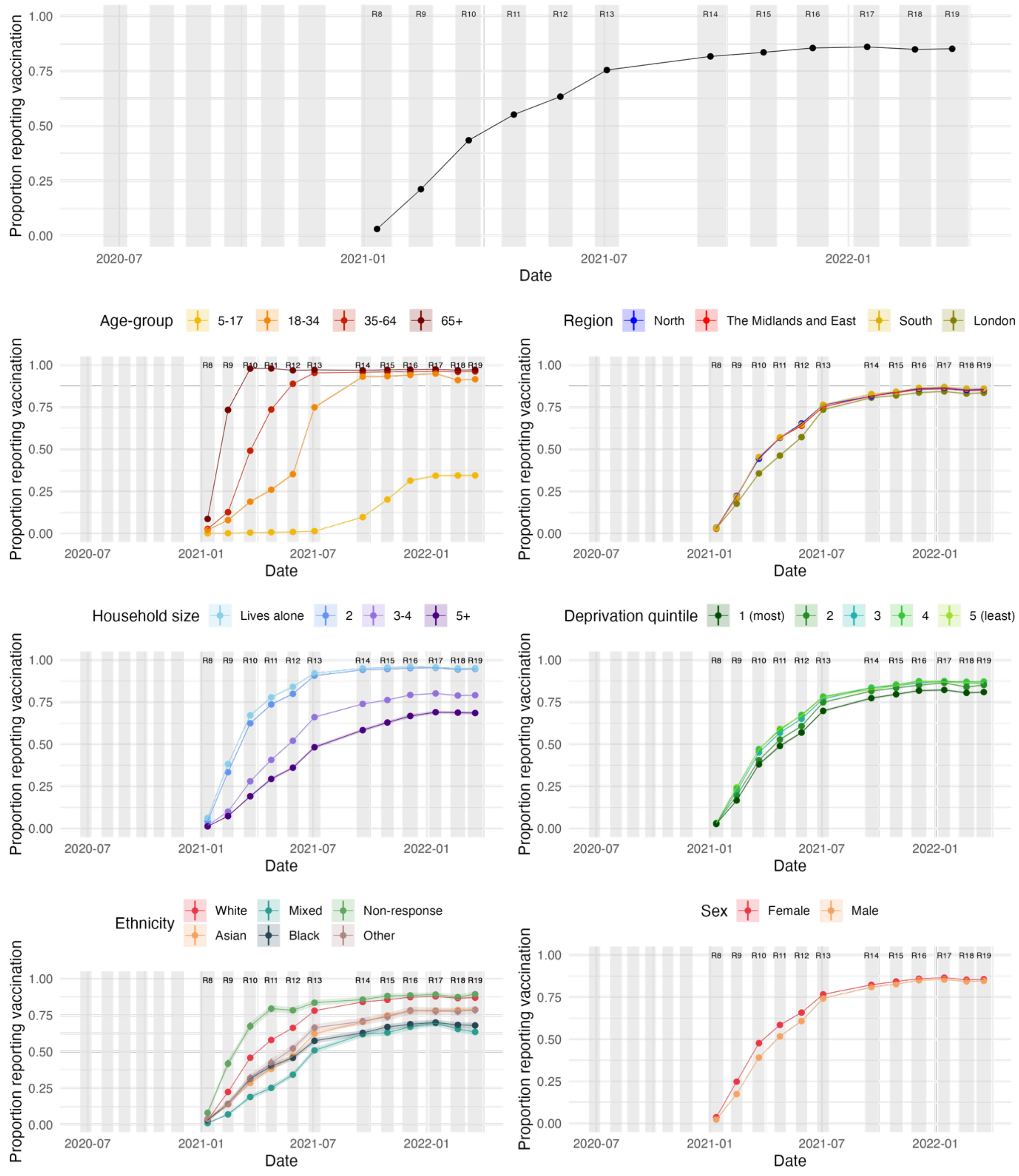

**Figure S7** The proportion of people in England that would report having received at least one dose of a COVID-19 vaccine. Shaded regions and vertical lines show 95% confidence intervals about the estimated proportions.

*The proportion of people in England that would report being vaccinated with at least two doses*

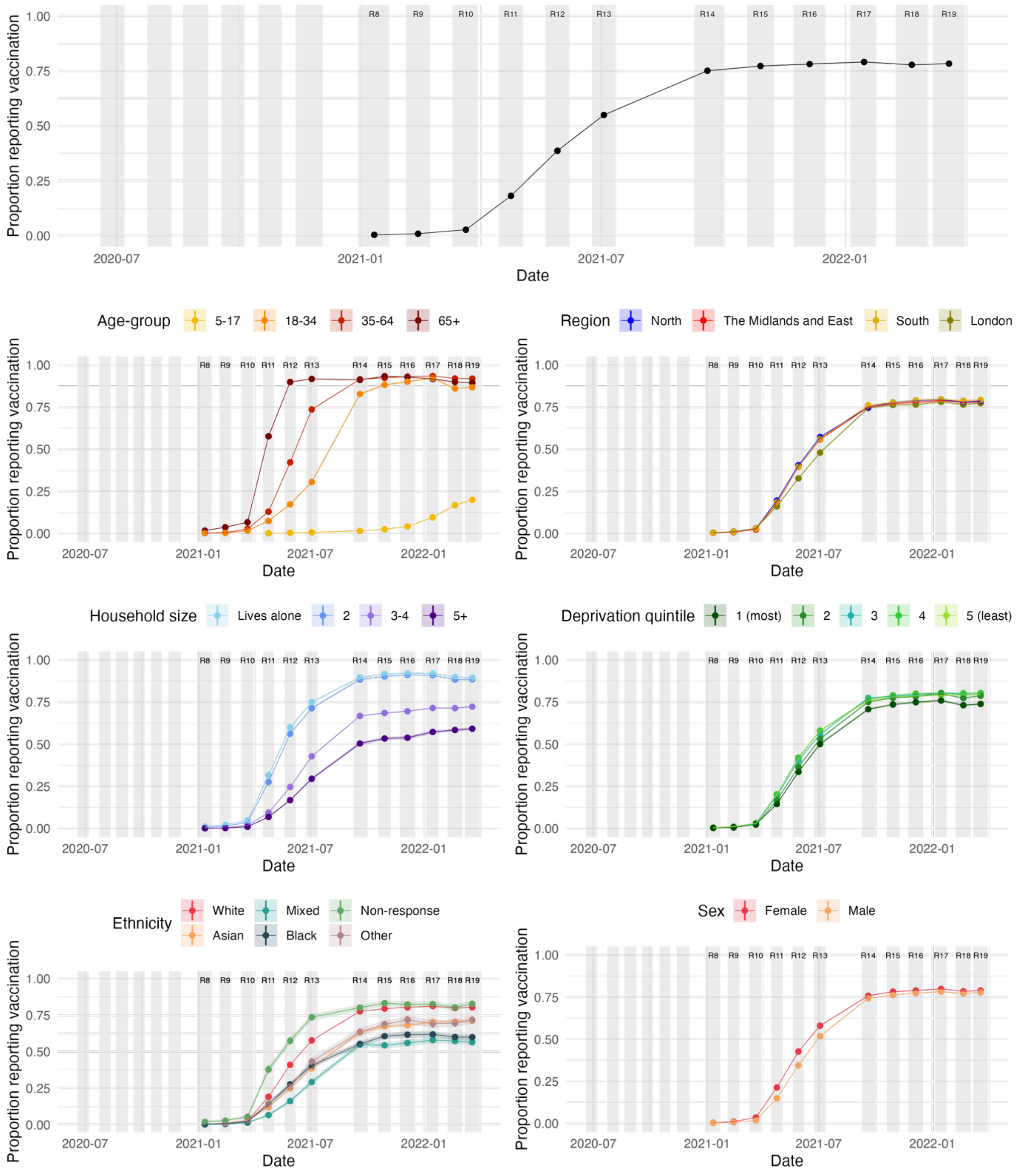

**Figure S8** The proportion of people in England that would report having received at least two doses of a COVID-19 vaccine. Shaded regions and vertical lines show 95% confidence intervals about the estimated proportions.

#### SECTION 2D: BEHAVIOURS STRATIFIED BY RISK BELIEFS, VACCINATION, AND INFECTION

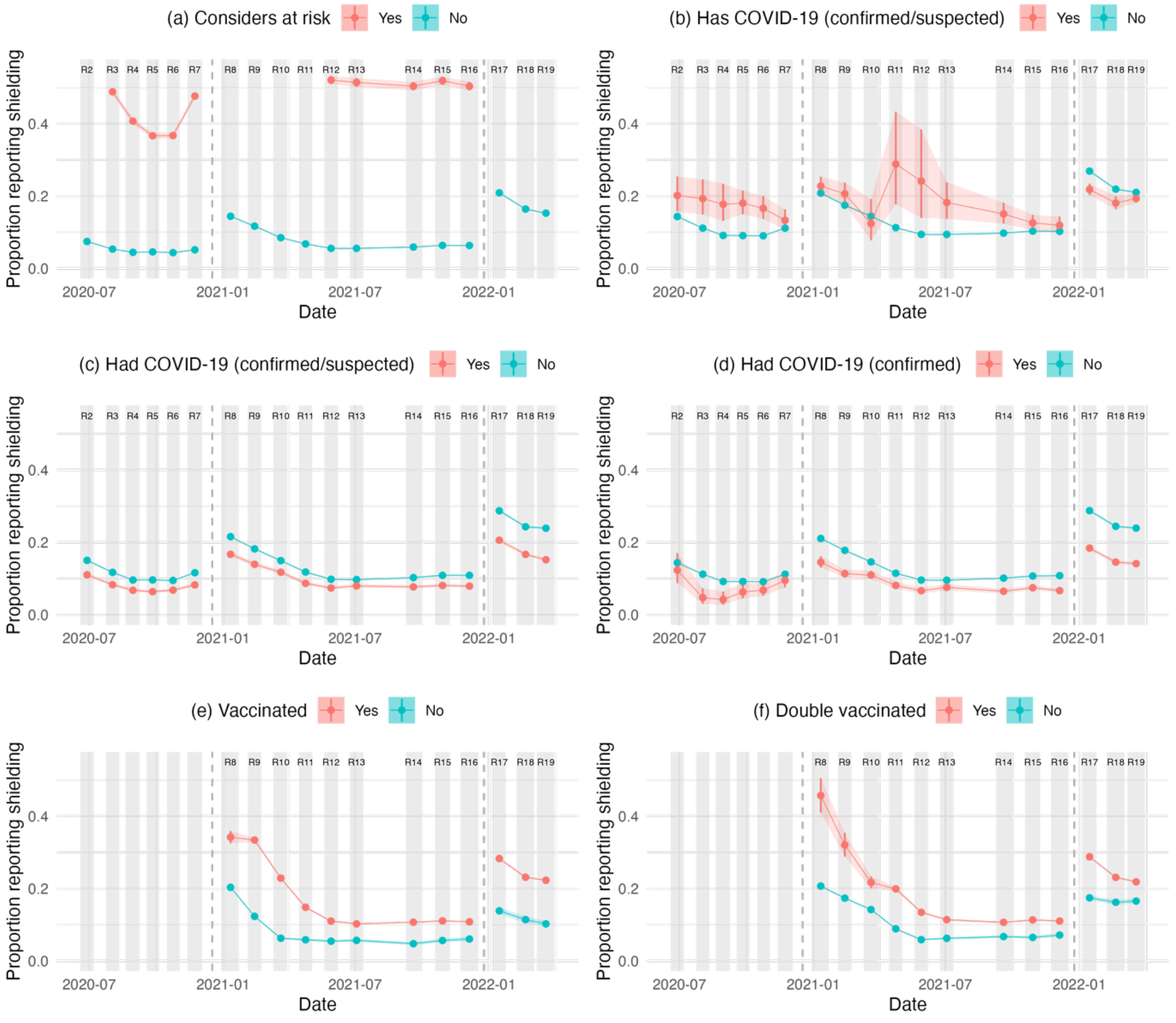

**Figure S9** The proportion of people that report shielding and/or taking specific precautions because they consider themselves "to be at risk of severe illness for COVID-19". Results are conditioned on whether the individual (a) reports considering themselves to be at risk of severe illness, (b) reports a recent confirmed and/or suspected COVID-19 infection, (c) reports a past confirmed and/or suspected COVID-19 infection, (d) reports a past COVID-19 infection confirmed by a positive test, (e) reports receiving at least one dose of a COVID-19 vaccination, and (f) reports receiving at least two doses of a COVID-19 vaccination. The question wording changes between REACT-1 rounds 7 and 8, and again between REACT-1 rounds 16 and 17 – this is marked by a dashed grey line. Shaded areas and vertical lines show 95% confidence intervals about the estimated proportion. The y-axis for (a) people who consider themselves at risk of severe illness is different to the remaining subplots.

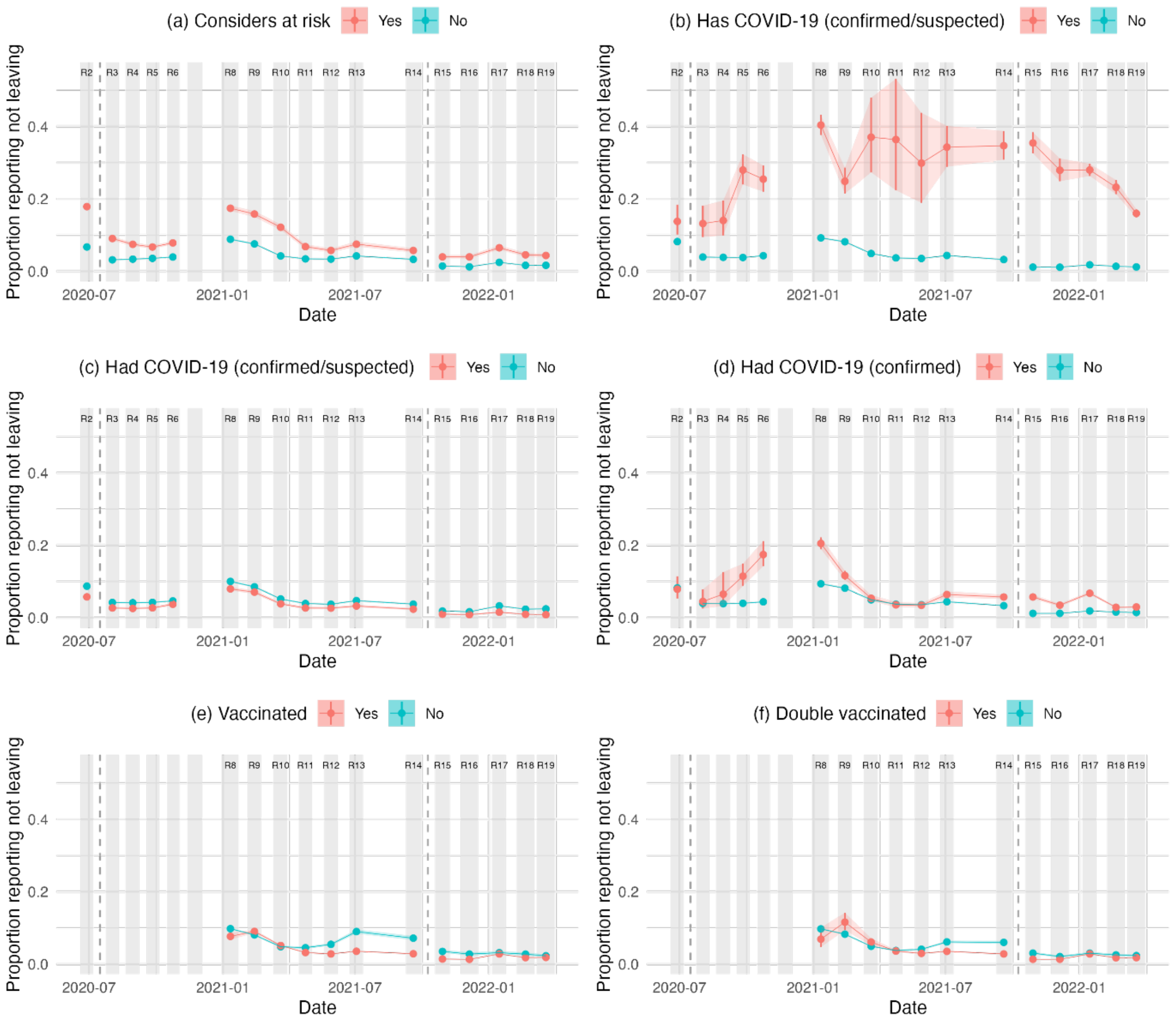

**Figure S10** The proportion of people that report not leaving the home in the 7 days prior to completing the questionnaire. Results are conditioned on whether the individual (a) reports considering themselves to be at risk of severe illness, (b) reports a recent confirmed and/or suspected COVID-19 infection, (c) reports a past confirmed and/or suspected COVID-19 infection, (d) reports a past COVID-19 infection confirmed by a positive test, (e) reports receiving at least one dose of a COVID-19 vaccination, and (f) reports receiving at least two doses of a COVID-19 vaccination. The question wording changes between REACT-1 rounds 2 and 3, and again between REACT-1 rounds 14 and 15 – this is marked by a dashed grey line. This question was not asking in round 7. Shaded areas and vertical lines show 95% confidence intervals about the estimated proportion.

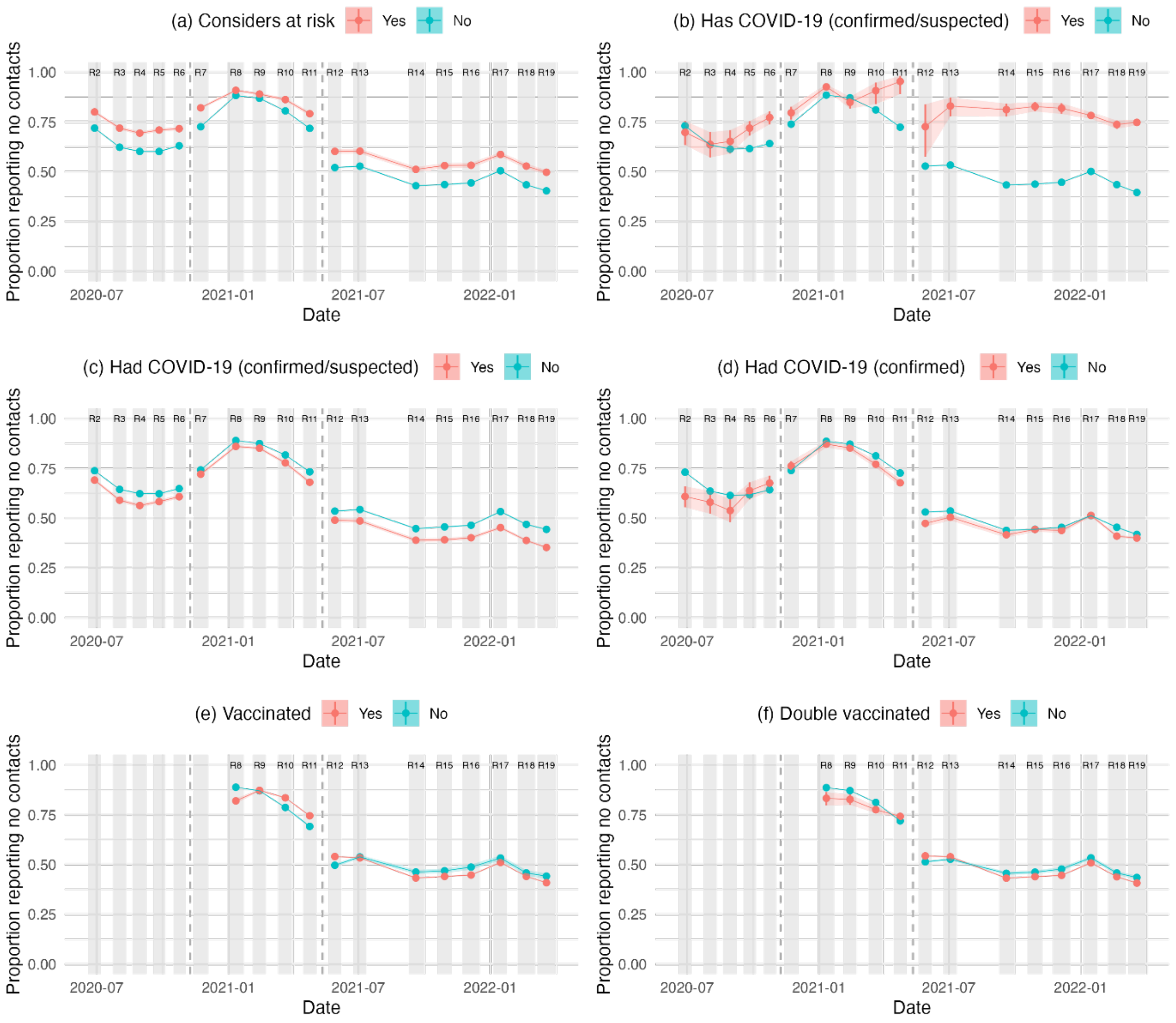

**Figure S11** The proportion of people that report having no contacts outside the home on the day preceding the questionnaire. Results are conditioned on whether the individual (a) reports considering themselves to be at risk of severe illness, (b) reports a recent confirmed and/or suspected COVID-19 infection, (c) reports a past confirmed and/or suspected COVID-19 infection, (d) reports a past COVID-19 infection confirmed by a positive test, (e) reports receiving at least one dose of a COVID-19 vaccination, and (f) reports receiving at least two doses of a COVID-19 vaccination. The question wording changes between REACT-1 rounds 6 and 7, and again between REACT-1 rounds 11 and 12 – this is marked by a dashed grey line. Shaded areas and vertical lines show 95% confidence intervals about the estimated proportion.

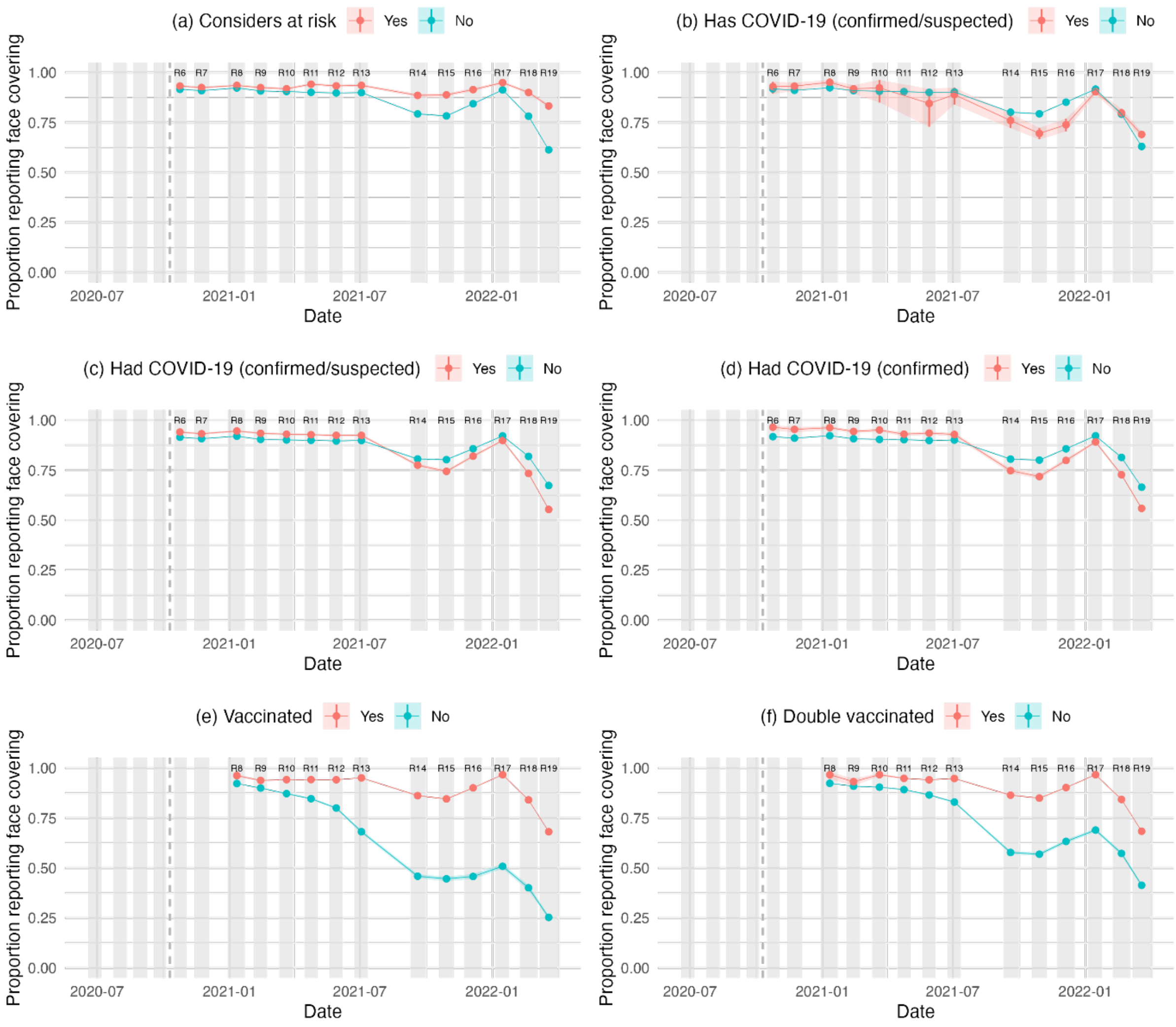

**Figure S12** The proportion of people that report wearing a face covering at least sometimes when leaving the home. Results are conditioned on whether the individual (a) reports considering themselves to be at risk of severe illness, (b) reports a recent confirmed and/or suspected COVID-19 infection, (c) reports a past confirmed and/or suspected COVID-19 infection, (d) reports a past COVID-19 infection confirmed by a positive test, (e) reports receiving at least one dose of a COVID-19 vaccination, and (f) reports receiving at least two doses of a COVID-19 vaccination. This question was asked in REACT-1 round 6 onwards. Shaded areas and vertical lines show 95% confidence intervals about the estimated proportion.

#### SECTION 2E: REASONS FOR LEAVING THE HOME

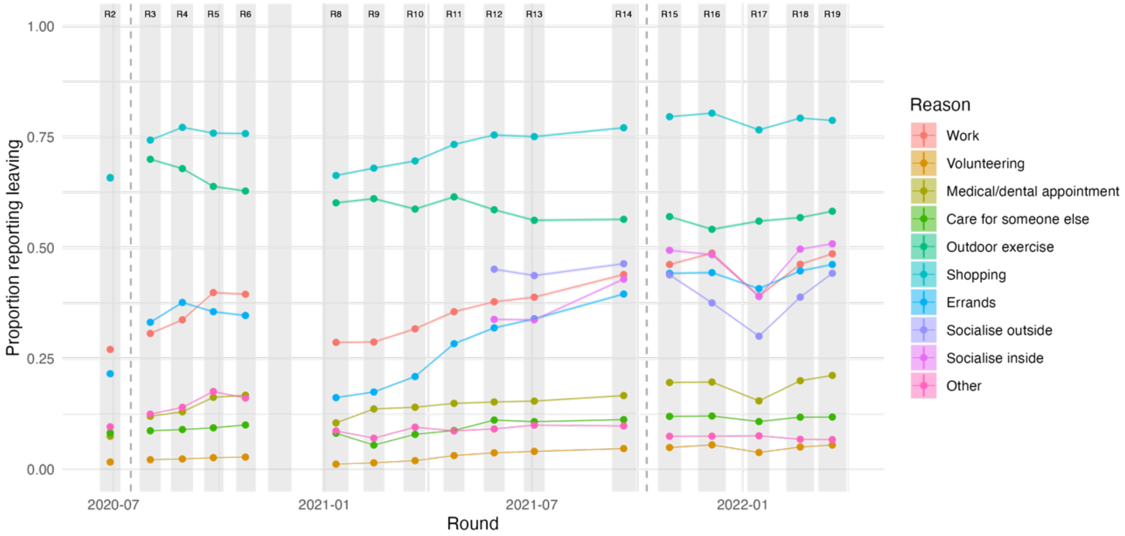

**Figure S13** The proportion of people that report leaving the home for various reasons. Coloured shaded areas and vertical lines show 95% confidence intervals about the estimated proportion. Vertical dashed lines indicate changes in question wording. This question was not asked in study round 7.

### SUPPLEMENTARY SECTION 3: LOGISTIC REGRESSION MODELS

Figures S14 to S24 present the results from logistic regression models that isolate the impact of each demographic variable on various reported behaviours. We include a summary of the results here.

Those that are more likely to report leaving to go to work (Figure S17) are:

- Those aged 18-34 years, and to a lesser extent aged 35-64, when compared to those aged 65+ and 5-17.
- Males
- Those not living in London in rounds 2 to 13, with the strongest effect being in rounds 8 and 9, as well as round 17. This effect reverses in round 19, with those living in London being more likely to report leaving home to go to work.
- Those in areas with greater socioeconomic deprivation (deprivation quintiles 4 & 5 [least deprived] are less likely to report leaving to work than those in deprivation quintile 1 [most deprived])
- Those in larger households

Those that are more likely to report leaving to volunteer (Figure S18) are:

- Older people (those aged 35-64 sand 65+ years)
- Females in rounds 10 onwards
- People living in London (and those living in the South West in rounds 4 to 6 and 14 to 17)
- People in areas with lower socioeconomic deprivation
- People that live alone

Those that are more likely to report leaving to attend a medical or dental appointment (Figure S19) are:

- Older people (those aged 35-64 sand 65+ years)
- Females

Those that are more likely to report leaving to care for someone else (Figure S20) are:

- People aged 35-64 years, and also those aged 65+, when compared to those aged 18-34 and those aged 5-17
- People living outside of London, particularly in the North East in earlier rounds
- People in areas with lower socioeconomic deprivation
- People living alone or with one other person (compared to those living in larger households)

Those that are more likely to report leaving for outdoor exercise (Figure S21) are:

- People living in London
- People in areas with lower socioeconomic deprivation

- People identifying as white
- People living in smaller households

Those that are more likely to report leaving to go shopping (Figure S22) are:

- People aged 35-64 years. Also those aged 65+ in rounds 4 to 6 and rounds 11 to 19.
- Females
- People in areas with lower socioeconomic deprivation in rounds 10 to 19
- People living alone

Those that are more likely to report leaving for errands (Figure S23) are:

- People aged 18-34 years
- People in London

Those that are more likely to report leaving for other reasons (Figure S24) are:

- People aged 65+ years and 5-17 years (with exceptions to the latter in study rounds 17, 18, and 19)
- Females
- People in wealthier areas
- People not identifying as white

These “other” reasons should not be compared across rounds because the proportion of people that ticked “other” depends on which pre-set options are available.

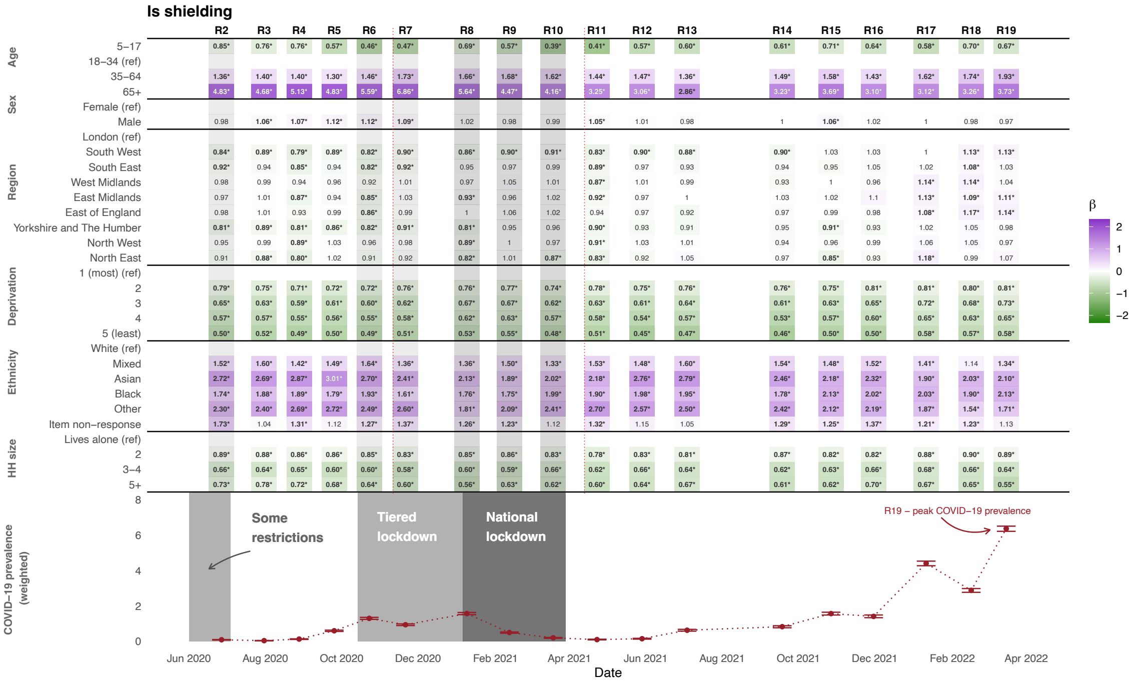

**Figure S14.** Odds ratios for whether an individual would report <u>shielding and/or taking specific precautions</u>. Estimated effects are mutually adjusted for the other variables listed in the table. * indicates the odds ratio is statistically significantly different from 1 at the $\alpha = 0.05$ significance level. The question wording changed between study rounds 7 and 8, and again between study rounds 16 and 17 as indicated by vertical dashed lines – see supplementary section 1 for further detail.

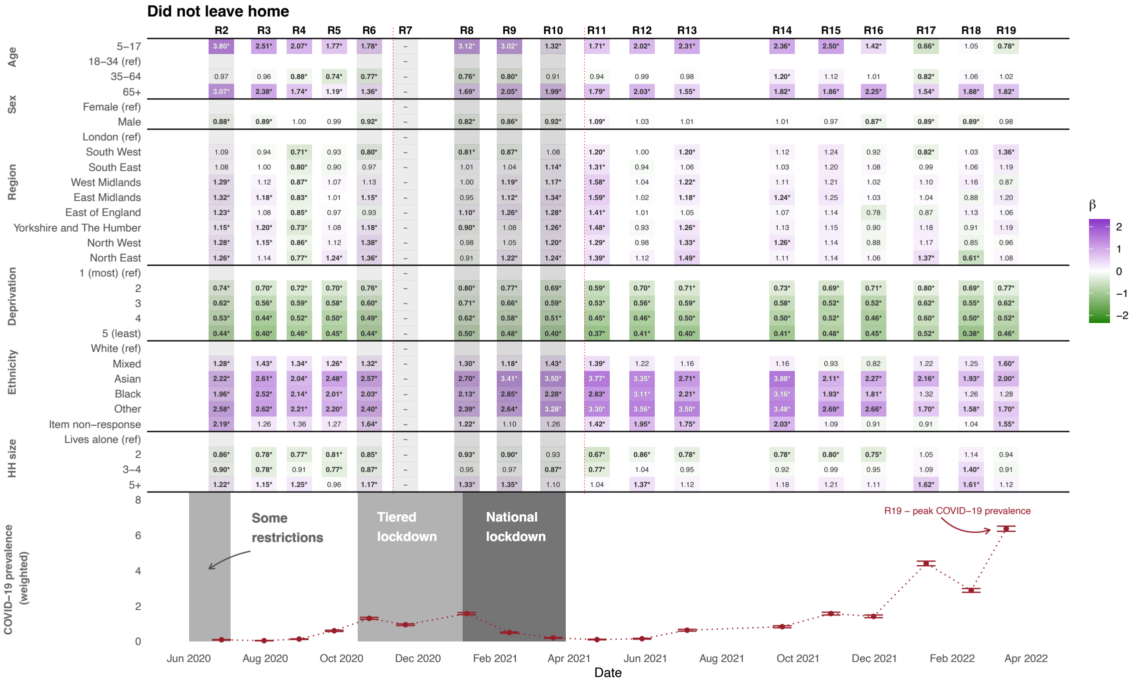

**Figure S15.** Odds ratios for whether an individual would report <u>not leaving the house in the seven days preceding completing the questionnaire</u>. Estimated effects are mutually adjusted for the other variables listed in the table. * indicates the odds ratio is statistically significantly different from 1 at the $\alpha = 0.05$ significance level. This question was not asked in round 7, and the question wording changed between study rounds 2 and 3, and again between study rounds 14 to 15, denoted with a vertical dashed line.

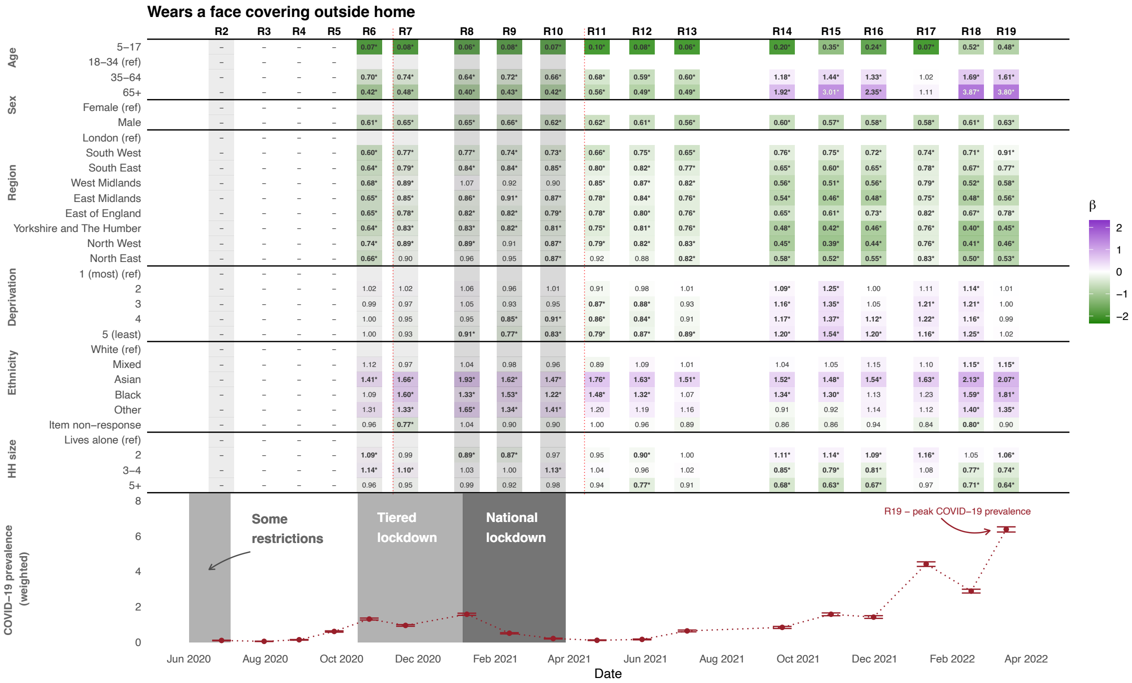

**Figure S16.** Odds ratios for whether an individual would report wearing a face covering outside the home. Estimated effects are mutually adjusted for the other variables listed in the table. * indicates the odds ratio is statistically significantly different from 1 at the $\alpha = 0.05$ significance level. This question was not asked prior to study round 6.

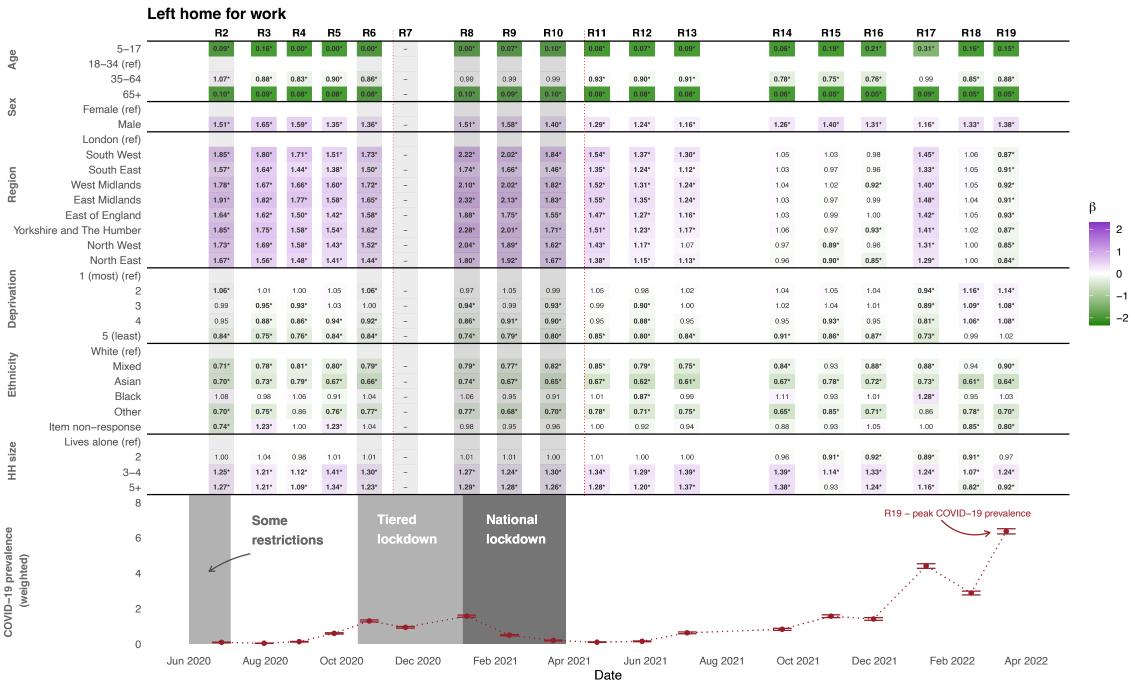

**Figure S17.** Odds ratios for whether an individual would report leaving home to go to work. Estimated effects are mutually adjusted for the other variables listed in the table. * indicates the odds ratio is statistically significantly different from 1 at the $\alpha = 0.05$ significance level. This question was not asked in round 7, and the question wording changed between study rounds 2 and 3, and again between study rounds 14 to 15, denoted with a vertical dashed line.

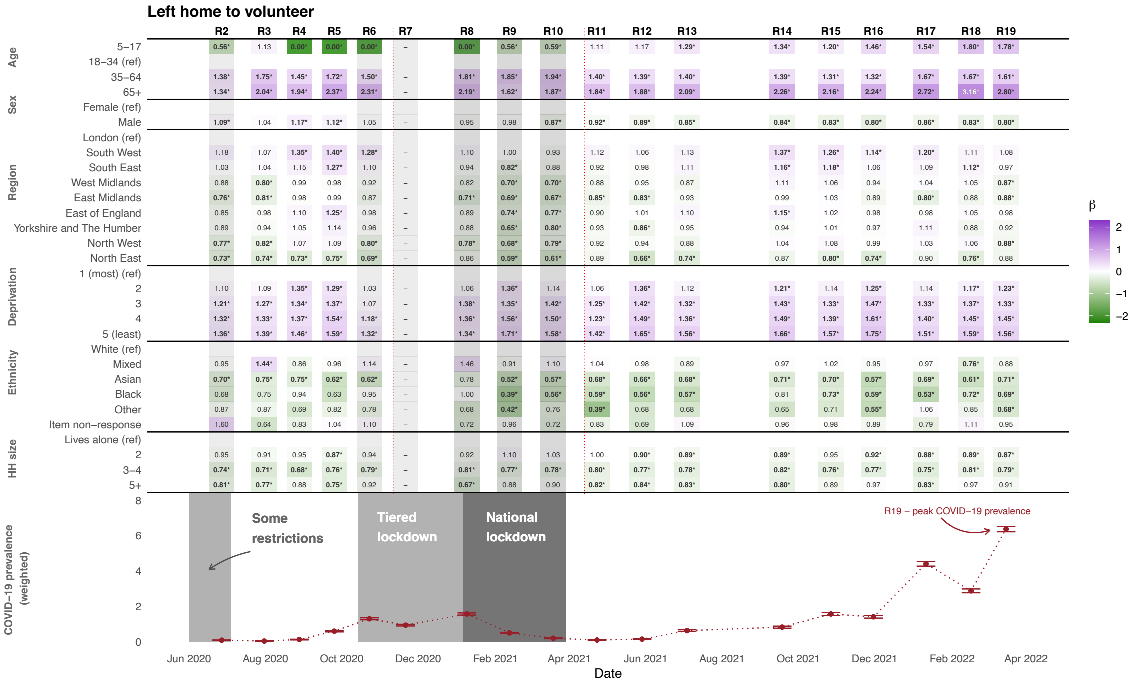

**Figure S18.** Odds ratios for whether an individual would report <u>leaving home to volunteer</u>. Estimated effects are mutually adjusted for the other variables listed in the table. * indicates the odds ratio is statistically significantly different from 1 at the $\alpha = 0.05$ significance level. This question was not asked in round 7, and the question wording changed between study rounds 2 and 3, and again between study rounds 14 to 15, denoted with a vertical dashed line.

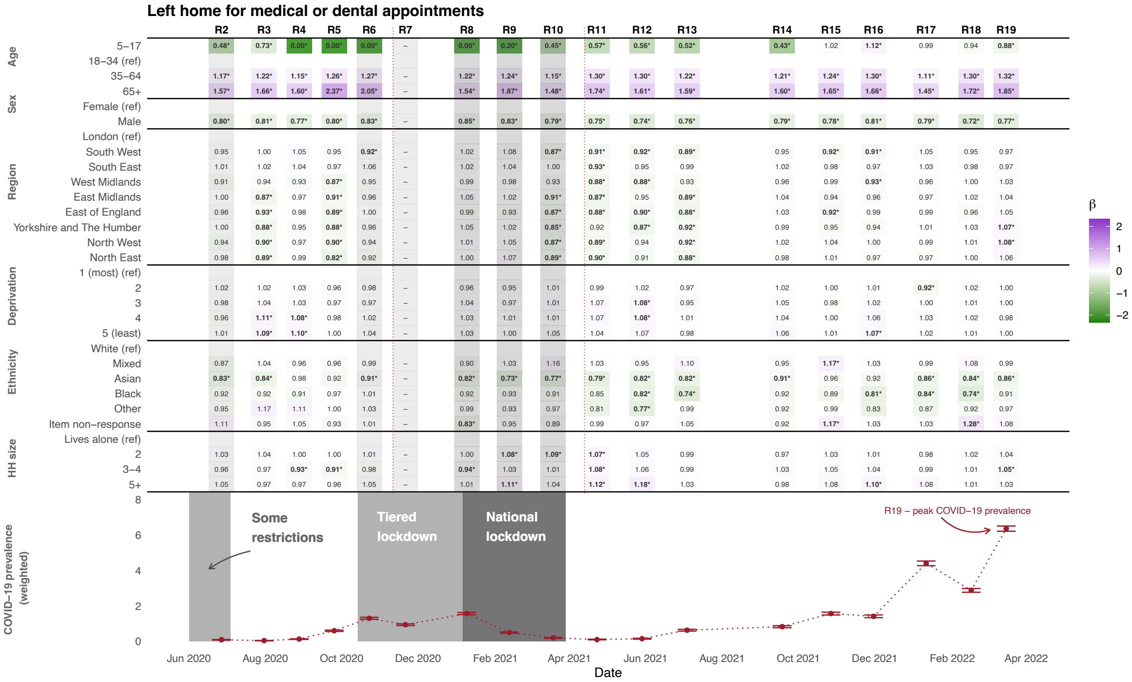

**Figure S19.** Odds ratios for whether an individual would report leaving home for medical and/or dental care. Estimated effects are mutually adjusted for the other variables listed in the table. * indicates the odds ratio is statistically significantly different from 1 at the $\alpha = 0.05$ significance level. This question was not asked in round 7, and the question wording changed between study rounds 2 and 3, and again between study rounds 14 to 15, denoted with a vertical dashed line.

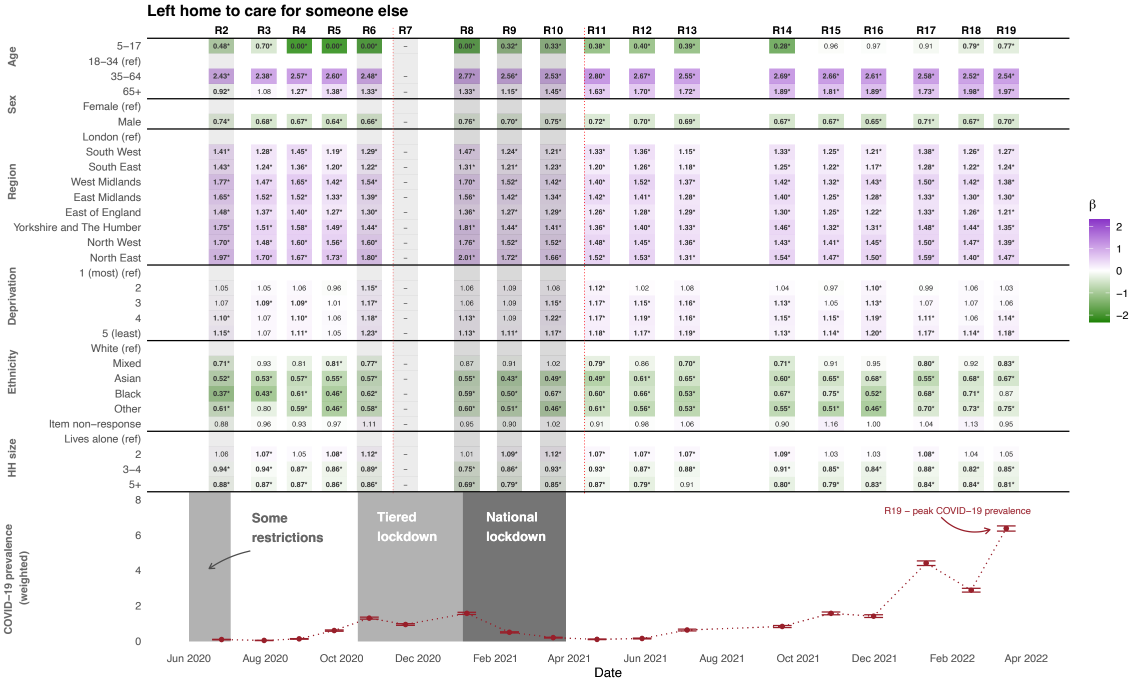

**Figure S20.** Odds ratios for whether an individual would report leaving home to care for somebody else. Estimated effects are mutually adjusted for the other variables listed in the table. * indicates the odds ratio is statistically significantly different from 1 at the $\alpha = 0.05$ significance level. This question was not asked in round 7, and the question wording changed between study rounds 2 and 3, and again between study rounds 14 to 15, denoted with a vertical dashed line.

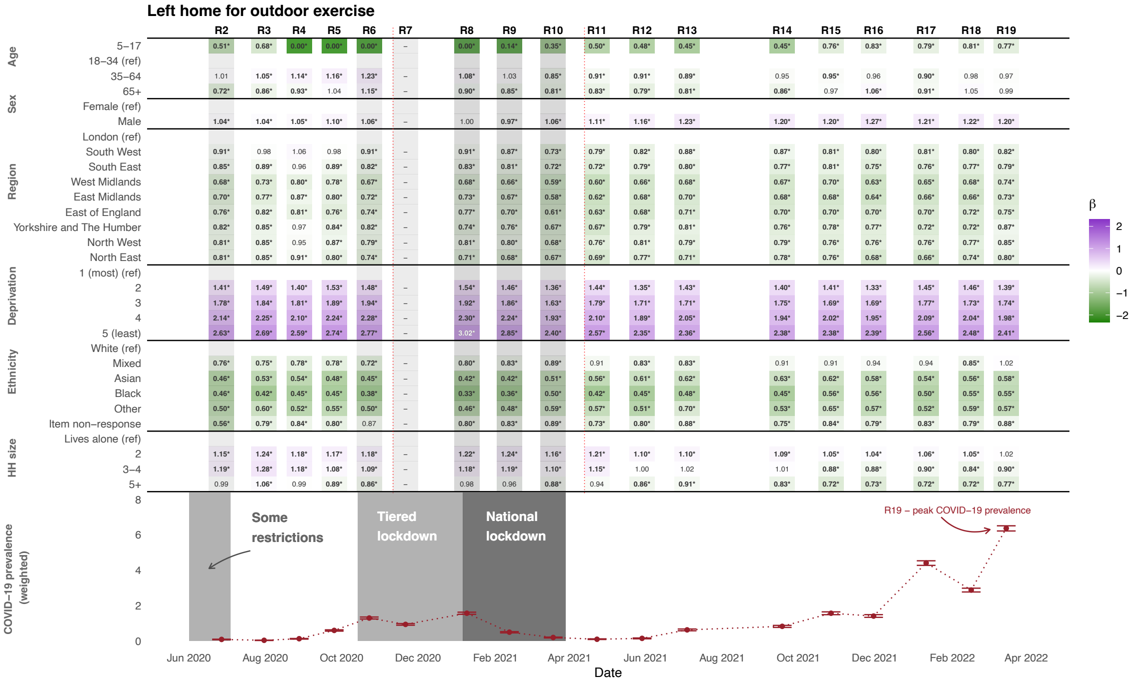

**Figure S21.** Odds ratios for whether an individual would report <u>leaving home to exercise outdoors</u>. Estimated effects are mutually adjusted for the other variables listed in the table. * indicates the odds ratio is statistically significantly different from 1 at the $\alpha = 0.05$ significance level. This question was not asked in round 7, and the question wording changed between study rounds 2 and 3, and again between study rounds 14 to 15, denoted with a vertical dashed line.

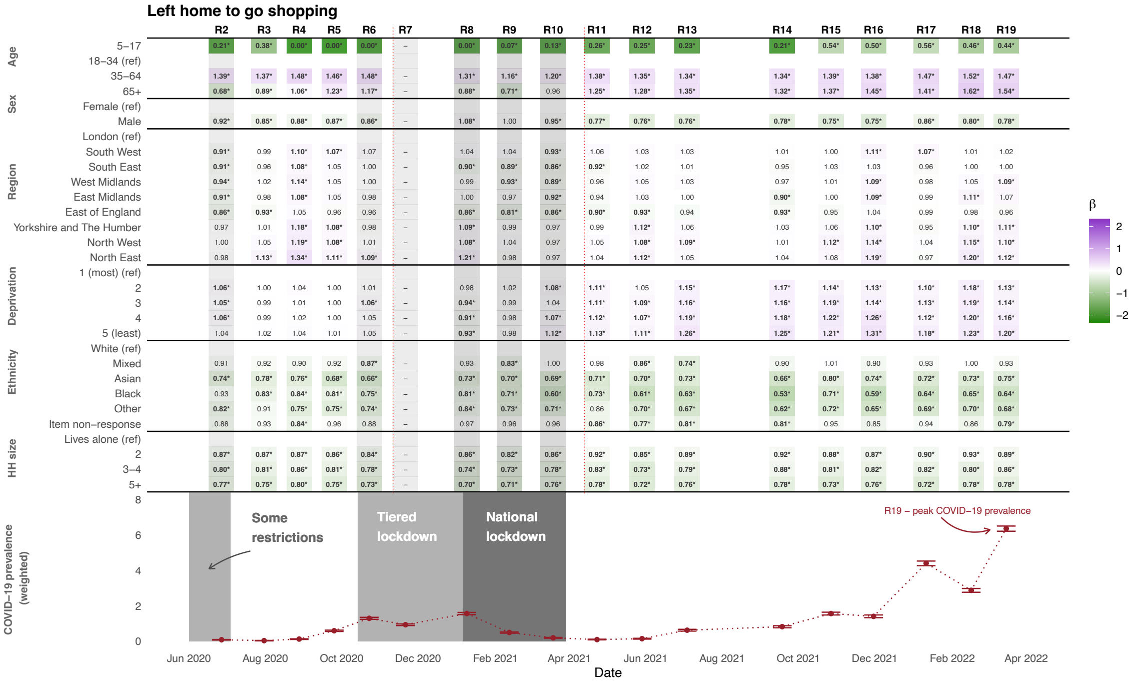

**Figure S22.** Odds ratios for whether an individual would report <u>leaving home for shopping</u>. Estimated effects are mutually adjusted for the other variables listed in the table. * indicates the odds ratio is statistically significantly different from 1 at the $\alpha = 0.05$ significance level. This question was not asked in round 7, and the question wording changed between study rounds 2 and 3, and again between study rounds 14 to 15, denoted with a vertical dashed line.

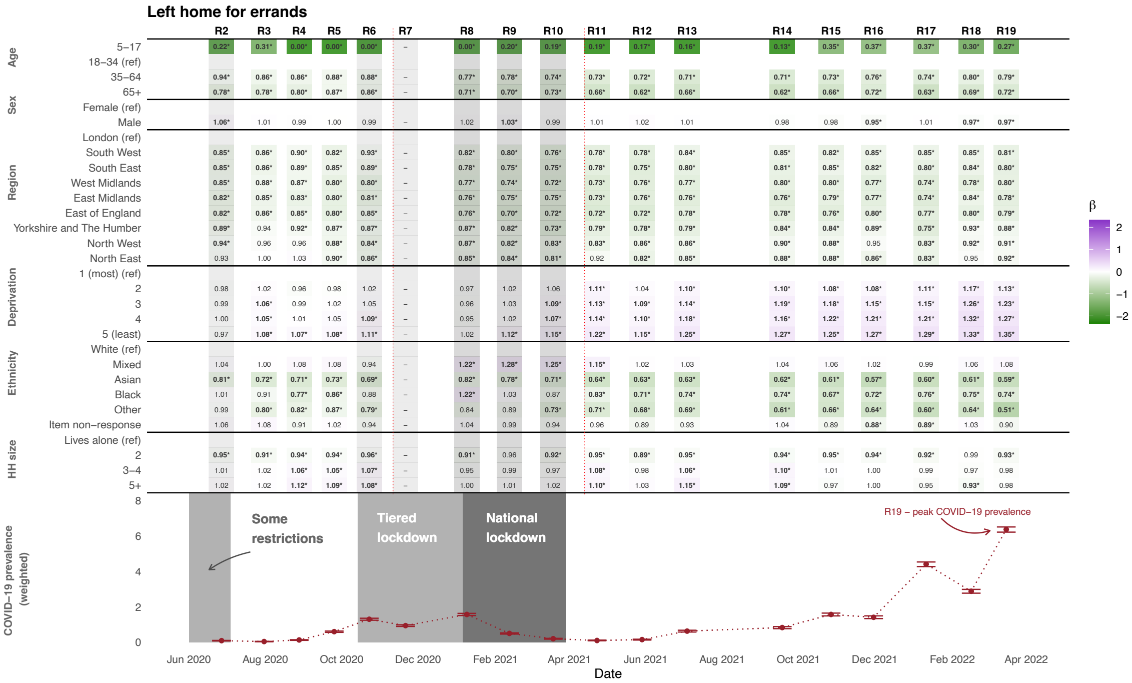

**Figure S23.** Odds ratios for whether an individual would report <u>leaving home for errands</u>. Estimated effects are mutually adjusted for the other variables listed in the table. * indicates the odds ratio is statistically significantly different from 1 at the $\alpha = 0.05$ significance level. This question was not asked in round 7, and the question wording changed between study rounds 2 and 3, and again between study rounds 14 to 15, denoted with a vertical dashed line.

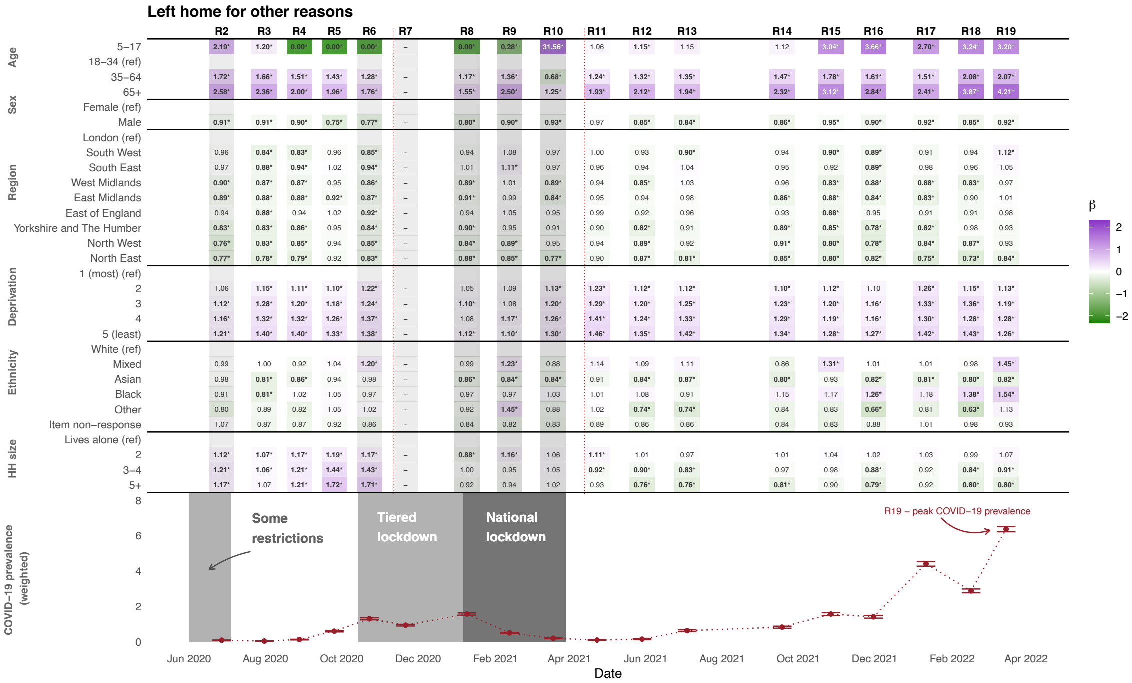

**Figure S24.** Odds ratios for whether an individual would report leaving home for other reasons. Estimated effects are mutually adjusted for the other variables listed in the table. * indicates the odds ratio is statistically significantly different from 1 at the $\alpha = 0.05$ significance level. This question was not asked in round 7, and the question wording changed between study rounds 2 and 3, and again between study rounds 14 to 15, denoted with a vertical dashed line. The change in question wording is particularly important here, as "other" may include or exclude specific reasons depending on the available options (see supplementary table S3).

Figures S25 to S35 present the results from logistic regression models that isolate the impact of each demographic variable on various reported behaviours, with a further adjustment for a binary variable indicating whether the respondent thought they had had COVID in the preceding two weeks (question 'COVIDA', described in section 1a of the supplementary material) .

The results of the logistic regression models are robust to this additional adjustment.

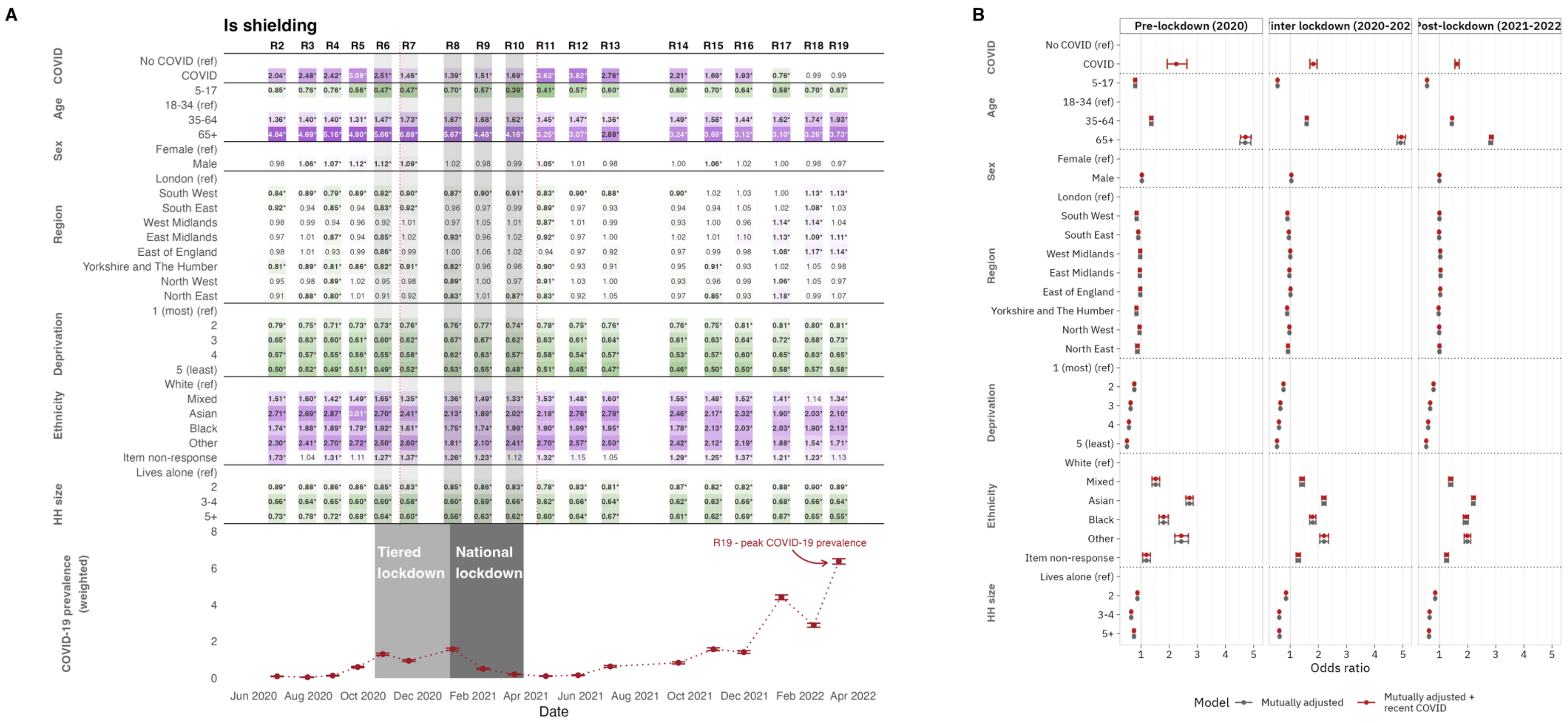

**Figure S25. A.** Odds ratios for whether an individual would report shielding and/or taking specific precautions. Estimated effects are mutually adjusted for the other variables listed in the table. * indicates the odds ratio is statistically significantly different from 1 at the $\alpha = 0.05$ significance level. The question wording changed between study rounds 7 and 8, and again between study rounds 16 and 17 as indicated by vertical dashed red lines – see supplementary section 1 for further detail.

**B.** Forest plot comparing odds ratios for models with and without adjustment for suspected recent COVID-19.

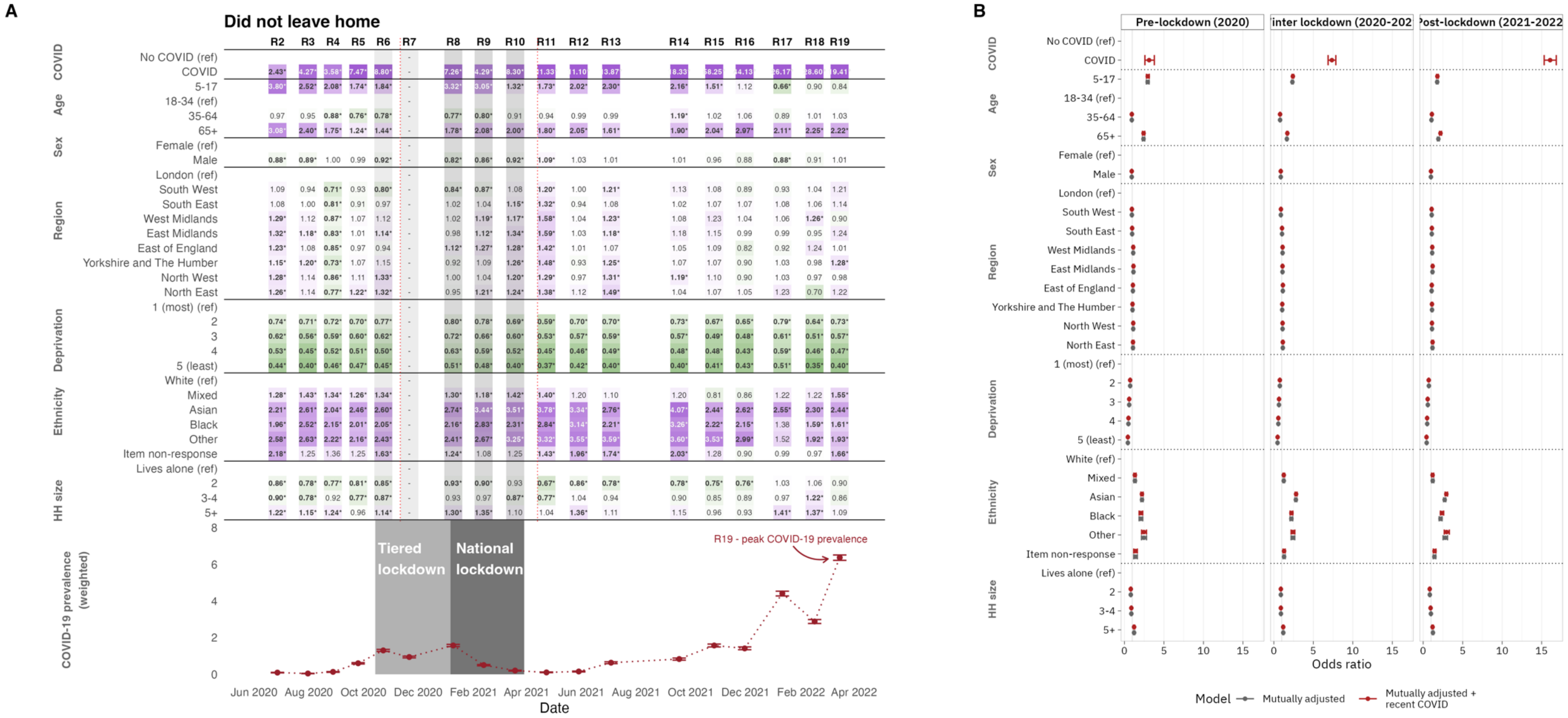

**Figure S26. A.** Odds ratios for whether an individual would report <u>not leaving the house in the seven days preceding completing the questionnaire</u>. Estimated effects are mutually adjusted for the other variables listed in the table. * indicates the odds ratio is statistically significantly different from 1 at the $\alpha = 0.05$ significance level. This question was not asked in round 7, and the question wording changed between study rounds 2 and 3, and again between study rounds 14 to 15, denoted with a vertical dashed line.

**B.** Forest plot comparing odds ratios for models with and without adjustment for suspected recent COVID-19.

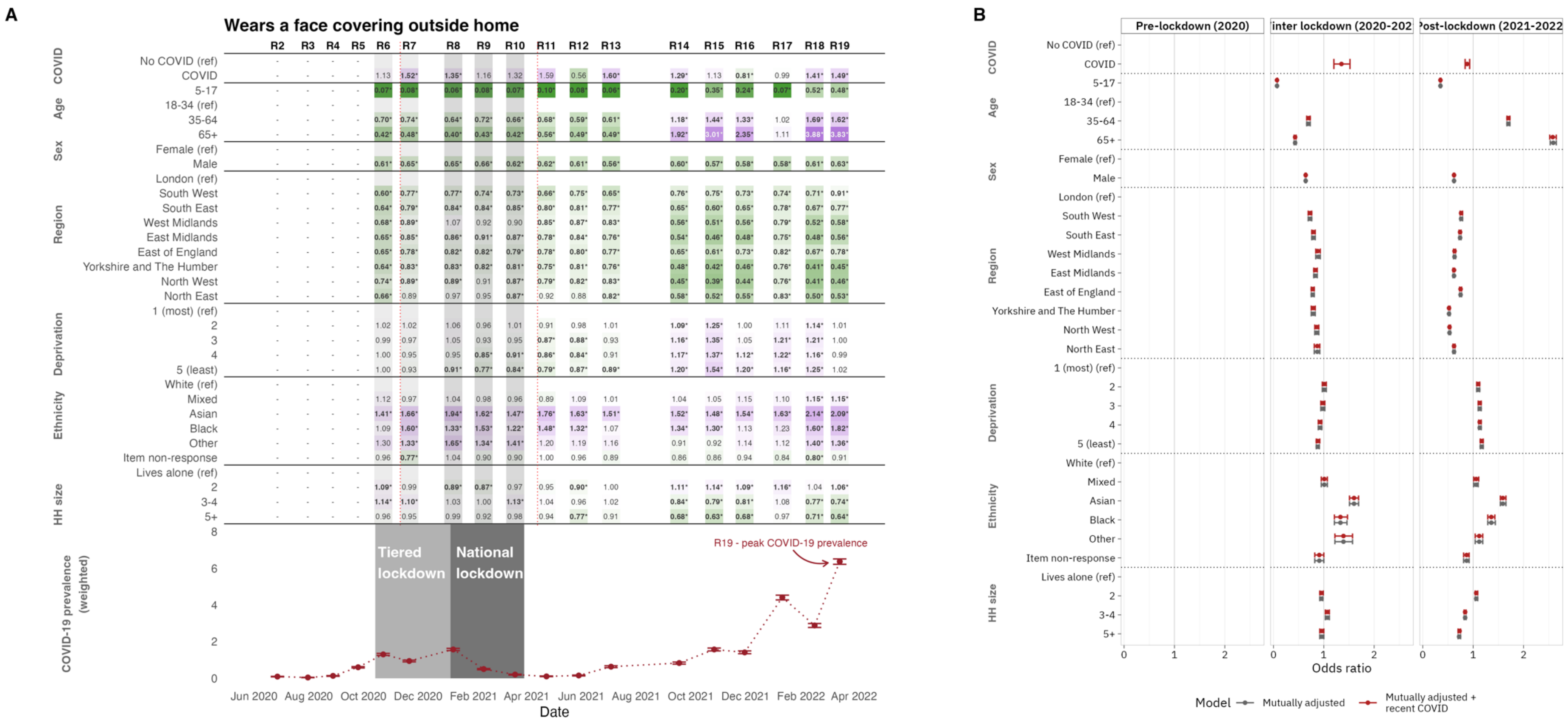

**Figure S27. A.** Odds ratios for whether an individual would report <u>wearing a face covering outside the home</u>. Estimated effects are mutually adjusted for the other variables listed in the table. * indicates the odds ratio is statistically significantly different from 1 at the $\alpha = 0.05$ significance level. This question was not asked prior to study round 6.

**B.** Forest plot comparing odds ratios for models with and without adjustment for suspected recent COVID-19.

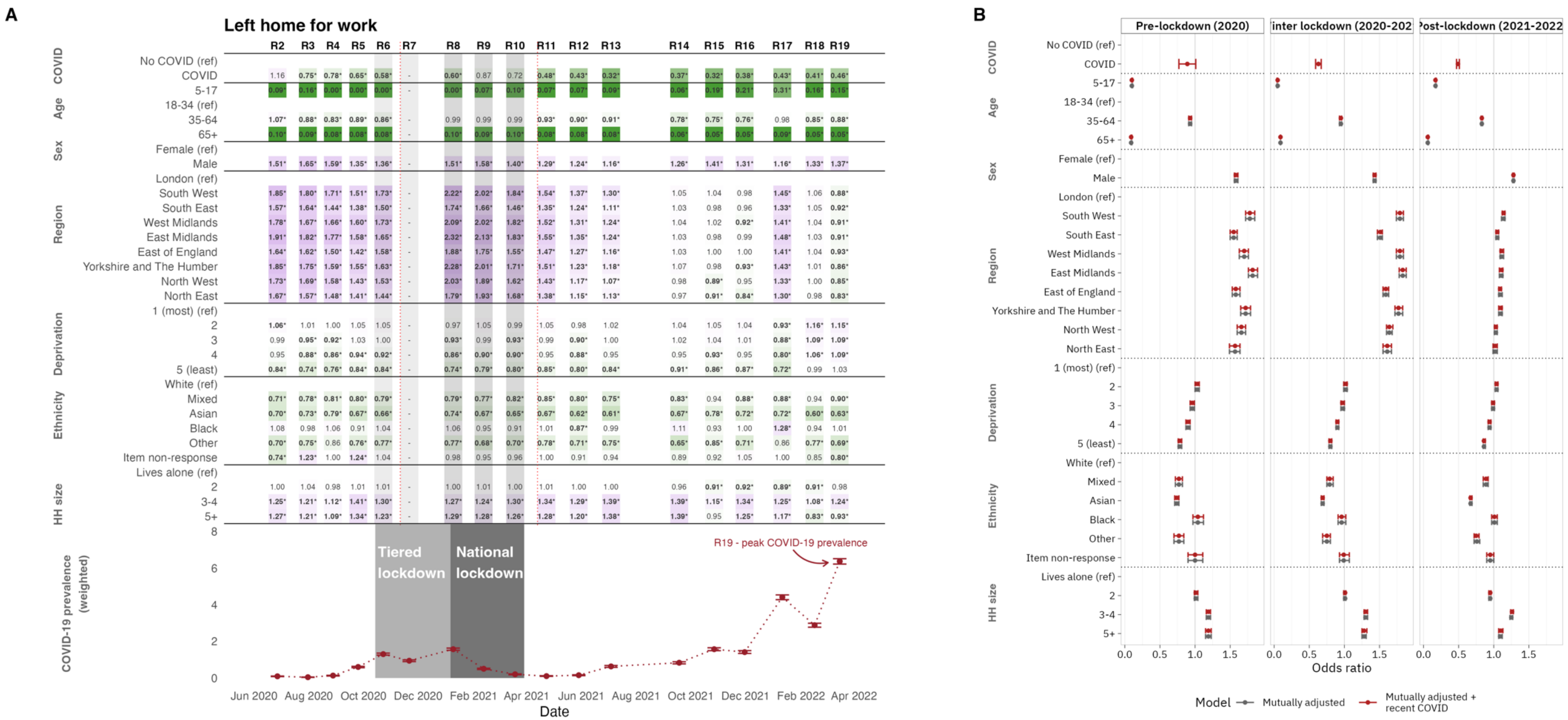

**Figure S28. A.** Odds ratios for whether an individual would report leaving home to go to work. Estimated effects are mutually adjusted for the other variables listed in the table. * indicates the odds ratio is statistically significantly different from 1 at the $\alpha = 0.05$ significance level. This question was not asked in round 7, and the question wording changed between study rounds 2 and 3, and again between study rounds 14 to 15, denoted with a vertical dashed line. **B.** Forest plot comparing odds ratios for models with and without adjustment for suspected recent COVID-19.

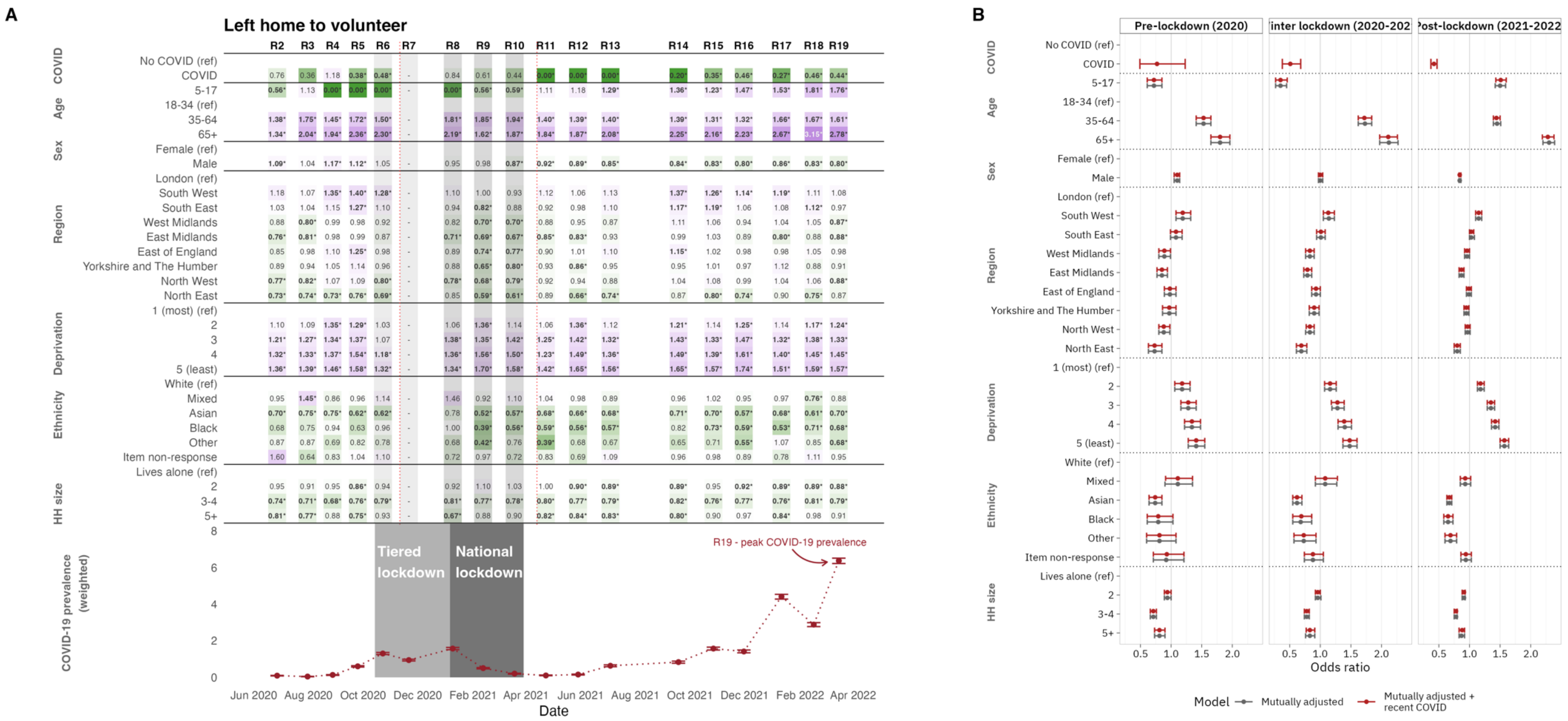

**Figure S29. A.** Odds ratios for whether an individual would report leaving home to volunteer. Estimated effects are mutually adjusted for the other variables listed in the table. * indicates the odds ratio is statistically significantly different from 1 at the $\alpha = 0.05$ significance level. This question was not asked in round 7, and the question wording changed between study rounds 2 and 3, and again between study rounds 14 to 15, denoted with a vertical dashed line. **B.** Forest plot comparing odds ratios for models with and without adjustment for suspected recent COVID-19.

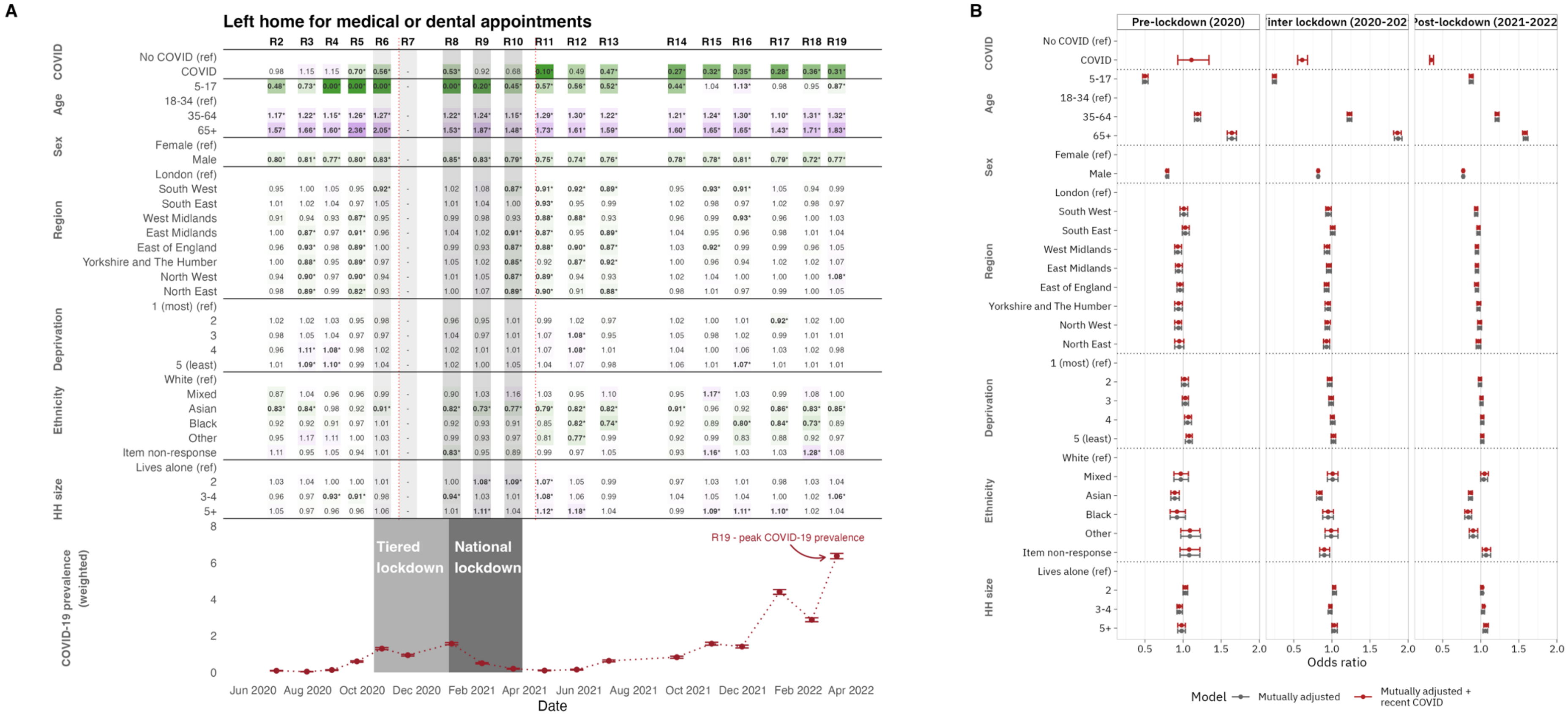

**Figure S30. A.** Odds ratios for whether an individual would report <u>leaving home for medical and/or dental care</u>. Estimated effects are mutually adjusted for the other variables listed in the table. * indicates the odds ratio is statistically significantly different from 1 at the $\alpha = 0.05$ significance level. This question was not asked in round 7, and the question wording changed between study rounds 2 and 3, and again between study rounds 14 to 15, denoted with a vertical dashed line.

**B.** Forest plot comparing odds ratios for models with and without adjustment for suspected recent COVID-19.

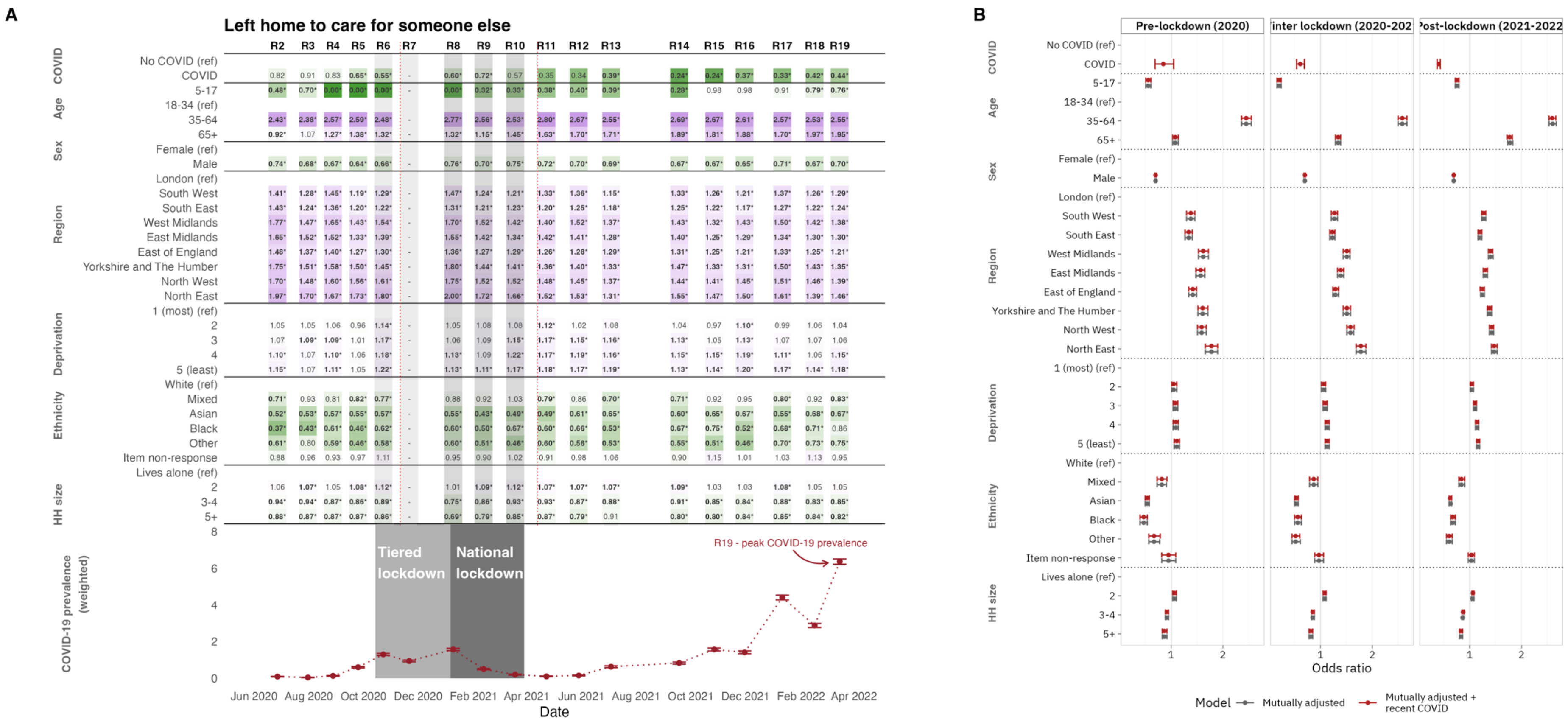

**Figure S31. A.** Odds ratios for whether an individual would report <u>leaving home to care for somebody else</u>. Estimated effects are mutually adjusted for the other variables listed in the table. * indicates the odds ratio is statistically significantly different from 1 at the $\alpha = 0.05$ significance level. This question was not asked in round 7, and the question wording changed between study rounds 2 and 3, and again between study rounds 14 to 15, denoted with a vertical dashed line.

**B.** Forest plot comparing odds ratios for models with and without adjustment for suspected recent COVID-19.

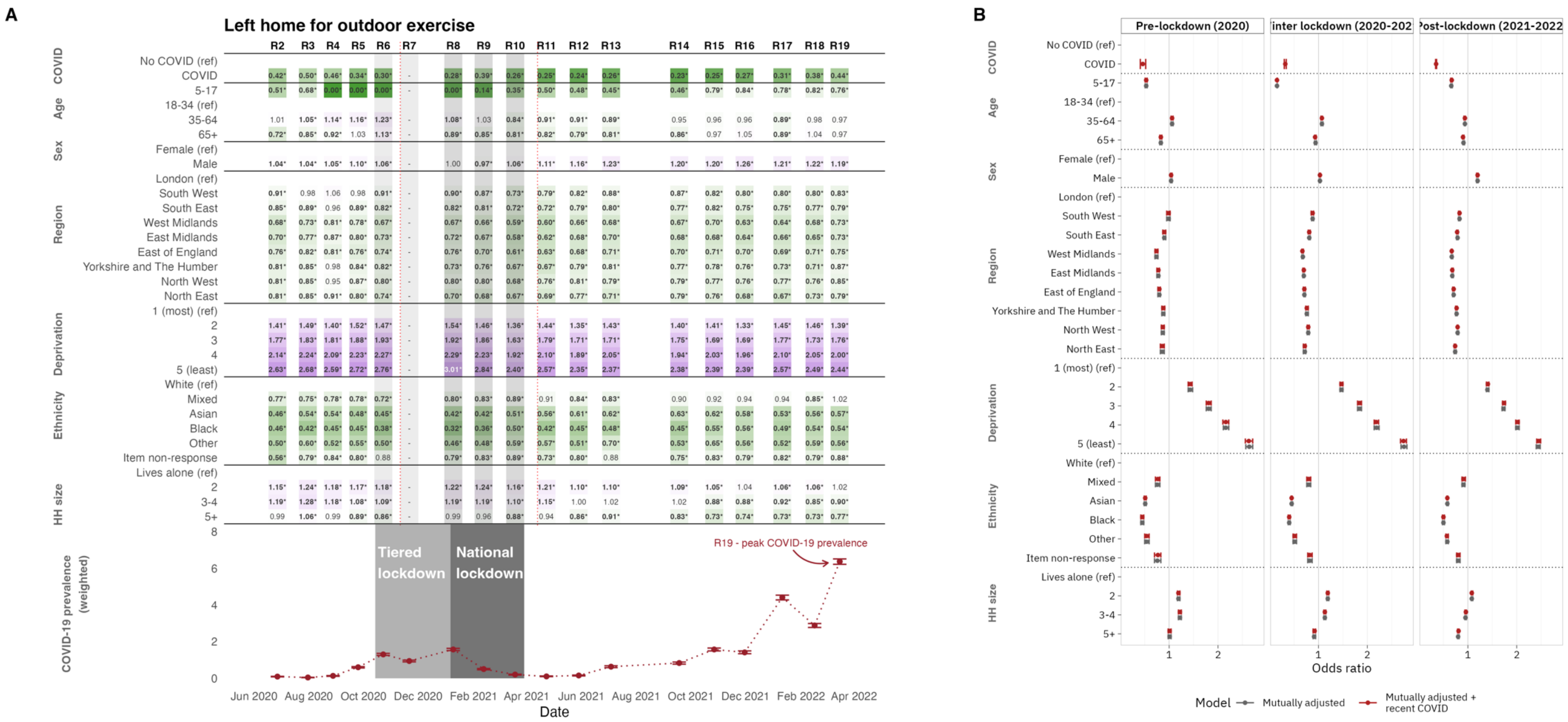

**Figure S32. A.** Odds ratios for whether an individual would report <u>leaving home to exercise outdoors</u>. Estimated effects are mutually adjusted for the other variables listed in the table. * indicates the odds ratio is statistically significantly different from 1 at the $\alpha = 0.05$ significance level. This question was not asked in round 7, and the question wording changed between study rounds 2 and 3, and again between study rounds 14 to 15, denoted with a vertical dashed line.

**B.** Forest plot comparing odds ratios for models with and without adjustment for suspected recent COVID-19.

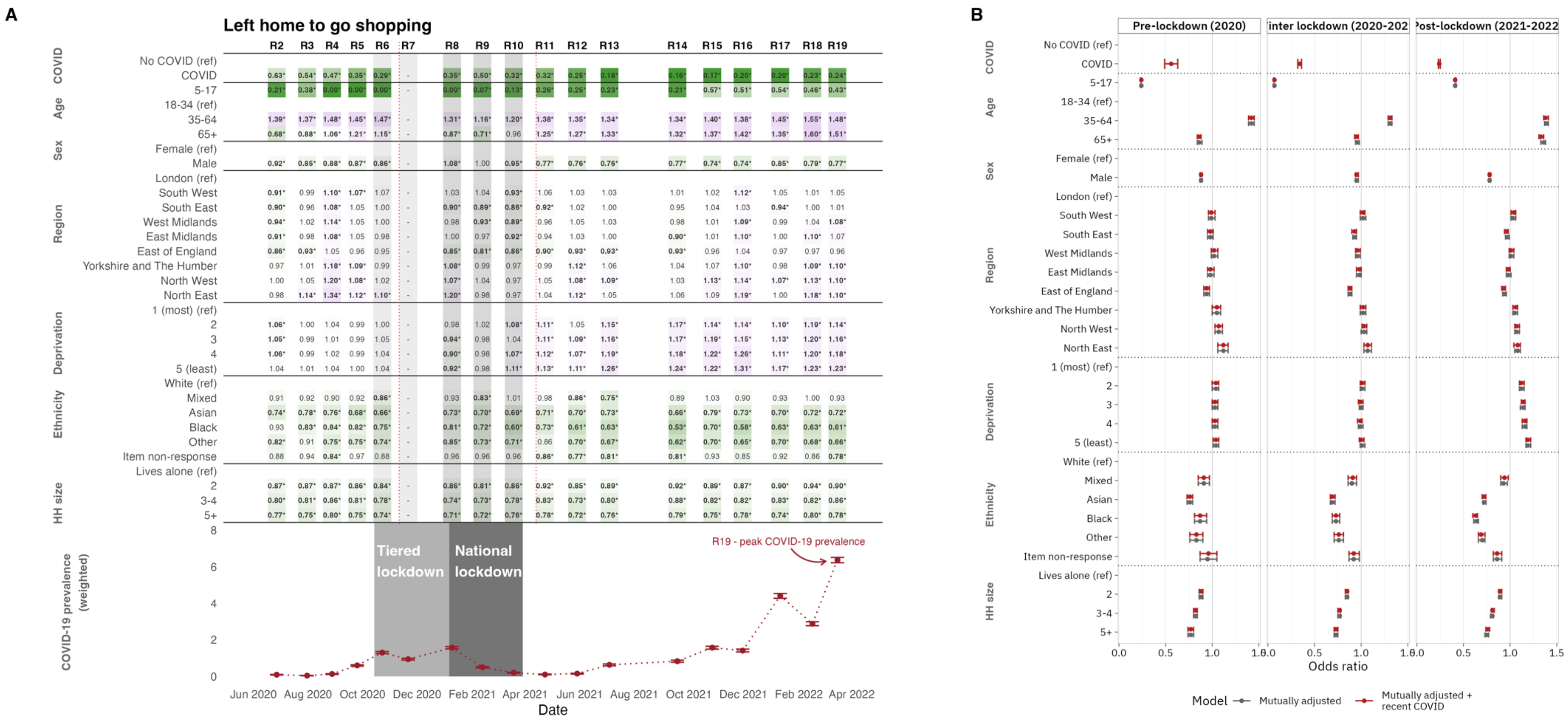

**Figure S33. A.** Odds ratios for whether an individual would report <u>leaving home for shopping</u>. Estimated effects are mutually adjusted for the other variables listed in the table. * indicates the odds ratio is statistically significantly different from 1 at the $\alpha = 0.05$ significance level. This question was not asked in round 7, and the question wording changed between study rounds 2 and 3, and again between study rounds 14 to 15, denoted with a vertical dashed line. **B.** Forest plot comparing odds ratios for models with and without adjustment for suspected recent COVID-19.

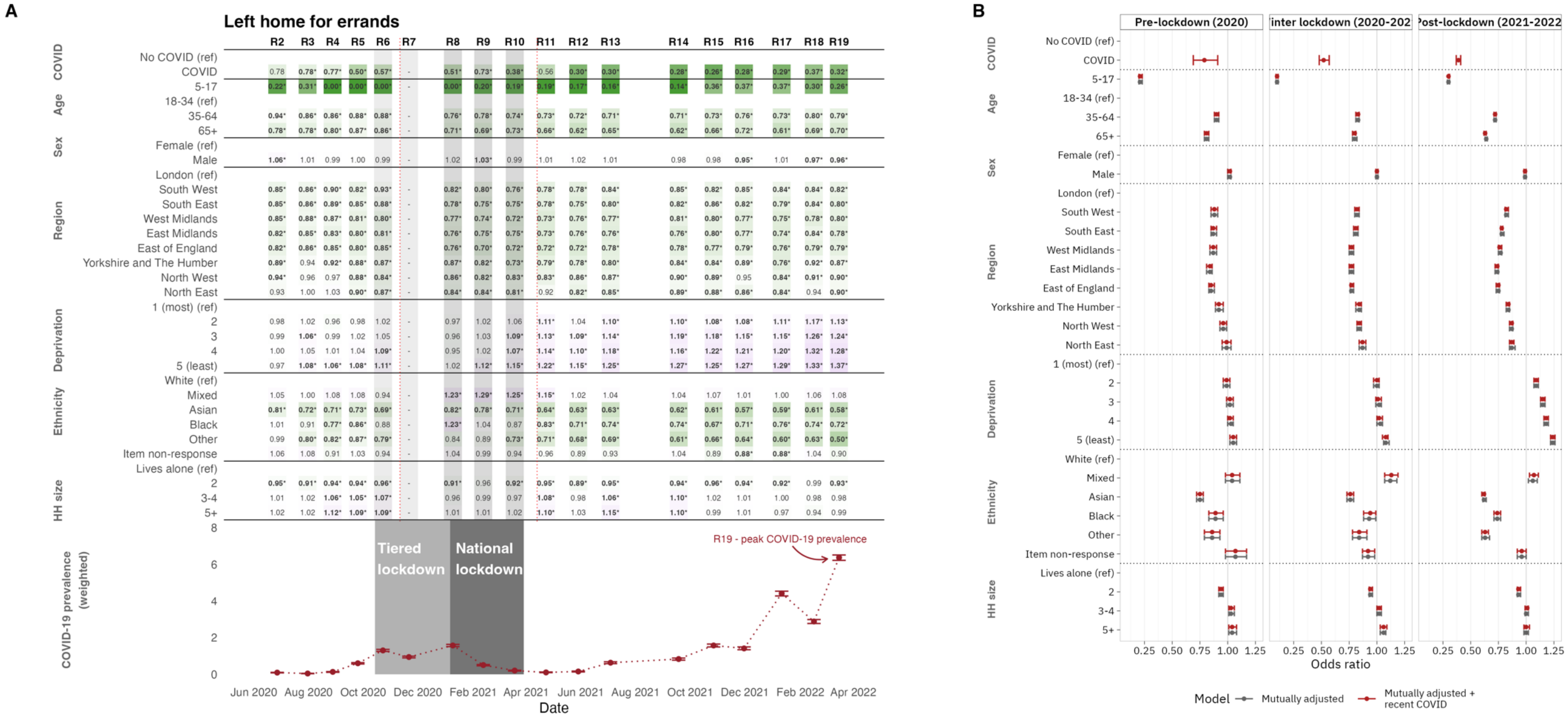

**Figure S34. A.** Odds ratios for whether an individual would report leaving home for errands. Estimated effects are mutually adjusted for the other variables listed in the table. * indicates the odds ratio is statistically significantly different from 1 at the $\alpha = 0.05$ significance level. This question was not asked in round 7, and the question wording changed between study rounds 2 and 3, and again between study rounds 14 to 15, denoted with a vertical dashed line. **B.** Forest plot comparing odds ratios for models with and without adjustment for suspected recent COVID-19.

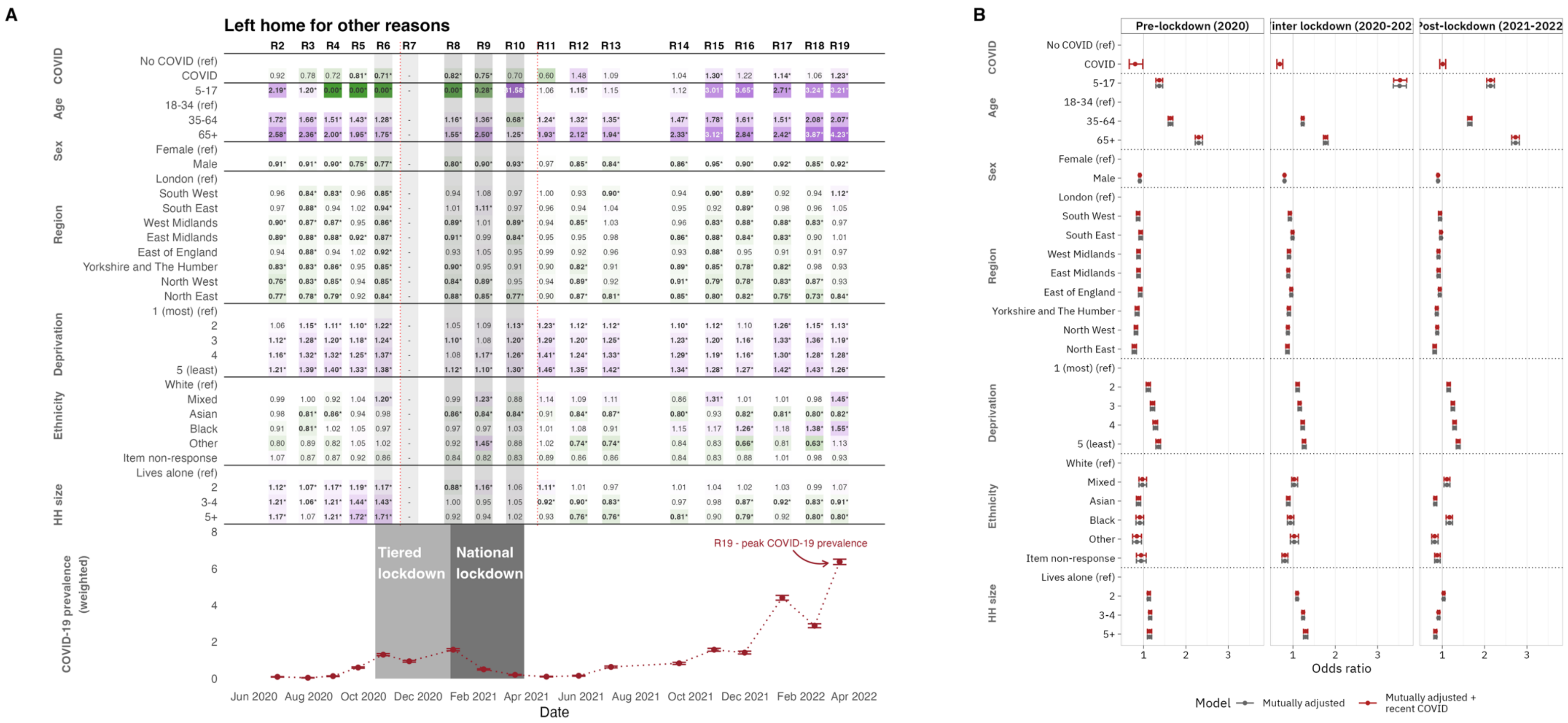

**Figure S35.** Odds ratios for whether an individual would report <u>leaving home for other reasons</u>. Estimated effects are mutually adjusted for the other variables listed in the table. * indicates the odds ratio is statistically significantly different from 1 at the $\alpha = 0.05$ significance level. This question was not asked in round 7, and the question wording changed between study rounds 2 and 3, and again between study rounds 14 to 15, denoted with a vertical dashed line. The change in question wording is particularly important here, as "other" may include or exclude specific reasons depending on the available options (see supplementary table S3).

**B.** Forest plot comparing odds ratios for models with and without adjustment for suspected recent COVID-19.

### Supplementary Section 4: Correlations between community-level mobility data and reported behaviour measures

Due to complications that could arise due to multicollinearity and autocorrelation in the behavioural data, here we report pairwise sample correlation coefficients between each of six community-level mobility measures and reported behaviour measures (Table S7). Overall, correlations are weaker in rounds 15 to 19, despite the random forest model often predicting mobility series just as well (Table 2).

**Table S7** Pairwise sample correlation coefficients between the six community-level mobility measures (Google data) and the proportions in REACT-1 survey rounds regarding not leaving the house or reporting particular reasons for leaving the house in the day before the survey was completed. These correlations were computes separately for rounds 2 to 6, rounds 8 to 14 and rounds 15 to 19. Positive correlations are shaded blue and negative correlations are shaded red, with stronger shades indicating stronger correlations.

#### a) Rounds 2 to 6

| | Grocery | Retail | Parks | Transit | Workplaces | Residential |
|---|---|---|---|---|---|---|
| Work | 0.49 | 0.30 | -0.38 | 0.23 | 0.47 | -0.32 |
| Volunteering | 0.44 | 0.48 | -0.14 | 0.49 | 0.68 | -0.56 |
| Medical appointment | 0.63 | 0.68 | -0.24 | 0.62 | 0.87 | -0.74 |
| Caring for someone | 0.63 | 0.35 | -0.48 | 0.16 | 0.51 | -0.29 |
| Socialise (in public) | -0.25 | 0.76 | 0.69 | 0.68 | -0.04 | -0.64 |
| Socialise (in private) | -0.42 | 0.61 | 0.82 | 0.62 | -0.23 | -0.52 |
| Exercise | -0.43 | -0.10 | 0.56 | 0.00 | -0.44 | 0.11 |
| Shopping | 0.42 | 0.89 | 0.02 | 0.75 | 0.62 | -0.83 |
| Errands | 0.34 | 0.95 | 0.15 | 0.83 | 0.60 | -0.90 |
| Other | 0.61 | 0.64 | -0.13 | 0.71 | 0.87 | -0.78 |
| Didn't leave | -0.29 | -0.87 | -0.30 | -0.82 | -0.53 | 0.86 |

#### b) Rounds 8 to 14

| | Grocery | Retail | Parks | Transit | Workplaces | Residential |
|---|---|---|---|---|---|---|
| Work | 0.30 | 0.34 | 0.32 | 0.34 | 0.26 | -0.32 |
| Volunteering | 0.84 | 0.90 | 0.74 | 0.91 | 0.88 | -0.90 |
| Medical appointment | 0.63 | 0.61 | 0.46 | 0.66 | 0.73 | -0.68 |
| Caring for someone | 0.72 | 0.82 | 0.74 | 0.80 | 0.72 | -0.79 |
| Exercise | -0.66 | -0.61 | -0.52 | -0.63 | -0.68 | 0.65 |
| Shopping | 0.74 | 0.84 | 0.73 | 0.81 | 0.69 | -0.79 |
| Errands | 0.89 | 0.96 | 0.82 | 0.96 | 0.87 | -0.94 |
| Didn't leave | -0.88 | -0.78 | -0.79 | -0.82 | -0.81 | 0.84 |

c) Rounds 15 to 19

| | Grocery | Retail | Parks | Transit | Workplaces | Residential |
|---|---|---|---|---|---|---|
| Work | 0.38 | 0.38 | 0.16 | 0.48 | 0.48 | -0.55 |
| Volunteering | 0.28 | 0.29 | 0.09 | 0.36 | 0.26 | -0.31 |
| Medical appointment | 0.54 | 0.58 | 0.21 | 0.72 | 0.65 | -0.73 |
| Caring for someone | 0.38 | 0.36 | -0.02 | 0.22 | 0.02 | -0.07 |
| Socialising (outside) | 0.38 | 0.45 | 0.76 | 0.67 | 0.32 | -0.76 |
| Socialising (inside) | 0.38 | 0.52 | 0.38 | 0.68 | 0.31 | -0.69 |
| Exercise | -0.19 | -0.15 | 0.25 | -0.07 | -0.14 | 0.05 |
| Shopping | 0.40 | 0.47 | -0.10 | 0.33 | 0.03 | -0.13 |
| Errands | 0.34 | 0.39 | 0.29 | 0.60 | 0.48 | -0.67 |
| Other | 0.05 | 0.00 | -0.17 | -0.18 | -0.22 | 0.31 |
| Walking a dog/pet | 0.07 | 0.11 | 0.12 | 0.07 | -0.11 | 0.00 |
| School/university | 0.01 | -0.03 | 0.21 | 0.26 | 0.46 | -0.45 |
| Holiday | 0.29 | 0.43 | 0.61 | 0.38 | -0.21 | -0.30 |
| Haven't left | -0.54 | -0.64 | -0.14 | -0.68 | -0.39 | 0.56 |

These results are very reassuring for cross-validation in terms of the magnitude and direction of the correlations. For example, for the mobility measure relating to time spent in residential locations, there are strong negative correlations with shopping, errand and medical appointments, for example, and strong positive correlations with “didn’t leave” the home. Similarly reporting “shopping” as a reason for leaving the home is strongly positively correlated with “grocery” and “retail” mobility measures.

### Supplementary Section 5: Stringency and mobility data

We also test the ability of the Google mobility data to model the stringency and containment indices using the same framework as we use to model the stringency and containment indices from REACT-data. Random forest regression is used to predict both the stringency and containment indices using the six Google mobility series.

To ensure comparisons with REACT-based models are as similar as possible, we only leverage data on days from which we have REACT data. We also fit two separate models for each index, reflecting the separate models estimated in the main paper. However, this is not an entirely like-for-like comparison, as we use four covariates in the primary analysis (four behavioural questions), whereas here we leverage all six covariates in the mobility series.

We find that estimates derived from REACT data are slightly outperformed by estimates derived from mobility data in rounds 2-to-5, when there is little overall change in the response. However, estimates derived from REACT data in rounds 6-to-19 substantially outperform those derived from mobility data.

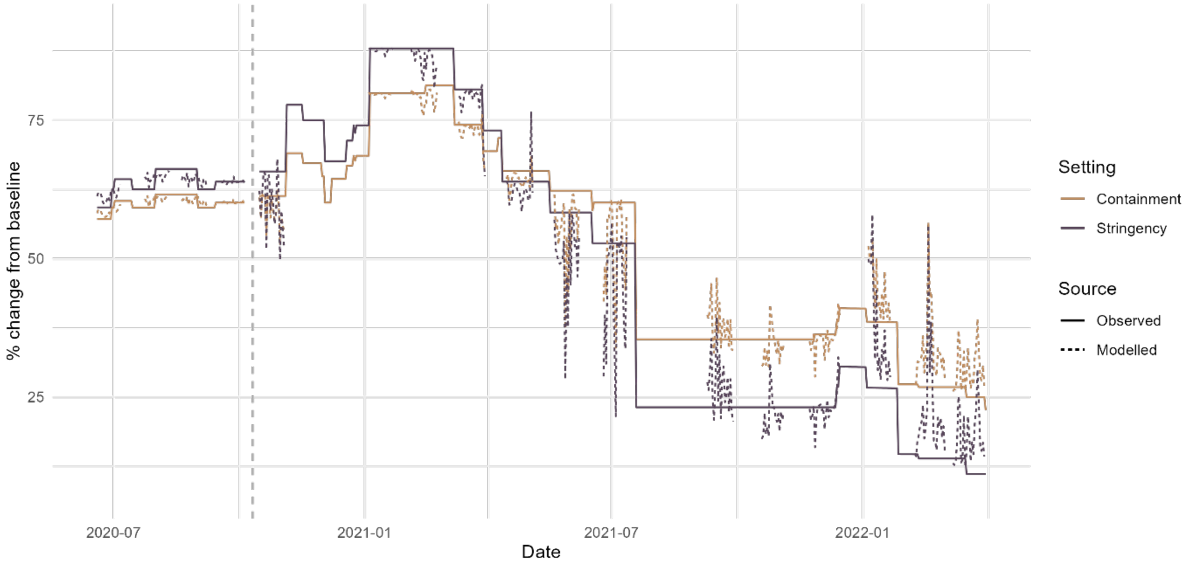

**Figure S36.** The OxCGRT stringency and containment and health indices (solid lines) between June 2020 and March 2022 and the model predictions based on Google mobility data. The vertical dashed lines demark individual model fits, selected to align with models fit in the primary analysis.

We also present the out-of-bag and within-bag proportion of variance explained for the four models considered in the main paper (three fitting mobility data and one fitting OxCGRT indices) in tables S8-S9.

**Table S8** Number of days analysed, sample variance and proportion of variance explained (PVE) for each of six mobility measures by the model (analogous to $R^2$) for out-of-bag prediction (where the model is trained on a subset of the data and performance is evaluated by predicting out of sample), and within-bag prediction.

| | | **Rounds 2-6** | **Rounds 8-14** | **Rounds 15-19** |
|---|---|---|---|---|
| *Number of days* | | 94 | 136 | 105 |
| Grocery | *Sample variance* | 3.12 | 130.36 | 12.55 |
| | Out-of-bag PVE | 0.73 | 0.96 | 0.68 |
| | Within-bag PVE | 0.95 | 0.99 | 0.95 |
| Retail | *Sample variance* | 132.72 | 464.09 | 32.56 |
| | Out-of-bag PVE | 0.96 | 0.97 | 0.81 |
| | Within-bag PVE | 0.99 | 0.99 | 0.97 |
| Parks | *Sample variance* | 811.22 | 1363.29 | 196.1 |
| | Out-of-bag PVE | 0.85 | 0.86 | 0.72 |
| | Within-bag PVE | 0.97 | 0.97 | 0.95 |
| Transit | *Sample variance* | 23.91 | 216.54 | 32.52 |
| | Out-of-bag PVE | 0.87 | 0.97 | 0.86 |
| | Within-bag PVE | 0.98 | 1 | 0.98 |
| Workplaces | *Sample variance* | 26.66 | 84.69 | 22.36 |
| | Out-of-bag PVE | 0.89 | 0.93 | 0.76 |
| | Within-bag PVE | 0.98 | 0.99 | 0.95 |
| Residential | *Sample variance* | 6.46 | 27.35 | 4.28 |
| | Out-of-bag PVE | 0.93 | 0.97 | 0.84 |
| | Within-bag PVE | 0.99 | 0.99 | 0.97 |

**Table S9. Number of days analysed, sample variance** and proportion of variation explained (PVE) by the model (analogous to $R^2$) for out-of-bag prediction (where the model is trained on a subset of the data and performance is evaluated by predicting out of sample), and within-bag prediction.

| | | Rounds 6-19 |
|---|---|---|
| *Number of days* | | *259* |
| Stringency | *Sample variance* | *760.14* |
| | Out-of-bag PVE | 0.91 |
| | Within-bag PVE | 0.98 |
| Containment | *Sample variance* | *381.75* |
| | Out-of-bag PVE | 0.90 |

| | | |
|---|---|---|
| | Within-bag PVE | 0.98 |

#### Supplementary Section 6: Social data portal

To enable future research, we have made aggregated data available in a portal [here]. This tool allows researchers to access estimates of the proportion of various groups of people in England that report performing various behaviours.

Results by round present survey-weighted estimates of the proportion of England that would report the selected outcome. Confidence intervals reflect uncertainty from extrapolating the survey to the general population. Daily results present the unweighted proportion of participants who reported the selected outcome. Confidence intervals are the 95% Binomial proportion interval, calculated using the “exact” method from the Hmisc package in R.

Users can select from a range of outcomes (e.g. shielding, left home for work, reports a previous positive test), grouping (e.g. age-group, ethnicity), and conditioning (e.g. those who report belief of risk of severe illness, a previous positive test).

Data are censored such that any single group with fewer than 10 responses, or fewer than 5 true and/or 5 false responses are excluded. This means we will never report proportions of 0% or 100% - any downstream analysis should ensure this is accounted for.

At the time of publication this portal does not contain all possible outcomes, although we plan on adding more over time. If you would benefit from the inclusion of additional outputs, please contact Professor Paul Elliott at Imperial College London.

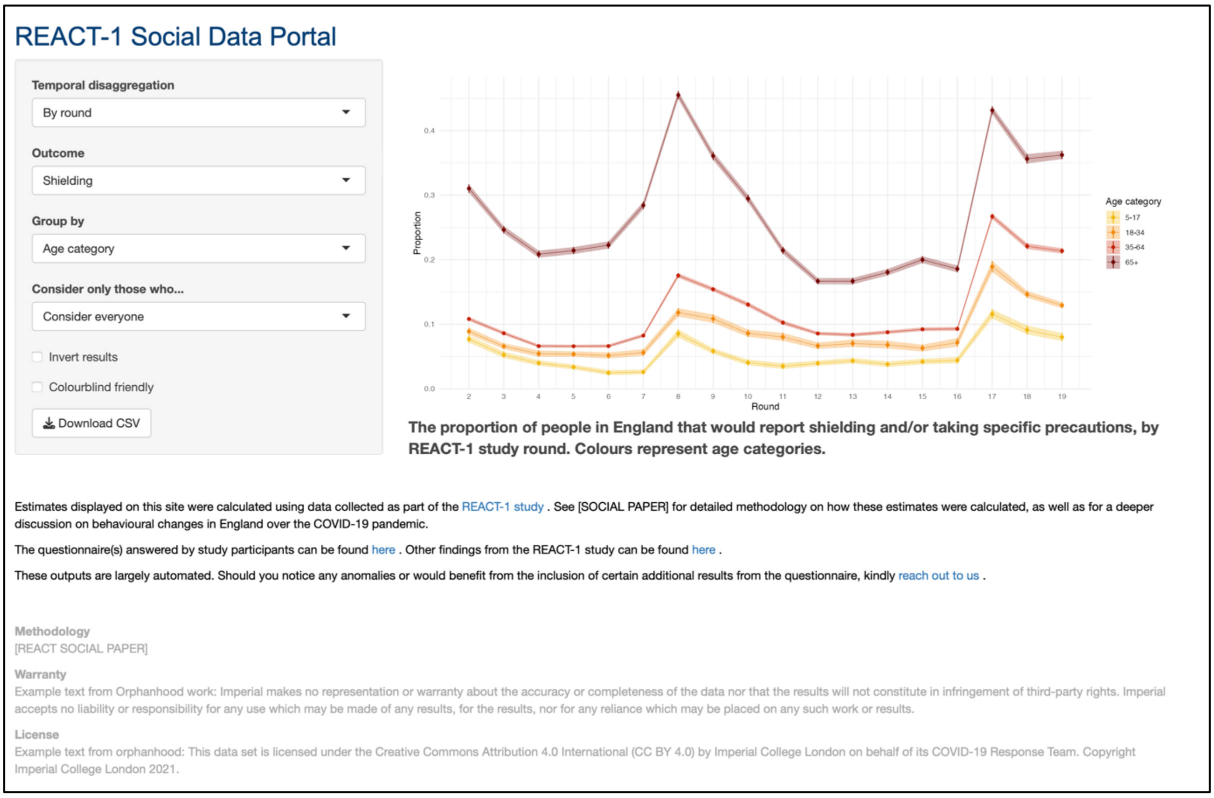

**Figure S37** An example window of the REACT-1 social data portal.
